# Quantifying the genetic separability of disease subtypes

**DOI:** 10.64898/2026.09.18.26363395

**Authors:** Chao Ning, Jasper Hof, Doug Speed

## Abstract

Genetic prediction of clinically defined disease subtypes could advance precision medicine, but polygenic risk scores (PRSs) often show limited ability to distinguish subtypes, either because the subtypes share genetic architecture or because available PRSs are inaccurate. We introduce GenSep, a statistical framework that estimates the oracle case–case AUC attainable if the true additive genetic values of both subtypes were known, and the proportion of this genetic separation recovered by subtype-specific PRSs. Across 18 subtype pairs in UK Biobank, oracle AUCs ranged from 0.605 to 1.000, whereas PRSs trained within UK Biobank achieved a median observed case–case AUC of only 0.543 (range, 0.521–0.752), reaching a median of only 26.8% (range, 9.7–50.4%) of the above-chance discrimination attainable at the oracle AUC. Oracle AUCs were generally consistent among UK Biobank, All of Us and FinnGen, and among European, African and Admixed American ancestry groups within the Million Veteran Program. PRSs trained on FinnGen GWAS, despite 2.5- to 3.8-fold larger discovery samples, showed no significant improvement over internally trained PRSs, whereas consortium-scale GWAS improved case–case AUC in three of four overlap-free comparisons but remained far below the oracle AUC. GenSep distinguishes subtype pairs whose discrimination is limited by intrinsic genetic overlap from those whose prediction is limited by current PRS accuracy, identifying where larger studies and better predictors could improve genetic discrimination.

## Introduction

Many diseases are clinically heterogeneous and are classified into subtypes based on symptom patterns^1^, etiological factors^2^, molecular profiles^3^, anatomical site^4^, age at onset^5^, disease course^6^ or treatment response^7^. Such classifications can predict disease course and guide treatment selection^8,9^. In practice, however, the correct subtype is not always evident when a patient presents, and for some diseases uncertain or incorrect classification can delay or misdirect treatment^10^. Genetic information could support classification when subtypes differ in their underlying genetic liabilities. For example, genetic risk scores can help distinguish type 1 diabetes from type 2 and monogenic diabetes^11,12^. However, clinical heterogeneity only partly reflects genetic heterogeneity: subtypes may share much of their genetic architecture^13,14^, which limits their genetic separability, that is, the extent to which cases of one subtype can be distinguished from those of another on the basis of genetic differences alone. Quantifying this separability can provide insight into disease biology^13^, support genetically informed subtype classification^15,16^, and indicate where genetic prediction can contribute to precision medicine^17^.

Previous studies have examined genetic differences among disease subtypes using several complementary approaches. Subtype-specific genome-wide association studies (GWAS) and tests of heterogeneity in variant effects can identify loci whose effects differ across clinically defined subgroups^18^. Global tests of genome-wide association statistics can assess whether subsets of variants have different effects between disease subgroups^19^, whereas genetic correlations quantify the similarity of genome-wide variant effects between the subgroups^20^. Case–case association methods directly identify variants with different allele frequencies between cases of related diseases or subtypes^21^. More recently, the Genetic DIstance of disorder Subtypes (GDIS) framework combines genome-wide heritability and genetic correlation estimates to infer genetic distance in direct subtype–subtype comparisons^22^. At the individual level, polygenic risk scores (PRSs) aggregate effects across variants and have been used to distinguish clinically overlapping conditions^11^. Multivariate approaches combine disorder-specific PRSs while accounting for their correlations to estimate the probability of each diagnosis^23^. Although these approaches show that genetic information can help distinguish disease subtypes, the maximum discrimination attainable from genetic variation remains unclear, as does the extent to which current prediction models capture this potential.

Current genome-wide PRSs often provide only limited or moderate discrimination between disease subtypes or clinically related disorders^23,24^. This may reflect substantial overlap in the genetic architectures of the subtypes, which limits how well genetic information can distinguish between them^13^. Alternatively, the subtypes may be genetically separable, but available PRSs may recover only a fraction of this separation because finite discovery GWAS sample sizes limit the accuracy of estimated variant effects and hence of predicted genetic liabilities^25^. Distinguishing these explanations requires comparing the discrimination achieved by available PRSs with the maximum discrimination supported by the underlying genetic architecture. Previous work has related the maximum attainable case–control discrimination of a genomic prediction model to disease prevalence and liability-scale heritability^26^, but extending this result to subtype discrimination requires jointly accounting for the prevalence and heritability of each subtype, as well as their genetic correlation.

Here we introduce GenSep, a statistical framework that quantifies the genetic separability of two clinically defined disease subtypes. GenSep uses the SNP heritability and population prevalence of each subtype, as well as the genetic correlation between the two subtypes, to estimate the oracle case–case area under the receiver operating characteristic curve (AUC), which represents the maximum subtype discrimination attainable from additive genetic effects under the liability-threshold model^27^ when the true additive genetic values underlying both subtypes are known. By incorporating subtype-specific PRSs, GenSep also quantifies the proportion of the above-chance oracle AUC achieved by available predictors (AUC recovery). We validated GenSep through simulations and applied it to 18 disease-subtype pairs in UK Biobank^28^. We then assessed the consistency of oracle AUC estimates across biobanks and ancestries, using the All of Us Research Program^29^, FinnGen^30^ and the Million Veteran Program (MVP)^31^, and evaluated how much PRSs based on larger external discovery GWAS can improve case–case discrimination.

## Results

### The GenSep framework: genetic ceiling and PRS recovery

GenSep addresses two questions for a pair of clinically defined subtypes: (1) how well the subtypes could be separated if the additive genetic value of every case were known, and (2) how much of that separation current PRSs achieve. **Fig. 1** summarizes the key equations from the GenSep framework; full derivations are given in **Methods** and **Supplementary Notes 1** and **2**. To answer the first question, GenSep considers an optimized genetic difference score, *D* = *w*_1_*g*_1_ − *w*_2_*g*_2_, where *g*_1_ and *g*_2_ are the standardized additive genetic values for subtypes 1 and 2, respectively, and the weights *w*_1_ and *w*_2_ are chosen to maximize how well *D* discriminates between the two subtypes. GenSep then computes two quantities. The genetic separation variance, *V*_S_, measures the distance between the mean values of *D* in the two subtype groups (**Methods**): it is 0 when the groups cannot be distinguished based on their additive genetic values and increases as they become more distinct. Meanwhile, the oracle case–case AUC (AUC_oracle_) is the probability that a randomly picked subtype-1 case has a higher value of *D* than a randomly picked subtype-2 case. We compute AUC_oracle_ analytically under the liability-threshold model (the analytical oracle AUC; equation (2), **Methods**), and **Fig. 1** also reports a simplified approximation (equation (3), **Methods**), which shows that AUC_oracle_ depends mainly on *V*_S_.

**Fig. 1:**
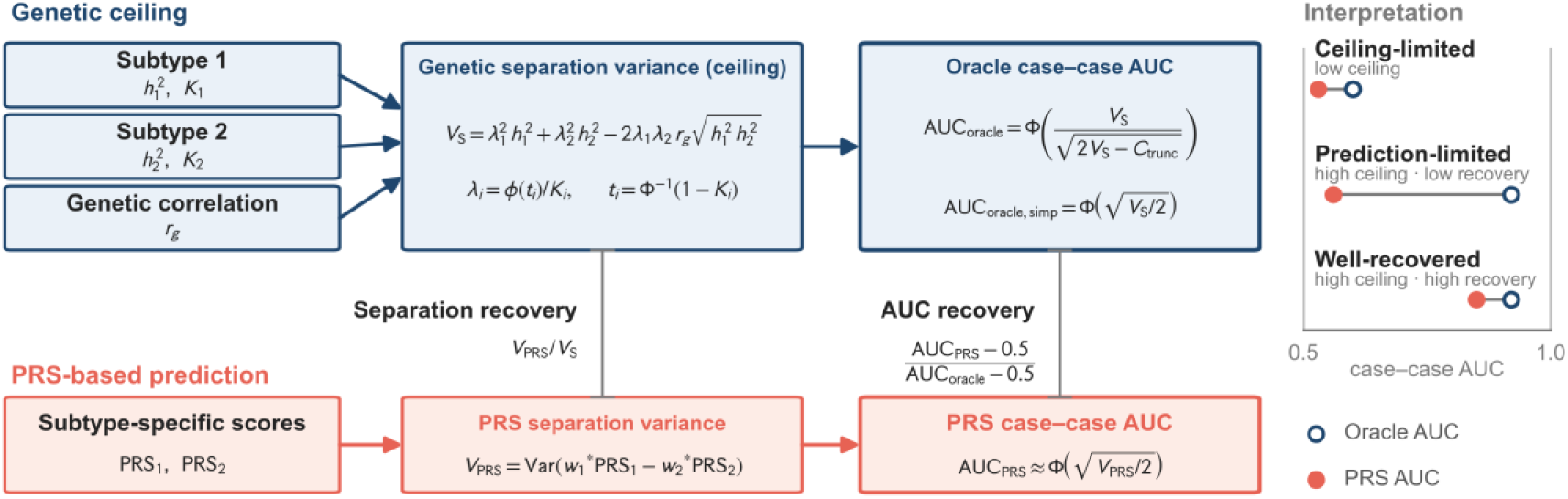
Framework for the genetic ceiling and PRS recovery in disease-subtype discrimination. The upper branch defines the oracle AUC from subtype-specific liability-scale SNP heritabilities, 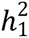 and 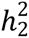, population prevalences, *K*_1_ and *K*_2_, and genetic correlation, *r*_g_. These quantities determine the liability thresholds, *t*_1_ and *t*_2_, selection intensities, *λ*_1_ and *λ*_2_, genetic separation variance, *V*_S_, and the oracle case– case AUC (AUC_oracle_). The truncation term, *C*_trunc_, accounts for the reduction in the sum of the within-subtype variances induced by liability-threshold ascertainment. The simplified approximation, 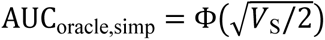, provides a simple link between genetic separation variance and the oracle AUC. The lower branch represents current prediction using subtype-specific PRSs, PRS_1_ and PRS_2_, which are combined into an optimal linear case–case score, 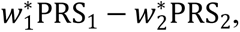, where 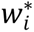 are the optimal PRS weights. Its separation variance, *V*_PRS_, is the PRS analogue of *V*_S_ and determines the PRS case–case AUC through the same simplified approximation, 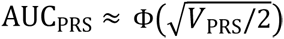. Vertical lines denote separation recovery, *V*_PRS_/*V*_S_, the fraction of the genetic separation variance captured by the PRS-based score, and AUC recovery, (AUC_PRS_ - 0.5)/(AUC_oracle_ - 0.5), the fraction of the above-chance oracle AUC achieved by the PRS-based score. Right, schematic examples of the three categories, ceiling-limited (low ceiling), prediction-limited (high ceiling, low AUC recovery) and well-recovered (high ceiling, high AUC recovery), with open blue circles denoting oracle AUC and filled orange circles denoting PRS AUC.

GenSep answers the second question in a similar way, except that it now considers scores where *g*_1_ and *g*_2_ are replaced by PRS-based estimates of the additive genetic values for each subtype, with the weights re-derived for these scores (**Supplementary Note 2**). The resulting quantities are *V*_PRS_ and AUC_PRS_, which describe how well we can distinguish the subtype groups based on PRSs. Note that *V*_S_ and AUC_oracle_ depend on five parameters (the SNP heritabilities and prevalences of each subtype, as well as their genetic correlation), while *V*_PRS_ and AUC_PRS_ depend on seven parameters (the five listed above, and also the accuracy of PRS for each subtype).

We refer to *V*_S_ and AUC_oracle_ as genetic ceilings, because they are upper bounds for *V*_PRS_ and AUC_PRS_, respectively. We compare the PRS-based quantities with their ceilings by computing the separation recovery *V*_PRS_/*V*_S_, which is the fraction of the available genetic separation that the scores capture, and the AUC recovery (AUC_PRS_ − 0.5)/(AUC_oracle_ − 0.5), which is the fraction of the above-chance oracle AUC that the PRSs achieve. Because the two measures are closely related (AUC recovery is approximately the square root of separation recovery; **Supplementary Note 3**), we report AUC recovery throughout. Furthermore, the oracle AUC and AUC recovery can be used to assign each subtype pair to one of three categories (**Fig. 1**, right): ceiling-limited (low ceiling), prediction-limited (high ceiling, low AUC recovery) and well-recovered (high ceiling, high AUC recovery).

## Simulations

We performed extensive simulations to validate GenSep (see **Methods** for simulation details). We first examined the oracle AUC across a grid of subtype heritabilities, prevalences, genetic correlations and residual correlations. For each parameter setting, we generated cases for each subtype under the bivariate liability-threshold model, and calculated the empirical oracle AUC using a score that optimally combined the true genetic values for the two subtypes.

Across the full parameter grid, oracle AUC was largely determined by the genetic separation variance *V*_S_. Both empirical and analytical oracle AUCs increased almost monotonically with *V*_S_, as expected from the simplified approximation (**Fig. 2a**). The analytical oracle AUC closely matched the empirical oracle AUC (mean absolute error, 0.0005), whereas the simplified approximation captured the main monotonic trend but was slightly conservative at intermediate *V*_S_ values (**Fig. 2a**, **Supplementary Figs. 1** and **2**).

**Fig. 2:**
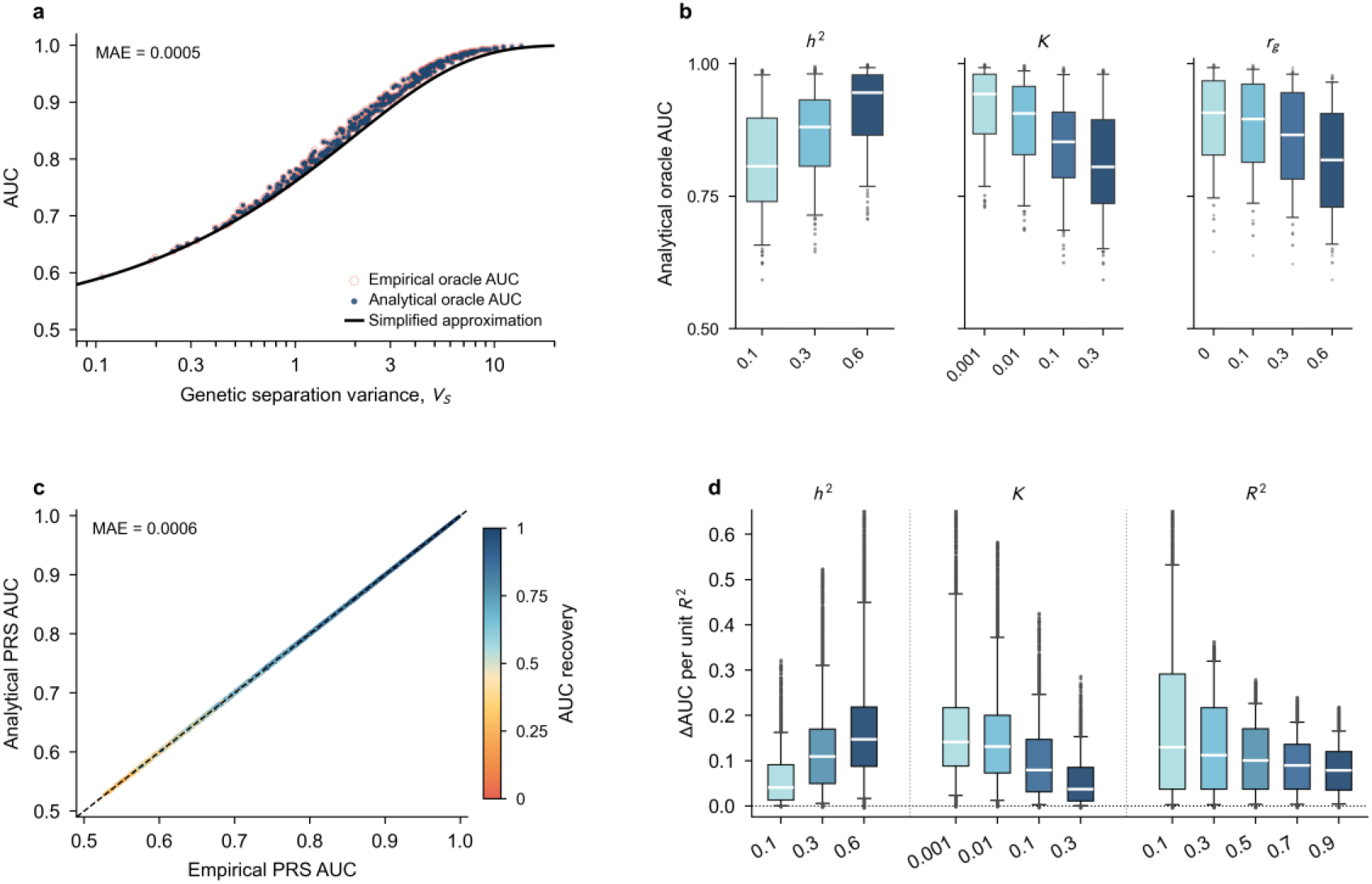
Validation of the oracle AUC and PRS recovery in simulations. **a,** Oracle AUC as a function of genetic separation variance *V*_S_ (log scale) in simulations. Open red circles denote oracle AUC measured directly from simulated case sets, filled blue points denote analytical oracle AUC, and the black curve denotes the simplified approximation. Each point is one of the 576 settings of the simulation grid (Methods); empirical values are means over 30 replicates of 5,000 cases per subtype. MAE denotes the mean absolute difference between the analytical and empirical AUCs (also in **c**). **b,** Distributions of the analytical oracle AUC over the simulation grid, shown separately by subtype liability heritability *h*^2^, prevalence *K* and cross-subtype genetic correlation *r*_g_. The *h*^2^ and *K* sections pool the two subtype arms because the model is symmetric between subtypes, and boxes are shaded light to dark across the levels of their own group. **c,** Analytical PRS-based case–case AUC plotted against the empirical AUC measured directly from simulated case sets. Each point denotes one of 20,736 parameter combinations spanning subtype heritabilities (*h*^2^), prevalences (*K*), genetic correlation (*r*_g_) and per-subtype PRS accuracies (*R*^2^). Points are colored by AUC recovery, the fraction of the above-chance oracle AUC achieved by the PRS-based score. Empirical values are means over 30 replicates of approximately 5,000 cases per subtype, evaluated at the same optimal PRS weights as the analytical values (Supplementary Fig. 6); the dashed line denotes identity. **d,** Gain in PRS-based case–case AUC after increasing PRS accuracy, measured as ΔAUC/Δ*R*^2^ between adjacent *R*^2^ grid levels while holding all other parameters fixed. Effects are stratified by the heritability *h*^2^, prevalence *K* and baseline PRS accuracy *R*^2^ (before the increment) of the subtype whose PRS accuracy was increased. Boxes in **b** and **d** show medians, interquartile ranges, 5th–95th percentile whiskers and individual outlying points; they summarize the spread of the parameter grid rather than sampling error, and the dotted line in **d** denotes zero gain. Each box in **b** summarizes 384 (*h*^2^), 288 (*K*) or 144 (*r*_g_) analytical values, with the two subtype arms pooled for *h*^2^ and *K*; each box in **d** summarizes 11,520 (*h*^2^), 8,640 (*K*) or 6,912 (*R*^2^) one-step increments. No statistical tests were performed.

Oracle AUC generally increased with subtype heritability but decreased with subtype prevalence and genetic correlation (**Fig. 2b** and **Supplementary Fig. 3**). For example, oracle AUC reached a maximum of 0.999 when both subtypes had high heritability 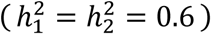, low prevalence (*K*_1_ = *K*_2_ = 0.001), and no genetic correlation (*r*_g_ = 0). Conversely, when heritability was low 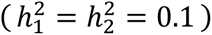, prevalence was high (*K*_1_ = *K*_2_ = 0.3), and genetic correlation was strong (*r*_g_ = 0.6), oracle AUC dropped to 0.59. Residual correlation (*r*_e_) had negligible effect on oracle AUC: changing *r*_e_ while keeping the other parameters fixed produced nearly identical estimates (**Supplementary Fig. 4**), which is consistent with the analytical formula being independent of *r*_e_.

We further confirmed that genetic separation variance and oracle AUC could be accurately estimated using subtype-specific SNP heritabilities and cross-subtype genetic correlations derived from GWAS summary statistics. Substituting SumHer^32^ estimates of heritability and genetic correlation into the analytical formulas closely recovered the corresponding values calculated from the prespecified simulation parameters, with similar accuracy when all controls were used and when controls were subsampled to 1:1 or 1:4 case–control ratios (Pearson *r* = 0.995–0.998 for the genetic separation variance, **Supplementary Fig. 5a–c**; 0.988–0.996 for the oracle AUC, **Supplementary Fig. 5d–f)**. These findings support the use of GWAS summary statistics, together with prespecified subtype prevalences, to estimate oracle AUC in subsequent real-data analyses.

We next validated the finite-accuracy PRS extension by adding the accuracy of each subtype-specific PRS to the simulation grid. Across the parameter grid, PRS-based case–case AUCs predicted by the analytical formula, which replaces the true genetic values by scores of known accuracies (**Methods** and **Supplementary Note 2**), closely matched those calculated directly from the simulated data (**Fig. 2c** and **Supplementary Figs. 6–8**).

Improving the PRS for a single subtype yielded variable gains in case–case discrimination across genetic architectures (**Fig. 2d**). The marginal increase in case– case AUC per unit improvement in PRS accuracy was greater for subtypes with higher heritability and lower prevalence, but diminished as baseline PRS accuracy increased. These patterns held across levels of cross-subtype genetic correlation (**Supplementary Fig. 9**). At higher genetic correlations, the relative benefit of improving one subtype’s PRS over the other depended more strongly on their heritabilities and prevalences and less on their baseline PRS accuracies. Thus, the benefit of improving a subtype-specific PRS depends on both its underlying genetic architecture and current predictive accuracy, rather than on predictive accuracy alone.

### Genetic ceilings and PRS recovery across 18 UK Biobank subtype pairs

We applied GenSep to 18 disease-subtype pairs in UK Biobank, defined by ICD-10 diagnosis codes (**Supplementary Table 1**). For each pair, we estimated the SNP heritability of each subtype and their cross-subtype genetic correlation, and then derived the genetic separation variance and oracle AUC (**Supplementary Table 2**). Genetic separation variance ranged from 0.13 for acute myocardial infarction versus chronic ischemic heart disease to 12.5 for type 1 versus type 2 diabetes (median, 0.68). Oracle AUCs varied widely across pairs (range, 0.605–1.000; median, 0.725), with 11 of 18 contrasts reaching acceptable discrimination and three of these reaching excellent discrimination, using the conventional thresholds of AUC > 0.70 and > 0.80, respectively^33^. The highest genetic separation was observed for type 1 versus type 2 diabetes, hypothyroidism versus hyperthyroidism and Crohn disease versus ulcerative colitis, with oracle AUCs of 1.000, 0.914 ± 0.027 and 0.822 ± 0.051 (estimate ± block-jackknife standard error for the last two), respectively, with the first estimate subject to a boundary effect. Its interpretation is limited by the large uncertainty of the type 1 diabetes heritability estimate: varying the heritability across the estimates from other cohorts and definitions gave oracle AUCs from 0.88 to 1.00 (**Supplementary Note 3**). These estimates were robust to the assumed population prevalences: halving or doubling the prevalence of either or both subtypes changed the oracle AUC by at most 0.018 (**Supplementary Note 3** and **Supplementary Fig. 10**).

We next assessed how much of the oracle AUC was recovered by subtype-specific PRSs constructed using MegaPRS^34^. PRS-based case–case AUCs predicted by the analytical formula were in close empirical agreement with those observed in UK Biobank (Pearson *r* = 0.975). The calibration slope (ordinary least-squares regression of observed on predicted AUC, with intercept) was 0.98, with a mean absolute error of 0.013 (**Supplementary Fig. 11**). Observed case–case AUCs were below their oracle values for all 18 pairs (**Fig. 3a** and **Supplementary Table 3**). They ranged from 0.521 to 0.752 (median, 0.543), and only type 1 versus type 2 diabetes (0.752) and hypothyroidism versus hyperthyroidism (0.649) exceeded 0.60; AUC recovery ranged from 0.097 to 0.504 (median, 0.268; **Fig. 3a**), and only type 1 versus type 2 diabetes reached 0.50.

**Fig. 3:**
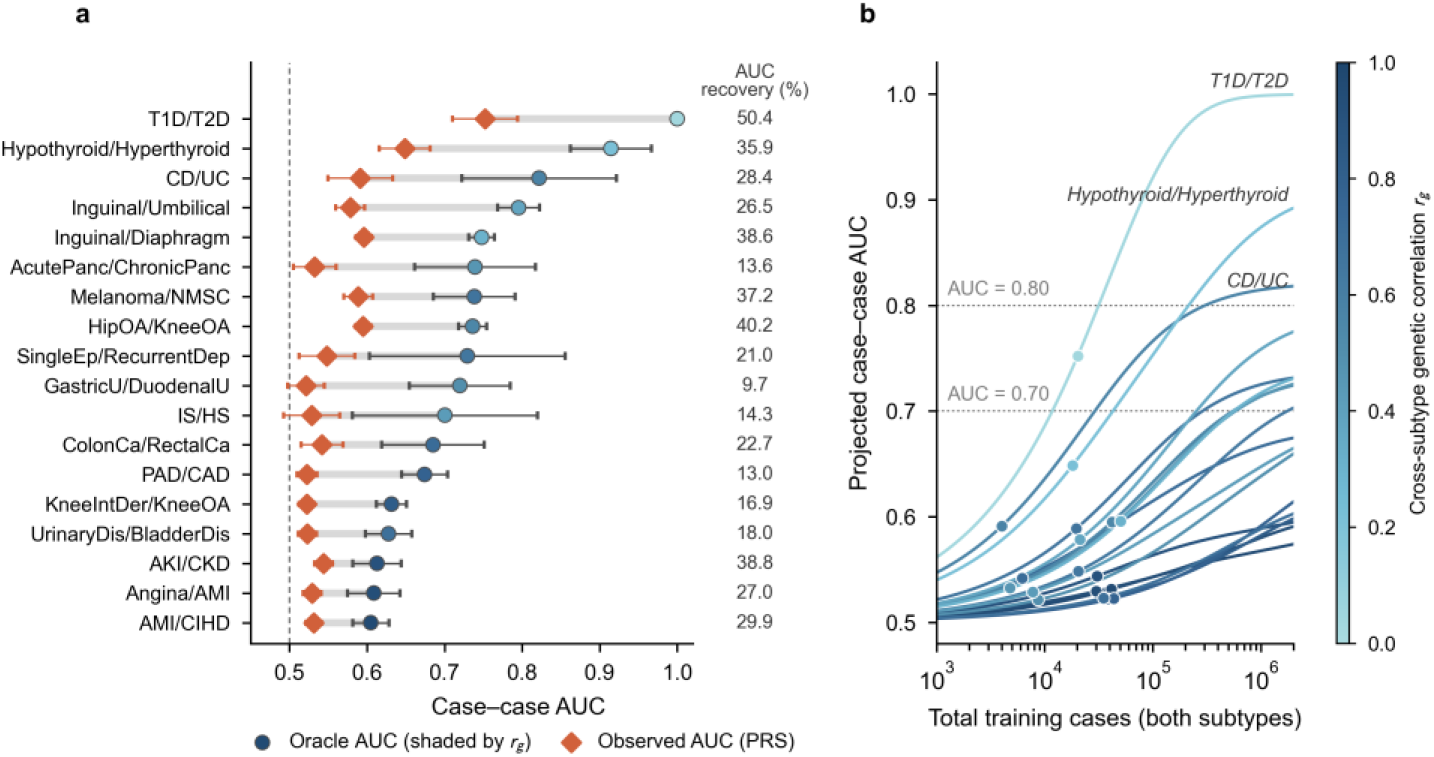
Oracle and observed case–case AUCs for 18 subtype pairs in UK Biobank, with sample-size projections. **a,** Oracle and observed AUCs for 18 disease-subtype pairs (case numbers in Supplementary Table 1) in UK Biobank. Circles denote the oracle AUC, the discrimination achievable by the genetic discriminant if the true additive genetic values were observed (Methods), with markers shaded by genetic correlation; error bars denote ±1.96 block-jackknife s.e. for oracle AUC. Observed PRS-based case– case AUCs in held-out UK Biobank test data are shown as diamonds. They were evaluated in the 20% held-out test set of each pair; error bars denote ±1.96 block-jackknife s.e. over evaluation individuals (Methods). Grey horizontal connectors span from the observed AUC to the oracle AUC, showing the gap due to finite PRS accuracy. The dashed vertical line denotes AUC = 0.5. The column at the right gives AUC recovery, (AUC_PRS_ − 0.5)/(AUC_oracle_ − 0.5) (%). **b,** Projected observed case–case AUC as a function of the total number of training cases across the two subtypes (log scale), assuming an unlimited number of controls. Each curve is one subtype pair, colored by its cross-subtype genetic correlation; the filled marker indicates the current UK Biobank training size, and dotted horizontal lines mark AUC = 0.70 and 0.80. Each curve is anchored at the AUC currently observed in held-out test data and asymptotes at that pair’s oracle AUC.

To estimate the training sample sizes needed to reach target AUCs, we projected case–case AUC as a function of the total number of training cases across the two subtypes, assuming an unlimited number of controls (**Fig. 3b**). Under this projection, the AUC approaches each pair’s oracle value as the training samples of both subtypes grow. When both samples grew in proportion to their current sizes, all 11 pairs with oracle AUCs above 0.70 were projected to reach an AUC of 0.70, nine of them with at most two million in total across the two subtypes. An AUC of 0.80 could be reached by increasing the training cases of only the smaller subtype for type 1 versus type 2 diabetes (approximately 940 type 1 diabetes cases, with type 2 diabetes at its current 20,000) and for hypothyroidism versus hyperthyroidism (approximately 36,000 hyperthyroidism cases, with hypothyroidism at its current 17,000), whereas Crohn disease versus ulcerative colitis required both subtypes to grow, to approximately 98,000 and 207,000 cases under proportional growth (**Supplementary Table** 4).

### Cross-cohort and cross-ancestry comparison of genetic ceilings

To assess the reproducibility of these oracle AUCs across cohorts and ancestries, we repeated the analysis in All of Us, FinnGen and the MVP (the latter providing European, African and Admixed American analyses) (**Fig. 4**, **Table 1** and **Supplementary Table 2**). Each comparison was restricted to subtype pairs with reliable estimates in both analyses, that is, positive heritability estimates for both subtypes and a genetic correlation estimate not on an estimator boundary (**Supplementary Note 3**). The comparisons were organized along two dimensions: first among European-ancestry analyses of different cohorts, which also differ in phenotype recording; then among ancestry groups within MVP, with the cohort and phenotyping system shared.

**Fig. 4:**
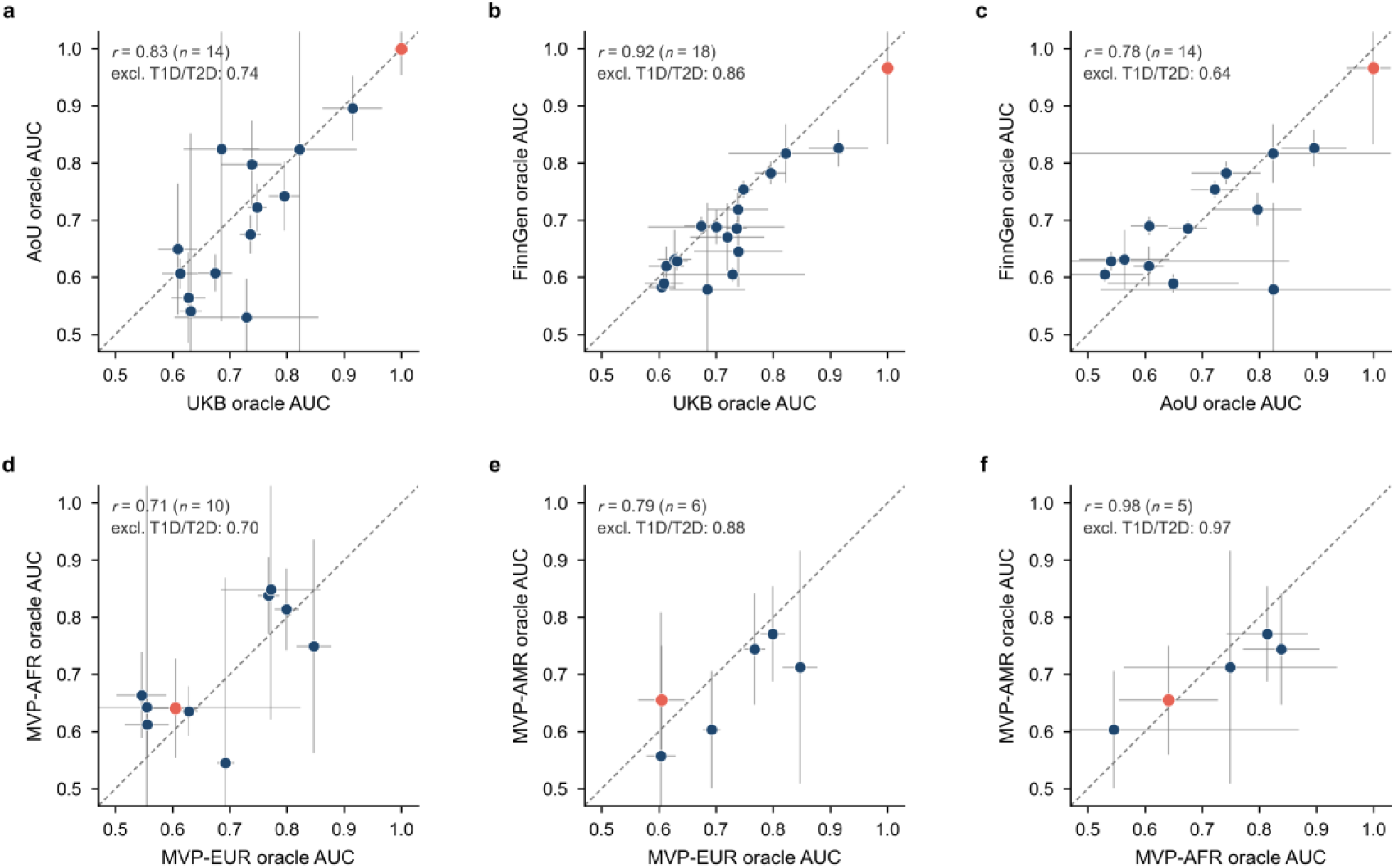
Oracle AUCs across cohorts and ancestries. Oracle case–case AUCs were compared across subtype pairs with reliable estimates in both analyses. Each point is one subtype pair, and panels carry different numbers of points because a pair enters only where it is reliable in both analyses; all pairs are shown in blue except T1D/T2D, highlighted in red. Dashed lines denote identity, and error bars represent ±1.96 block-jackknife s.e. obtained via 200 genomic blocks, around the point estimates; *n* = 14 (**a**), 18 (**b**), 14 (**c**), 10 (**d**), 6 (**e**) and 5 (**f**) subtype pairs. Each panel reports the Pearson correlation, *r*, across all shared pairs and after excluding T1D/T2D. Pearson correlations are descriptive and were not tested; per-pair differences are tested in Supplementary Figs. 14 and 15. Top row, comparisons among the European-ancestry analyses of the three biobanks. **a,** All of Us (AoU) versus UK Biobank (UKB). **b,** FinnGen versus UKB. **c,** FinnGen versus AoU. Bottom row, cross-ancestry comparisons within MVP. **d,** African-ancestry (MVP-AFR) versus European-ancestry analyses. **e,** Admixed American (MVP-AMR) versus European-ancestry analyses. **f,** Admixed American versus African-ancestry analyses.

**Table 1:** Oracle case–case AUC for the 18 subtype pairs across cohorts and ancestries.

| Subtype pair | Abbreviation | UK Biobank | All of Us | FinnGen | MVP-EUR | MVP-AFR | MVP-AMR |
| --- | --- | --- | --- | --- | --- | --- | --- |
| Type 1 diabetes mellitus – Type 2 diabetes mellitus | T1D/T2D | 1.000 (0.002) | 0.999 (0.024) | 0.966 (0.068) | 0.605 (0.021) | 0.641 (0.044) | 0.655 (0.049) |
| Hypothyroidism – Hyperthyroidism / thyrotoxicosis | Hypothyroid/Hyperthyroid | 0.914 (0.027) | 0.895 (0.029) | 0.826 (0.017) | 0.847 (0.015) | 0.749 (0.095) | 0.713 (0.104) |
| Crohn disease – Ulcerative colitis | CD/UC | 0.822 (0.051) | 0.824 (0.198) | 0.817 (0.026) | 0.699 (0.049) | — | — |
| Inguinal hernia – Umbilical hernia | Inguinal/Umbilical | 0.795 (0.014) | 0.742 (0.031) | 0.782 (0.010) | 0.799 (0.011) | 0.814 (0.036) | 0.771 (0.043) |
| Inguinal hernia – Diaphragmatic hernia | Inguinal/Diaphragm | 0.748 (0.008) | 0.722 (0.021) | 0.754 (0.008) | 0.768 (0.009) | 0.838 (0.034) | 0.744 (0.050) |
| Acute pancreatitis – Chronic pancreatitis | AcutePanc/ChronicPanc | 0.739 (0.040) | — | 0.645 (0.032) | 0.555 (0.137) | 0.642 (0.296) | — |
| Malignant melanoma of skin – Non-melanoma skin cancer | Melanoma/NMSC | 0.738 (0.027) | 0.797 (0.039) | 0.719 (0.015) | 0.571 (0.011) | — | — |
| Hip osteoarthritis / coxarthrosis – Knee osteoarthritis / gonarthrosis | HipOA/KneeOA | 0.736 (0.009) | 0.675 (0.017) | 0.686 (0.006) | — | — | — |
| Depressive episode – Recurrent depressive disorder | SingleEp/RecurrentDep | 0.729 (0.064) | 0.530 (0.034) | 0.605 (0.006) | — | — | — |
| Gastric ulcer – Duodenal ulcer | GastricU/DuodenalU | 0.720 (0.033) | — | 0.670 (0.030) | 0.772 (0.044) | 0.848 (0.116) | — |
| Ischemic stroke / cerebral infarction – Hemorrhagic stroke | IS/HS | 0.700 (0.061) | — | 0.687 (0.015) | 0.641 (0.111) | — | — |
| Colon cancer – Rectal cancer | ColonCa/RectalCa | 0.685 (0.034) | 0.824 (0.154) | 0.579 (0.077) | 0.586 (0.095) | — | — |
| Peripheral artery disease – Coronary artery disease | PAD/CAD | 0.674 (0.015) | 0.607 (0.016) | 0.690 (0.008) | 0.693 (0.008) | 0.545 (0.166) | 0.603 (0.052) |
| Internal derangement of knee – Knee osteoarthritis / gonarthrosis | KneeIntDer/KneeOA | 0.632 (0.010) | 0.541 (0.159) | 0.628 (0.009) | — | — | — |
| Other disorders of urinary system – Other disorders of bladder | UrinaryDis/BladderDis | 0.628 (0.015) | 0.564 (0.040) | 0.631 (0.026) | 0.555 (0.019) | 0.612 (0.029) | — |
| Acute kidney injury / acute kidney failure – Chronic kidney disease | AKI/CKD | 0.613 (0.016) | 0.606 (0.013) | 0.619 (0.018) | 0.628 (0.008) | 0.635 (0.022) | — |
| Angina pectoris – Acute myocardial infarction | Angina/AMI | 0.609 (0.017) | 0.649 (0.058) | 0.589 (0.008) | 0.604 (0.013) | — | 0.557 (0.128) |
| Acute myocardial infarction – Chronic ischemic heart disease | AMI/CIHD | 0.605 (0.012) | — | 0.583 (0.005) | 0.546 (0.022) | 0.663 (0.038) | — |

Values are oracle case–case AUC with block-jackknife standard error in parentheses. MVP-EUR, MVP-AFR and MVP-AMR denote the European, African and Admixed American ancestry analyses of the Million Veteran Program. A dash means that no estimate is reported, either because the estimate failed quality control (a non-positive subtype heritability, or a cross-subtype genetic correlation that is missing or lies on the $\pm 0.999$ estimator boundary) or because the cohort cannot reach that pair: MVP phecodes do not preserve the ICD-10 distinctions used for HipOA/KneeOA, KneeIntDer/KneeOA or the depression contrast, no Admixed American summary statistics are released for six further phecodes, and MVP African has too few melanoma cases.

Oracle AUCs were generally consistent among the European-ancestry analyses of the three biobanks: Pearson *r* = 0.83 between All of Us and UK Biobank (14 pairs), 0.92 between FinnGen and UK Biobank (18 pairs) and 0.78 between FinnGen and All of Us (14 pairs), and 0.74, 0.86 and 0.64, respectively, after excluding type 1 versus type 2 diabetes, whose near-perfect oracle AUC strongly influences the correlations (**Fig. 4a–c**).

Agreement was weaker between MVP European and UK Biobank (*r* = 0.44, 15 pairs; **Supplementary Figs. 12** and **13a**) but increased to 0.71, within the range of the three biobanks, after excluding type 1 versus type 2 diabetes. Agreement with UK Biobank was weak both for the genetic separation variance (*r* = 0.04, driven by its very large value for type 1 versus type 2 diabetes in UK Biobank; Spearman *ρ* = 0.54) and the genetic correlation (*r* = 0.48), and improved to 0.80 and 0.72 after excluding type 1 versus type 2 diabetes (**Supplementary Fig. 12** and **Supplementary Table 5**). Under the MVP phecode definitions (250.1 and 250.2), the two diabetes subtypes are near-indistinguishable genetically (*r*_g_ = 0.95 ± 0.02 in MVP European, against 0.04 ± 0.04 in UK Biobank), and the oracle AUC for this pair falls from 1.000 in UK Biobank to 0.605 in MVP European (**Supplementary Fig. 13b** and **Supplementary Tables 2** and **6**). Broadening the diabetes case definitions to any ICD-10 code starting with E10 and any code starting with E11, respectively, raised the genetic correlation from 0.04 to 0.56 in UK Biobank and from −0.03 to 0.43 in All of Us, and lowered the oracle AUC from 1.000 to 0.871 and from 0.999 to 0.890 (**Supplementary Fig. 13b**).

Of the 87 per-pair differences in oracle AUC among the European-ancestry analyses of the four cohorts, 14 were significant after Benjamini–Hochberg correction (*q* < 0.05), 11 of them in four pairs: type 1 versus type 2 diabetes, melanoma versus non-melanoma skin cancer, peripheral artery versus coronary artery disease and hip versus knee osteoarthritis (**Supplementary Fig. 14** and **Supplementary Table 7**).

Varying ancestry within MVP while holding the cohort and phenotyping constant, oracle AUCs were correlated at *r* = 0.71 between European and African (10 pairs), 0.79 between European and Admixed American (6 pairs) and 0.98 between African and Admixed American (5 pairs); excluding type 1 versus type 2 diabetes gave 0.70, 0.88 and 0.97, suggesting that this definitional issue was unlikely to differentially affect comparisons within MVP (**Fig. 4d–f** and **Supplementary Table 5**). No per-pair difference reached *q* < 0.05 in the 21 comparisons of oracle AUC (**Supplementary Fig. 15**). Thus, within this limited set of 5 to 10 eligible subtype pairs, we detected no statistically significant ancestry-related differences in oracle AUC.

### Transferability of subtype-specific PRSs trained in external biobank and consortium GWAS

The extent to which the observed case–case AUC approaches the oracle AUC depends partly on discovery GWAS sample size, which is often limited within a single target cohort. We therefore evaluated whether larger external discovery GWAS could narrow this gap. For each subtype pair, we compared subtype-specific PRSs constructed using GWAS summary statistics from the target cohort itself, FinnGen and published consortium meta-analyses (**Fig. 5a,b**).

**Fig. 5:**
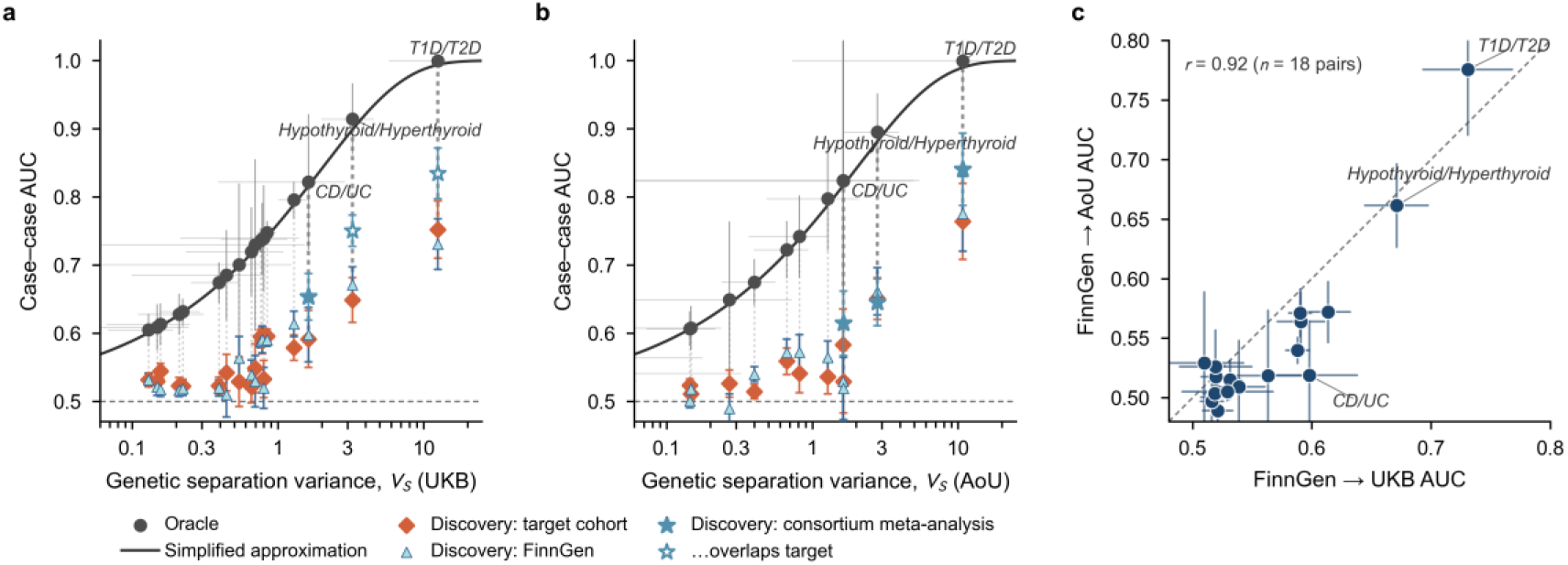
Case–case AUC of internally, externally and consortium-trained PRSs relative to the oracle ceiling. **a,b,** Case–case AUC plotted against genetic separation variance *V*_S_ (log scale), with UKB (**a**) and AoU (**b**) as target cohorts. The solid curve shows the simplified approximation, which lies below the analytical oracle AUC over the range shown. Dark points and error bars show target-specific oracle AUC estimates and ±1.96 block-jackknife s.e. obtained via 200 genomic blocks. Dotted lines connect each oracle estimate to the observed AUC achieved in the same target cohort by a PRS trained internally (diamonds), in FinnGen (triangles), or using published consortium GWAS summary statistics (stars). Error bars on PRS-based AUCs are ±1.96 s.e. from a delete-a-block jackknife over test individuals (Methods); 18 (**a**) and 14 (**b**) subtype pairs for internally trained and FinnGen-trained scores, **b** being restricted to pairs with a reliable All of Us oracle AUC, and 3 pairs for consortium-trained scores in each target cohort. Open stars denote the two consortium-trained scores in UKB whose discovery GWAS included UKB participants (type 1 versus type 2 diabetes and hypothyroidism versus hyperthyroidism); filled stars denote the comparisons without sample overlap, on which the reported gains are based. **c,** AUC of FinnGen-trained scores in AoU plotted against the corresponding AUC in UKB for the 18 subtype pairs, with ±1.96 jackknife s.e. on both axes. The dashed line indicates identity.

FinnGen had larger effective discovery sample sizes than the target-cohort training sets for all 18 subtype pairs in both cohorts: the FinnGen-to-target ratio, averaged geometrically over the two subtypes of each pair, had a median of 2.5 in UK Biobank (range, 1.1–9.6) and 3.8 in All of Us (range, 1.4–8.3). However, PRSs constructed using FinnGen GWAS summary statistics (FinnGen-trained PRSs) did not improve case–case AUC on average: the mean difference from internally trained PRSs was −0.002 in UK Biobank (range, −0.032 to +0.035) and +0.007 in All of Us (−0.053 to +0.079), and neither differed significantly from zero (paired *t*-test, *P* = 0.73 and 0.31, respectively). Individual pairs differed in both directions, with two significant differences in UK Biobank and three in All of Us after correction for the 18 comparisons within each cohort (*q* < 0.05; **Supplementary Fig. 16** and **Supplementary Table 8**), and the external-minus-internal difference was not associated with the discovery sample-size ratio in either target cohort (*r* = 0.11 and 0.09 for the log-transformed ratio; **Supplementary Fig. 17**). Imperfect transfer was also evident in the discovery GWAS themselves: the genetic correlations of the subtypes between FinnGen and the target cohort were generally below 1 (median 0.83 in UK Biobank and 0.76 in All of Us), significantly for 24 of 36 and 22 of 34 subtype comparisons (one-sided Wald test, Benjamini–Hochberg *q* < 0.05; **Supplementary Table 8**). Thus, the advantage of larger external GWAS may be offset by reduced transferability arising from differences in LD, allele frequencies and causal effect sizes^35,36^, and pairs with larger sample-size advantages did not show greater improvements in case–case AUC.

FinnGen-trained PRSs showed highly consistent performance across UK Biobank and All of Us. Observed case–case AUCs from FinnGen-trained PRSs were highly correlated between UK Biobank and All of Us across all 18 subtype pairs (*r* = 0.92), although they were on average 0.020 lower in All of Us (paired *t*-test, *P* = 0.007), with lower values in 15 of 18 pairs (**Fig. 5c**).

Consortium-scale discovery improved case–case AUC for the three subtype pairs with the highest oracle AUCs in UK Biobank (**Fig. 5a,b**). After excluding comparisons with evidence of discovery–target sample overlap, consortium-trained PRSs increased observed case–case AUC in three of the four comparisons and left it unchanged in the fourth (mean gain, 0.055; *q* values corrected across these four comparisons): from 0.529 with internally trained PRSs to 0.614 for Crohn disease versus ulcerative colitis in All of Us (*P* = 0.007, *q* = 0.009), from 0.764 to 0.840 for type 1 versus type 2 diabetes in All of Us (*P* = 0.005, *q* = 0.009), from 0.591 to 0.653 for Crohn disease versus ulcerative colitis in UK Biobank (*P* = 0.005, *q* = 0.009), and from 0.651 to 0.645 for hypothyroidism versus hyperthyroidism in All of Us (*P* = 0.78, *q* = 0.78).

These subtype pairs therefore remain prediction-limited: for several of these comparisons, larger discovery GWAS improved observed case–case AUC, but even consortium-scale data left much of the genetic ceiling unrealized.

## Discussion

We developed GenSep to estimate the maximum case–case discrimination attainable from additive genetic variation and quantify the proportion of the underlying genetic separation recovered by current PRSs. Simulations confirmed close agreement between analytical and empirical estimates of oracle AUC and showed that genetic architecture and PRS accuracy jointly determine subtype discrimination. Across 18 subtype pairs in UK Biobank, 11 had oracle AUCs above 0.70, including three above 0.80. By contrast, internally trained PRSs achieved a median AUC recovery of only 26.8% (range, 9.7–50.4%) and a median observed AUC of 0.543 (range, 0.521– 0.752). For the three pairs with oracle AUCs above 0.80, reaching a case–case AUC of 0.80 was projected to require approximately 940 type 1 diabetes cases (with type 2 diabetes at its current size), 36,000 hyperthyroidism cases (with hypothyroidism at its current size), or 98,000 Crohn disease and 207,000 ulcerative colitis cases in the training sample. Oracle AUCs were generally consistent among UK Biobank, All of Us and FinnGen and showed no significant ancestry-related differences within the Million Veteran Program, whereas differences in phenotype definition weakened the agreement between the Million Veteran Program and the other cohorts. FinnGen-trained PRSs, despite median 2.5- to 3.8-fold larger discovery samples, performed no better than internally trained PRSs, and substantially larger consortium GWAS raised case–case AUC by 0.062–0.086 in three of the four overlap-free comparisons and left it unchanged in the fourth.

These findings have three practical implications. First, the oracle AUC provides an interpretable measure of the maximum case–case discrimination achievable from additive genetic variation under the model, allowing direct comparisons across disease subtype pairs. Second, the oracle AUC helps assess whether clinically defined subtype distinctions correspond to differences in genetic liability. A high and reproducible ceiling supports a genetic basis for the clinical distinction, whereas a low ceiling suggests that the distinction is only weakly reflected in common-variant genetic architecture^13,14^. Cross-cohort consistency can strengthen this interpretation, whereas discrepancies may indicate sensitivity to phenotype definition or ascertainment. Third, comparing oracle and observed AUCs can guide efforts to improve subtype prediction for precision medicine. A high oracle AUC with low PRS recovery motivates larger discovery studies or improved genetic predictors, whereas a low oracle AUC points to greater reliance on clinical, molecular or environmental information.

Two previous studies are particularly relevant to GenSep. Wray et al. (2010)^26^ showed that the maximum case–control AUC attainable by a perfect genetic predictor depends on disease prevalence and liability-scale heritability, and related observed AUC to the proportion of genetic variance captured by the predictor. More recently, GDIS used subtype-specific heritabilities and genetic correlations to infer and visualize the genetic distance between subtype case groups^22^. GenSep extends both by translating subtype genetic architecture into an oracle case–case AUC derived from the prevalence and SNP heritability of each subtype and the genetic correlation between their liabilities, while quantifying the proportion of genetic separation recovered by individual PRSs (**Supplementary Note 1**).

The limited gains from larger external discovery GWAS are consistent with our framework: case–case AUC improves with subtype-specific PRS accuracy, but the extent of improvement depends on how well discovery effects transfer to the target cohort. Our sample-size projections suggest that FinnGen’s median 2.5- to 3.8-fold larger discovery samples would have substantially increased AUC if discovery effects had transferred perfectly to the target cohort. The below-unity genetic correlations between FinnGen and the target cohorts suggest that differences in linkage disequilibrium, allele frequencies, causal effect sizes and phenotype definition offset the benefits of larger discovery samples^35,36^. Larger consortium GWAS improved AUC for disease subtype pairs with high genetic ceilings, but substantial gaps to the oracle AUC remained. Approaching these ceilings will therefore require both larger discovery samples and closer alignment with the target population in ancestry and phenotype definitions.

Both the genetic ceiling and PRS recovery depend on how subtypes are defined and recorded. Misclassification can mask genetic differences between subtypes, inflate their estimated genetic correlation and reduce the estimated oracle AUC^37^. Consistent with this possibility, the oracle AUC for type 1 versus type 2 diabetes was approximately 1.00 under the strict UK Biobank definition but 0.605 under the broader MVP phecode definition. Misclassification in the evaluation set can also attenuate observed discrimination. If each recorded subtype group contains a fraction *ε* of cases from the other subtype, and misclassification is independent of the score within each true subtype, the observed AUC is given by AUC_obs_ = (1 − 2*ε*)AUC_true_ + *ε* (**Supplementary Note 3**). Thus, 5% contamination in each group reduces a true AUC of 0.80 to an observed AUC of 0.77. More accurate phenotyping may therefore improve both estimated genetic separability and observed discrimination. Where genetic ceilings remain low across cohorts with well-defined phenotypes, this may motivate the exploration of genetically informed subtypes.

Our study has several limitations. First, GenSep assumes a liability-threshold model with additive genetic effects and does not capture non-additive effects, gene– environment interactions or non-genetic determinants of subtype differences. Second, the oracle AUC represents a model-based genetic benchmark rather than an upper bound on classification using clinical, molecular or environmental information, and a high value does not by itself establish clinical utility. Third, cross-ancestry evaluation was limited by the number and size of eligible subtype pairs, restricting our ability to assess the generalizability of genetic ceilings and predictor recovery across populations. Further evaluation in larger and more diverse cohorts is therefore needed. Despite these limitations, GenSep provides a general statistical framework for distinguishing intrinsic genetic separability from incomplete predictor recovery, thereby identifying disease subtype pairs with the greatest potential for improved genetic prediction.

## Methods

### GenSep framework

We consider two clinically defined subtypes of a disease, indexed *i* ∈ {1,2}, each with a standardized liability *L_i_* = *G_i_* + *E_i_*, where *G_i_* is the additive genetic value captured by common variants and *E_i_* is the residual, with Var(*L_i_*) = 1 and 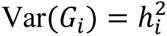, the liability-scale SNP heritability of subtype *i*. We define *G_i_* = *h_i_g_i_* and 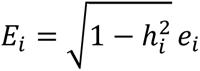, where *g_i_* and *e_i_* are standardized genetic and residual values with unit variance; the genetic values have correlation Cor(*g*_1_, *g*_2_) = *r*_g_, the residuals have correlation *r*_e_, and genetic and residual components are independent (Cov(*g_i_*, *e_j_*) = 0 for all *i* and *j*). An individual is a case of subtype *i* when *L_i_* > *t_i_*. The threshold is *t_i_* = Φ^−1^(1 − *K_i_*), where *K_i_* is the population prevalence and Φ is the standard normal cumulative distribution function. Conditioning on case status changes the distribution of the genetic values. The mean of *g_i_* among subtype-*i* cases is *λ_i_h_i_* rather than 0, where *λ_i_* = *φ*(*t_i_*)/*K_i_* is the selection intensity and *φ* is the standard normal density function. The variance of any variable correlated with *L_i_* is also reduced, by an amount governed by *δ_i_* = *λ_i_*(*λ_i_* − *t_i_*) (**Supplementary Note 1**).

Given the true genetic values, the optimal linear discriminant between the two case groups is *D* = *λ*_1_*h*_1_*g*_1_ − *λ*_2_*h*_2_*g*_2_ = *λ*_1_*G*_1_ − *λ*_2_*G*_2_, so that the weights on the standardized genetic values are *w_i_* = *λ_i_h_i_* and the weights on the raw genetic values are the selection intensities alone (**Supplementary Note 1**). Under this discriminant the difference between the mean of *D* in subtype-1 and in subtype-2 cases equals its marginal variance, which defines the oracle genetic separation variance

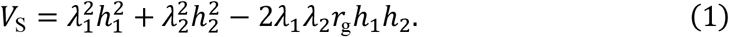

Approximating the distributions of *D* within each case group by normal distributions with their conditional means and variances gives the analytical oracle case–case AUC,

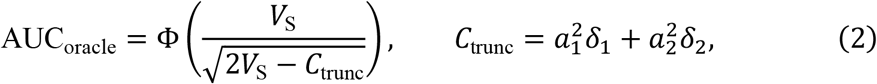

with 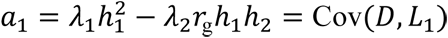 and 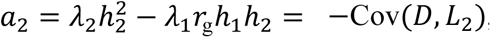, so that *C*_trunc_ is the reduction in the sum of the two within-group variances of *D* that results from selecting cases as the individuals above the liability threshold (**Supplementary Note 1**). AUC_oracle_ is the case–case discrimination attainable under the liability-threshold model when both subtypes’ true additive genetic values are observed without error. Dropping *C*_trunc_, which is non-negative and therefore makes the approximation slightly conservative, gives the simplified approximation

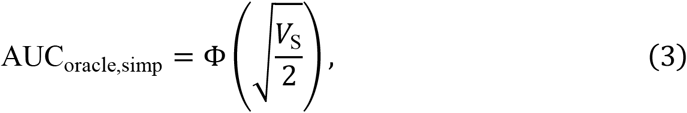

which makes the dependence on *V*_S_ explicit. Under this approximation the oracle AUC depends on the genetic architecture only through *V*_S_, so the effect of any change in heritability, prevalence or genetic correlation on the ceiling is determined entirely by its effect on *V*_S_ in equation (1). For the same genetic separation variance, equation (3) is never larger than equation (2), because the truncation term is non-negative. Equations (2) and (3) do not involve *r*_e_, so the oracle AUC is unaffected by the residual correlation (**Supplementary Note 1**).

Real predictors estimate *g_i_* with error. Writing the standardized PRS for subtype *i* as *ĝ_i_*, with accuracy 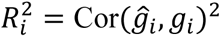, the two scores are combined into 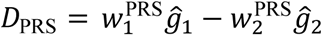. Here 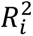 is the squared correlation between the PRS and the subtype’s true standardized genetic value; it is evaluated directly in simulations and estimated indirectly from case–control predictive performance in real data (**Supplementary Note 3**). The optimal weights are derived in the same way as for the true genetic values, but they depend on the difference in each score between the two case groups, which is attenuated by *R_i_*. They become *λ_i_h_i_* when both scores are perfect (**Supplementary Note 2**). *V*_PRS_, the PRS counterpart of *V*_S_, quantifies the separation between subtype-1 and subtype-2 cases achieved by the combined score, and it gives the model-predicted PRS case–case AUC in the same way that *V*_S_ gives the oracle AUC in equations (2) and (3) (**Supplementary Note 2**). We compare the PRS-based score with the genetic ceiling by its separation recovery, *f*, the fraction of the genetic separation variance captured by the score, and its AUC recovery, *f*_AUC_, the fraction of the above-chance oracle AUC that the score achieves:

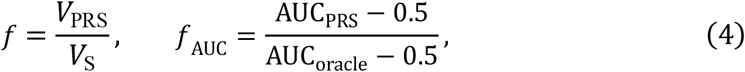

where AUC_PRS_ is the PRS case–case AUC. For oracle AUCs below about 0.8, *f*_AUC_ is close to 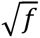 (**Supplementary Note 3** and **Supplementary Fig. 18**). In simulation, the true parameters are known, so *V*_PRS_ is computed exactly from them. Separately, in the real-data analysis we estimate it from the observed AUC by inverting equation (3), *V*_PRS_ = 2[Φ^−1^(AUC_PRS_)]^2^. Full derivations are given in **Supplementary Notes 1** and **2**.

The framework is implemented in GenSep, an open-source C++ program (https://chaoning.github.io/gensep/). Standard errors were estimated using a block jackknife with 200 contiguous genome blocks, as in LDAK SumHer^32,38^. In each leave-one-block-out replicate, 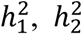 and *r*_g_ were re-estimated and all derived quantities were recalculated, so that for any quantity *θ*

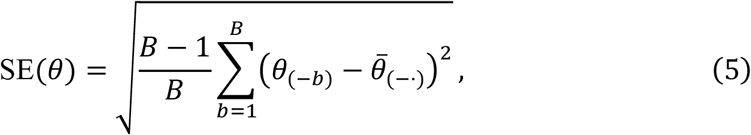

where *B* = 200, *θ*_(−*b*)_ is the estimate obtained with block *b* removed and *θ̅*_(−⋅)_ is the mean of the *B* replicate estimates (**Supplementary Note 3**).

### Simulations

We validated the oracle expressions on a full factorial grid of 1,728 settings (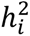 ∈ {0.1,0.3,0.6}; *K_i_* ∈ {0.001,0.01,0.1,0.3}; *r*_g_ ∈ {0,0.1,0.3,0.6}; *r*_e_ ∈ {0,0.3,0.6}), each replicated 30 times. Unless stated otherwise, results use the 576 settings with *r*_e_ = 0; the other residual-correlation levels are used only to test the effect of the residual correlation. In each replicate we drew bivariate normal genetic values and residuals with the specified correlations, applied the two thresholds and sampled 5,000 cases per subtype. The empirical oracle AUC was computed from the discriminant *D* = *w*_1_*g*_1_ − *w*_2_*g*_2_ formed from the true genetic values, in two ways. First, we fixed the weight at its analytical value *w_i_* = *λ_i_h_i_*, which tests equations (2) and (3) given the weight. Second, we fitted the weight in a random half of the cases and evaluated the AUC in the held-out half, which tests whether data-driven optimization recovers the analytical weight. To test the prediction that the oracle AUC does not depend on the residual correlation (**Supplementary Fig. 4**), we compared settings that differed only in *r*_e_. The PRS expressions were validated on a second grid in which each subtype’s score accuracy was additionally varied over 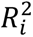 ∈ {0.1,0.3,0.5,0.7,0.9,1.0}. To represent PRSs with controlled prediction accuracy, scores were generated as 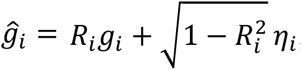, where *η_i_* is independent standard normal noise and the hat denotes a predictor of the genetic value. This simulation generated genetic values and residuals directly, without simulating or using individual-level genotypes. The empirical case–case AUC was computed between subtype-1 and subtype-2 cases from the combined score at the optimal weights (**Supplementary Note 2**), and compared with the model-predicted AUC computed from the setting’s parameters alone (**Supplementary Note 2**). To test whether the optimal weights can be recovered from data, the weights were also fitted by a grid search in a random half of the cases, the AUC was evaluated in the other half, and the fitted weights were compared with the closed-form weights. This grid comprised 20,736 settings (every combination of heritability, prevalence and score accuracy for each subtype, with genetic correlation 0, 0.1, 0.3 or 0.6 and no residual correlation), each replicated 30 times with 5,000 cases per subtype.

To confirm that the oracle quantities can be recovered from GWAS summary statistics rather than from known parameters, we simulated phenotypes on the UK Biobank genotypes: 324 parameter settings (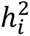 ∈ {0.1,0.3,0.6}; *K_i_* ∈ {0.01,0.1,0.3}; *r*_g_ ∈ {0,0.1,0.3,0.6}), each replicated 10 times, with 1,000 shared causal variants per replicate and correlated effect sizes drawn to the specified heritabilities and genetic correlation. In each simulated data set, a GWAS of each subtype was run in the training set with a linear model; 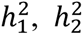 and *r*_g_ were estimated from the resulting summary statistics with SumHer^32^ and substituted into equations (1)–(3); and PRSs were built from the same summary statistics with MegaPRS^34^ and evaluated in the held-out test set to give the observed case–case AUC and the recovery of equation (4). The observed AUC was evaluated by two-fold cross-fitting within the test set. The test individuals were split once into two halves; in one half, PRS accuracy was estimated and the optimal weights (**Supplementary Note 2**) were calculated from it together with the true heritabilities, genetic correlation and prevalences of the simulated setting, and the resulting score was evaluated in the other half. The halves were then exchanged, and the two AUCs were averaged with weights proportional to the number of case pairs. Thus, no weight was derived from the individuals in whom it was evaluated. Because the true genetic values are retained, the estimated *V*_S_ and oracle AUC could be compared with the values implied by the simulation parameters, and each score’s accuracy 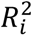 was both computed directly, as the squared correlation with the true genetic value, and inferred from the score’s case–control AUC as in the real data; the two routes were compared (**Supplementary Fig. 8**). Control-subsampling sensitivity analyses at case:control ratios of 1:1 and 1:4 used a subset of 45 parameter settings (**Supplementary Figs. 5 and 8**).

### Cohorts and genetic data

#### UK Biobank

UK Biobank is a population cohort of approximately 500,000 UK residents aged 40–69 years at recruitment^28^. All primary analyses were performed in UK Biobank: the 18 subtype pairs were defined here, and their oracle AUCs, PRS recovery and sample-size projections were estimated in it; FinnGen-trained and consortium-trained scores were also evaluated here, and its genotypes were used for the simulations. We used the directly genotyped array data, restricted to the European genetically inferred ancestry group (Data-Field 30079) with one member of each pair related at third degree or closer (KING kinship > 0.0442) removed, and retained autosomal variants with missingness below 5%, minor allele frequency above 1% and Hardy–Weinberg equilibrium *P* > 10^−8^; the analysis set comprises 357,373 unrelated individuals and 561,997 variants.

#### All of Us

All of Us is a US population cohort with genotype data linked to electronic health records^29^. It served as an independent cohort for replication, processed with the same pipeline, in which oracle AUCs and internally trained scores were estimated for the 18 pairs; FinnGen-trained and consortium-trained scores were also evaluated here. We used the Controlled Tier v9 curated data repository and its genotyped array data, restricted to the 229,152 participants with a European ancestry probability above 0.95, and retained autosomal variants with missingness below 5%, minor allele frequency above 1% and Hardy–Weinberg equilibrium *P* > 10^−8^, leaving 737,766 variants; the 224,334 participants with complete covariate data formed the analysis set.

#### FinnGen

FinnGen combines genotypes with national health registries for approximately 500,000 Finnish participants. It provided oracle AUCs for all 18 pairs and served as the external discovery GWAS for scores evaluated in UK Biobank and All of Us. We used only summary statistics from FinnGen data freeze 13^30^, a Finnish founder population with no sample overlap with UK Biobank and All of Us.

#### Million Veteran Program

The Million Veteran Program is a US cohort of military veterans with genotype data linked to health records. It provided oracle AUCs in three ancestry groups for the cross-cohort and cross-ancestry comparisons. We used the openly released per-phecode summary statistics^31^ (dbGaP accession phs002453) for the European, African and Admixed American ancestry groups.

#### Published consortium GWAS

For the three pairs with the highest oracle AUCs in UK Biobank (type 1 versus type 2 diabetes, hypothyroidism versus hyperthyroidism, and Crohn disease versus ulcerative colitis), published consortium meta-analyses provided discovery GWAS substantially larger than either biobank’s training set, and scores trained on them were evaluated in UK Biobank and All of Us. The discovery GWAS were the type 1 diabetes meta-analysis of McGrail et al. 2026^39^ (GCST90824163; 20,355 cases and 797,363 controls) and the T2DGGI European type 2 diabetes meta-analysis of Suzuki et al. 2024^40^ (GCST90492734; 242,283 cases and 1,569,734 controls); the Crohn disease and ulcerative colitis meta-analyses of Liu et al. 2023^41^ (GCST90446792, 20,873 cases and 346,719 controls; GCST90446794, 23,252 cases and 352,256 controls); and the hypothyroidism and hyperthyroidism meta-analyses of Figuerêdo et al. 2024^42^ (GCST90319320, 58,783 cases and 633,203 controls; GCST90319319, 6,442 cases and 736,566 controls). Sample overlap between each external GWAS and UK Biobank or All of Us was assessed with the cross-trait intercept of the bivariate SumHer regression^32^ between the external GWAS and the target cohort’s own GWAS of the same trait, which is expected to depart from zero when the two GWAS share samples. The intercepts indicated overlap with UK Biobank for the diabetes GWAS (0.153 for type 1 and 0.202 for type 2 diabetes) and the thyroid GWAS (0.335 for hypothyroidism and 0.051 for hyperthyroidism) but not for the inflammatory bowel disease GWAS (0.003 for Crohn disease and 0.026 for ulcerative colitis), and none of the external GWAS lists All of Us among its contributing cohorts. Consortium-trained scores were evaluated in both cohorts; the UK Biobank comparisons for type 1 versus type 2 diabetes and for hypothyroidism versus hyperthyroidism, which showed overlap, are shown in **Fig. 5** but excluded from the summary of transfer gains. Cohort details, endpoint and phecode mappings and the external GWAS are described in **Supplementary Note 3** and **Supplementary Table 1**.

### Subtype-pair definitions and prevalences

We analyzed 18 pairs of clinically defined subtypes in UK Biobank, selected by defined clinical criteria and by the quality of the UK Biobank genetic-parameter estimates (**Supplementary Note 3**). In each pair, the two subtypes were either subtypes of a single disease entity, distinguished by etiology or pathology, anatomical site, disease course or cell lineage, or related conditions of the same organ system that are classified as separate entities. Phenotypes were derived from ICD-10 diagnoses in the hospital inpatient records. The framework defines case status separately for each subtype, so an individual may be a case of both, and the phenotypes follow this definition. For four pairs (type 1 versus type 2 diabetes, Crohn disease versus ulcerative colitis, ischemic versus hemorrhagic stroke and hypothyroidism versus hyperthyroidism), we imposed mutually exclusive case definitions and excluded individuals carrying codes for both members of the pair, because dual coding in these pairs is more likely to reflect misclassification or diagnostic revision than true co-occurrence. For the remaining 14 pairs, individuals meeting both subtype definitions were retained in both subtype phenotypes and appear in both arms of the case–case evaluation. The ICD-10 codes defining each subtype and the control definitions are given for each pair in **Supplementary Table 1** and **Supplementary Note 3**.

In All of Us, the same definitions were applied to ICD-10 (ICD-10-CM) diagnoses in the electronic health records. FinnGen and MVP provide summary statistics for predefined phenotypes only, so each subtype was matched to the FinnGen endpoint or MVP phecode that most closely corresponded to its UK Biobank definition; 15 of the 18 pairs could be matched in MVP, the remaining three being indistinguishable in the phecode system (**Supplementary Table 1**). Population prevalences (*K*) were reference values chosen from published UK and European epidemiological estimates (**Supplementary Table 1**) and were held fixed across cohorts and ancestries. We assessed sensitivity to the assumed prevalences by halving or doubling the prevalence of either or both subtypes and re-estimating the oracle AUC; the estimates were not materially affected (**Supplementary Note 3** and **Supplementary Fig. 10**).

### Estimation of the genetic ceiling

Within UK Biobank and All of Us, we ran the GWAS of each subtype against controls using a linear model adjusted for sex, age, age^2^, age^3^, sex × age, sex × age^2^, sex × age^3^ and the first 20 (UK Biobank) or 10 (All of Us) genetic principal components. Observed-scale SNP heritabilities and the cross-subtype genetic correlation were estimated with SumHer^32^ from the two marginal GWAS. SumHer tagging files were computed with LDAK from UK Biobank and All of Us participants and, for MVP, from the ancestry-matched 1000 Genomes reference panels^43^; for FinnGen, which releases no individual-level genotypes, a tagging file was derived from the public FinnGen LD matrices (**Supplementary Note 3**). Heritabilities were converted to the liability scale using the population prevalence and the cohort case fraction^44^, and the liability-scale heritabilities, genetic correlation and prevalences were substituted into equations (1)–(3). The same estimation framework was applied in every cohort and ancestry, using cohort-specific summary statistics and case fractions and ancestry-matched LD references.

The estimates of a subtype pair in a given cohort were used only if both heritabilities were positive, the genetic correlation was finite and not at the ±0.999 estimator boundary, and all derived quantities existed; cross-cohort correlations and difference tests were restricted to pairs meeting this rule in both analyses (**Supplementary Table 2**). Re-estimating the oracle AUC of type 1 versus type 2 diabetes under seven cohort–definition combinations, from UK Biobank, All of Us, FinnGen and MVP, showed that the ceiling varies with the definition: the oracle AUC was 1.00 under the strict definition (an E10 code without E11 or E12–E14 codes, in UK Biobank and in All of Us) and 0.60 under the MVP phecode definition (250.1), as the genetic correlation between the two arms rose from near zero to 0.95. Within each of UK Biobank and All of Us, broadening the definitions to any ICD-10 code starting with E10 and any code starting with E11 lowered the oracle AUC from 1 to 0.87 and 0.89 (**Supplementary Note 3**).

### Polygenic scores and observed case–case discrimination

Polygenic scores were trained on discovery GWAS from the target cohort itself (internal) or from FinnGen and published consortium GWAS summary statistics (external). Individuals in each target cohort, UK Biobank or All of Us, were partitioned into a training set (80%) and a held-out test set (20%; *n* = 71,475 in UK Biobank and 44,867 in All of Us). Internal scores were trained on GWAS of each subtype in the training set; external scores were trained on FinnGen and consortium GWAS as described below. Subtype-specific scores were constructed with MegaPRS^34^ from each discovery GWAS, using default parameters. The accuracy of each score, 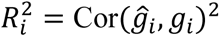, required by the PRS case–case AUC expressions, was inferred from its case–control AUC against its own marginal phenotype in the test set (**Supplementary Note 3**). This is a phenotype-based proxy: in simulation 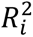 is computed directly from the true genetic values, whereas in real data it must be inferred from predictive performance. External scores were built from the FinnGen and consortium summary statistics after aligning them to the target cohort’s variants with MegaPRS. FinnGen overlaps neither target cohort, so its scores provide a test of transfer free of discovery–target sample overlap; consortium comparisons with evidence of discovery–target overlap are shown but excluded from the summary of transfer gains.

The observed case–case score is 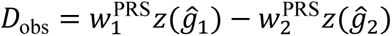, where *z*(⋅) standardizes each score within the test set. Here, 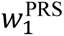 and 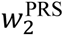 are the optimal weights (**Supplementary Note 2**), which are computed from each pair’s estimated heritabilities, genetic correlation, prevalences and PRS accuracies. To avoid estimating the PRS accuracies in the individuals in whom the weights are evaluated, the test individuals were split once into two halves. In one half, PRS accuracy was estimated (**Supplementary Note 3**) and the optimal weights (**Supplementary Note 2**) were calculated from it together with the estimated heritabilities, genetic correlation and prevalences, and the resulting score was evaluated in the other half. The halves were then exchanged, and the two AUCs were averaged with weights proportional to the number of case pairs (**Supplementary Note 3**). Thus, the PRS accuracies and score orientations were never estimated in the individuals in whom the score was evaluated, whereas the heritabilities and genetic correlation entering the weights are the full-cohort estimates of the genetic-ceiling analysis, which include the test individuals, and were held fixed, so that cross-fitting isolates the estimation of the accuracies and weights but not that of the genetic parameters. The resulting case-pair-weighted average of the two evaluation-half AUCs is AUC_PRS_. Observed AUCs were compared with the values predicted by the analytical PRS AUC expression (**Supplementary Note 2**) from each subtype pair’s estimated heritabilities, genetic correlation, prevalences and score accuracies. A sensitivity weighting was evaluated on the same halves: the weight direction that maximized the case–case AUC in the weight half, found by a grid search in 1° steps (**Supplementary Table 3**). Internally trained, FinnGen-trained and consortium-trained scores were all evaluated in the same way, in the same test individuals with the same case definitions and the same weighting rule, so that they can be compared directly (**Supplementary Note 3**).

### Sample-size projection

We projected the case–case AUC against the number of training cases per subtype. The projection rests on the standard relationship between polygenic-score accuracy and training sample size^25,45,46^: under the infinitesimal model, for the squared correlation *R*^2^ between a PRS and the true genetic liability, 1/*R*^2^ − 1 is proportional to 1/*N*_eff_, where *N*_eff_ = 4*N*_case_*N*_ctrl_/(*N*_case_ + *N*_ctrl_) is the effective discovery sample size. We applied this relationship to each subtype’s score separately. Let 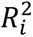 denote the accuracy of the score for subtype *i* on the genetic scale (**Supplementary Note 3**) and *κ_i_* the factor by which the effective sample size of arm *i* is multiplied; then

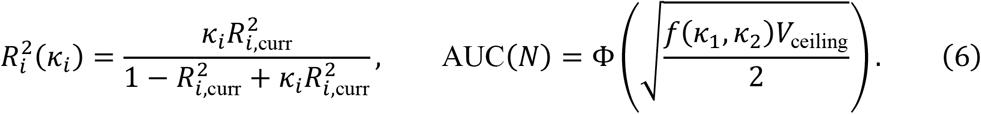

The two extrapolated accuracies were recombined through the simplified-approximation expressions for the observed case–case score (**Supplementary Note 2**), giving the separation variance *V*_PRS_(*κ*_1_, *κ*_2_) of the optimally weighted score and the recovered fraction *f*(*κ*_1_, *κ*_2_) = *V*_PRS_(*κ*_1_, *κ*_2_)/*V*_S_, which rises to 1 as both *κ_i_* grow; with *V*_ceiling_ = 2[Φ^−1^(AUC_oracle_)]^2^, every projected curve therefore asymptotes at the oracle AUC reported for that pair. The current accuracies 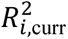 were obtained from the case–control AUC of each subtype’s score (**Supplementary Note 3**). The main projection scales both arms by the same factor and assumes an unlimited number of controls. We also projected the cases required when only one subtype’s training set grows and the other is held at its current size (**Supplementary Table 4**). The number of cases required to reach a target AUC was obtained by solving *f*(*κ*_1_, *κ*_2_) = *f*_target_ numerically. Because the projection holds the genetic architecture fixed and is calibrated to one observed AUC per pair, the required sample sizes should be interpreted as order-of-magnitude estimates rather than as the output of a power calculation (**Supplementary Note 3**).

## Statistical analysis

All tests are two-sided unless stated otherwise, and multiple-testing correction is by the Benjamini–Hochberg procedure^47^ within an explicitly stated family. Cross-cohort agreement was summarized by Pearson and Spearman correlations over the pairs reliable in both analyses (**Supplementary Note 3**), computed with and without type 1 versus type 2 diabetes, which is a high-leverage point in every panel; per-pair differences were tested by 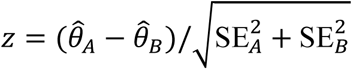, because the cohorts are independent samples, with correction within each combination of comparison and quantity. The external-minus-internal difference in case–case AUC was related to the log ratio of the FinnGen and internal discovery effective sample sizes by Pearson correlation. The genetic correlation of each subtype between its FinnGen endpoint GWAS and the target-cohort GWAS was estimated with SumHer, using the target-cohort tagging file and estimating the sample-overlap term, and was tested for being below 1 with a one-sided Wald test, with correction within each target cohort.

Standard errors on the oracle quantities are the 200-block genome jackknife described above. Standard errors on the observed case–case AUCs are a delete-a-block jackknife over the unique case participants of the test set, with *B* = min(200,size of the smaller arm) blocks: each participant was assigned to one block, deletion removed all case memberships of that participant, and the PRS accuracies and combination weights were refitted within each deletion, while the heritabilities, genetic correlation and discovery effect sizes were held fixed; this covers sampling of the individuals in whom the score was evaluated, not the estimation of the discovery effect sizes. Average differences across pairs were tested with a paired *t*-test.

### Software

Analyses used GenSep v1.0.0 (https://chaoning.github.io/gensep/), LDAK v6 (SumHer and MegaPRS), PLINK v1.9 and v2.0, and Python v3.13.

## Data availability

UK Biobank data are available to approved researchers through the UK Biobank Access Management System (https://www.ukbiobank.ac.uk); this work used data obtained under application 21432. UK Biobank has ethics approval from the North West Multi-centre Research Ethics Committee (REC reference 21/NW/0157), and all participants provided written informed consent. All of Us data are available to registered researchers through the Researcher Workbench (https://www.researchallofus.org); this work used the Controlled Tier v9 curated data repository. FinnGen DF13 summary statistics are publicly available at https://www.finngen.fi/en/access_results. Million Veteran Program summary statistics are available from dbGaP under accession phs002453. The consortium GWAS summary statistics are available from the GWAS Catalog under accessions GCST90824163, GCST90492734, GCST90446792, GCST90446794, GCST90319320 and GCST90319319.

## Code availability

GenSep is open-source software released under GPL-3.0, with documentation and source code at https://chaoning.github.io/gensep/.

## Acknowledgements

This research has been conducted using the UK Biobank Resource under Application Number 21432. We gratefully acknowledge All of Us participants for their contributions, without whom this research would not have been possible, and we thank the National Institutes of Health’s All of Us Research Program for making available the participant data examined in this study. We also acknowledge the participants and investigators of the FinnGen study and the Million Veteran Program. The computing for this project was performed on the GenomeDK cluster (Aarhus University) and the All of Us Researcher Workbench.

## Author contributions

C.N. and D.S. conceived and designed the study. C.N. developed the statistical framework, implemented the GenSep software and performed all simulations and real-data analyses, with supervision from D.S. J.H. contributed to the interpretation of the results. C.N. wrote the manuscript; J.H. and D.S. critically revised it. All authors reviewed and approved the final version of the manuscript.

