## Supplementary Information for "Quantifying the genetic separability of disease subtypes"

**Contents**

Supplementary Note 1: Case–Case AUC and Optimal Genetic Discriminant under the Liability-Threshold Model

Supplementary Note 2: PRS-Based Case–Case AUC and Optimal Genetic Discriminant under the Liability-Threshold Model

Supplementary Note 3: Detailed Methods

Supplementary Figs. 1–18

Supplementary Tables 1–8 (titles and legends; the tables themselves are provided as a separate Excel workbook)

Supplementary References

### Supplementary Notes

#### Supplementary Note 1: Case–Case AUC and Optimal Genetic Discriminant under the Liability-Threshold Model

##### 1.1 Overview

We derive the theoretical case–case discrimination accuracy for separating two disease subtypes when true genetic values are observed. The derivation is based on a bivariate liability-threshold model[1] and provides a true-genetic-value oracle benchmark for subtype discrimination.

Our goal is to derive the optimal linear genetic discriminant for distinguishing subtype 1 cases from subtype 2 cases. We work on the standardized genetic-value scale,

$$G_i = h_i g_i, g_i \sim N(0,1),$$

where $h_i^2$ denotes the liability-scale SNP heritability of subtype $i$. Under this notation, the optimal discriminant will have weights that explicitly depend on both subtype prevalence and subtype heritability.

Under the leading-order approximation derived in Section 1.8, the optimal genetic discriminant is

$$D_\lambda = \lambda_1 h_1 g_1 - \lambda_2 h_2 g_2,$$

where

$$\lambda_i = \frac{\phi(t_i)}{K_i}$$

is the liability-threshold selection intensity for subtype $i$.

The corresponding genetic separation variance is

$$V_\mathrm{S} = \lambda_1^2 h_1^2 + \lambda_2^2 h_2^2 - 2\lambda_1\lambda_2 r_\mathrm{g} h_1 h_2.$$

The corresponding analytical oracle case–case AUC (main-text equation (2)) is

$$\mathrm{AUC}_\mathrm{cc}^\mathrm{TGV} \approx \Phi\left(\frac{V_\mathrm{S}}{\sqrt{2V_\mathrm{S} - C_\mathrm{trunc}}}\right), \qquad C_\mathrm{trunc} = a_1^2\delta_1 + a_2^2\delta_2,$$

where $a_1 = \lambda_1 h_1^2 - \lambda_2 r_\mathrm{g} h_1 h_2$, $a_2 = \lambda_2 h_2^2 - \lambda_1 r_\mathrm{g} h_1 h_2$, $\delta_i = \lambda_i(\lambda_i - t_i)$ and $t_i = \Phi^{-1}(1 - K_i)$ is the liability threshold of subtype $i$. The term $C_\mathrm{trunc} \geq 0$ is the reduction in the sum of the two within-group variances of $D_\lambda$ that results from selecting cases as the individuals above the liability threshold (Sections 1.11–1.12).

Dropping $C_{\text{trunc}}$ gives the simplified approximation of main-text equation (3) (Section 1.13),

$$\mathrm{AUC}_{\mathrm{cc}}^{\mathrm{TGV}} \approx \Phi\left(\sqrt{\frac{V_{\mathrm{S}}}{2}}\right).$$

This expression shows that $V_{\mathrm{S}}$ is the primary determinant of case–case genetic discrimination.

**1.2 Liability-Threshold Model on the Standardized Genetic Scale**

For subtype $i \in \{1,2\}$, assume the liability-threshold model

$$L_i = G_i + E_i, \qquad \mathrm{Var}(L_i) = 1.$$

We write the genetic component as

$$G_i = h_i g_i,$$

where

$$g_i \sim N(0,1),$$

Here, $h_i^2$ is the liability-scale SNP heritability of subtype $i$, and $G_i$ is the corresponding additive genetic value. Therefore,

$$\mathrm{Var}(G_i) = h_i^2.$$

Similarly, write the residual component as

$$E_i = \sqrt{1 - h_i^2} e_i,$$

where

$$e_i \sim N(0,1).$$

Thus, the liability can be written as

$$L_i = h_i g_i + \sqrt{1 - h_i^2} e_i.$$

The standardized genetic components $g_1$ and $g_2$ are allowed to be correlated:

$$\mathrm{Cor}(g_1, g_2) = r_{\mathrm{g}}.$$

Therefore,

$$\mathrm{Cov}(g_1, g_2) = r_\mathrm{g}.$$

The covariance between the unstandardized genetic components is

$$\mathrm{Cov}(G_1, G_2) = \mathrm{Cov}(h_1 g_1, h_2 g_2) = r_\mathrm{g} h_1 h_2.$$

The residual components may also be correlated. Let

$$\mathrm{Cor}(e_1, e_2) = r_\mathrm{e}.$$

Then

$$\mathrm{Cov}(E_1, E_2) = r_\mathrm{e}\sqrt{(1 - h_1^2)(1 - h_2^2)}.$$

We assume that $(g_1, g_2)$ and $(e_1, e_2)$ are jointly Gaussian vectors, with within-vector correlations $r_\mathrm{g}$ and $r_\mathrm{e}$, and are independent of each other, so that all four components are jointly Gaussian and genetic and residual components are independent across all subtype combinations:

$$\mathrm{Cov}(g_i, e_j) = 0, i, j \in \{1,2\}.$$

Therefore, the liability-scale covariance between the two subtypes is

$$\begin{aligned}\mathrm{Cov}(L_1, L_2) \quad &= \mathrm{Cov}\left(h_1 g_1 + \sqrt{1 - h_1^2} e_1, h_2 g_2 + \sqrt{1 - h_2^2} e_2\right) \\ &= r_\mathrm{g} h_1 h_2 + r_\mathrm{e}\sqrt{(1 - h_1^2)(1 - h_2^2)}.\end{aligned}$$

Since each liability has unit variance, the liability-scale correlation is

$$r_L = r_\mathrm{g} h_1 h_2 + r_\mathrm{e}\sqrt{(1 - h_1^2)(1 - h_2^2)}.$$

Each case group is defined by a single liability threshold (Section 1.3), so the conditional distributions of $g_1$ and $g_2$ within each case group do not depend on the residual correlation; $r_\mathrm{e}$ therefore does not enter the genetic discriminant or its case–case AUC, although it affects the overlap between the two case groups through the liability correlation.

We condition subtype $i$ cases on the single event

$$L_i > t_i.$$

Thus, subtype definitions are allowed to overlap in principle.

#### 1.3 Liability Thresholds and Selection Intensities

Subtype $i$ cases are defined by

$$L_i > t_i.$$

Let $K_i$ denote the population prevalence of subtype $i$. Since

$$L_i \sim N(0,1),$$

we have

$$K_i = P(L_i > t_i) = 1 - \Phi(t_i),$$

where $\Phi(\cdot)$ is the standard normal cumulative distribution function. Therefore,

$$t_i = \Phi^{-1}(1 - K_i).$$

Define the inverse Mills ratio, also called the selection intensity,

$$\lambda_i = \lambda(t_i) = \frac{\phi(t_i)}{1 - \Phi(t_i)} = \frac{\phi(t_i)}{K_i},$$

where $\phi(\cdot)$ is the standard normal density function.

For a standard normal variable $Z \sim N(0,1)$, the upper-tail truncated mean is given by the standard truncated-normal result

$$E(Z \mid Z > t_i) = \lambda_i.$$

We also define the upper-tail truncation variance-reduction factor

$$\delta_i = \delta(t_i) = \lambda_i(\lambda_i - t_i).$$

For a standard normal variable,

$$\mathrm{Var}(Z \mid Z > t_i) = 1 - \delta_i.$$

Thus, $\lambda_i$ controls the mean shift caused by case ascertainment, while $\delta_i$ controls the reduction in variance caused by liability truncation.

A rarer subtype has a larger threshold $t_i$ and a larger selection intensity $\lambda_i$. Therefore, rarer subtype cases are selected from a more extreme tail of the liability distribution.

#### 1.4 Conditional Mean and Variance under Liability-Threshold Truncation

Let $X$ and $Y$ be jointly normally distributed with $\mathrm{E}(X) = \mathrm{E}(Y) = 0$ and $\mathrm{Var}(Y) = 1$. Because $X$ and $Y$ are jointly Gaussian, $X$ can be decomposed as

$$X = \beta Y + \varepsilon, \qquad \beta = \frac{\mathrm{Cov}(X,Y)}{\mathrm{Var}(Y)} = \mathrm{Cov}(X,Y),$$

where $\varepsilon$ is independent of $Y$ with $\mathrm{E}(\varepsilon) = 0$ and $\mathrm{Var}(\varepsilon) = \mathrm{Var}(X) - \beta^2$. Conditioning on $Y > t$ therefore acts on $X$ only through $Y$, and the truncation moments of Section 1.3 give

$$\mathrm{E}(X \mid Y > t) = \beta E(Y \mid Y > t) = \mathrm{Cov}(X,Y)\lambda(t),$$

and

$$\begin{aligned}\mathrm{Var}(X \mid Y > t) = \beta^2 \mathrm{Var}(Y \mid Y > t) + \mathrm{Var}(\varepsilon) = \beta^2\{1 - \delta(t)\} + \mathrm{Var}(X) - \beta^2 \\ = \mathrm{Var}(X) - \mathrm{Cov}(X,Y)^2\delta(t).\end{aligned}$$

Thus, truncating $Y$ above threshold $t$ shifts the mean of any correlated variable $X$ by an amount proportional to $\mathrm{Cov}(X,Y)$ and reduces its variance by $\mathrm{Cov}(X,Y)^2\delta(t)$. These identities are exact for the first and second moments under single-threshold truncation.

**1.5 Conditional Means of Standardized Genetic Values**

We first compute the expected standardized genetic values among subtype cases.

For subtype $i$, cases satisfy

$$L_i > t_i.$$

Using the conditional mean identity under liability-threshold truncation,

$$E(g_i \mid L_i > t_i) = \mathrm{Cov}(g_i, L_i)\lambda_i.$$

Because

$$L_i = h_i g_i + \sqrt{1 - h_i^2} e_i,$$

and

$$\mathrm{Cov}(g_i, e_i) = 0,$$

we have

$$\mathrm{Cov}(g_i, L_i) = \mathrm{Cov}\left(g_i, h_i g_i + \sqrt{1 - h_i^2} e_i\right) = h_i.$$

Therefore,

$$E(g_i \mid L_i > t_i) = h_i \lambda_i.$$

Specifically,

$$E(g_1 \mid L_1 > t_1) = h_1\lambda_1,$$

and

$$E(g_2 \mid L_2 > t_2) = h_2\lambda_2.$$

We also need the cross-subtype expectations. For subtype 1 cases,

$$E(g_2 \mid L_1 > t_1) = \mathrm{Cov}(g_2, L_1)\lambda_1.$$

Since

$$L_1 = h_1 g_1 + \sqrt{1 - h_1^2} e_1,$$

we have

$$\mathrm{Cov}(g_2, L_1) = h_1 \mathrm{Cov}(g_2, g_1) + \sqrt{1 - h_1^2} \mathrm{Cov}(g_2, e_1).$$

By cross-component independence,

$$\mathrm{Cov}(g_2, e_1) = 0.$$

Since

$$\mathrm{Cov}(g_2, g_1) = r_{\mathrm{g}},$$

we get

$$\mathrm{Cov}(g_2, L_1) = r_{\mathrm{g}} h_1.$$

Therefore,

$$E(g_2 \mid L_1 > t_1) = r_{\mathrm{g}} h_1 \lambda_1.$$

Similarly,

$$E(g_1 \mid L_2 > t_2) = r_{\mathrm{g}} h_2 \lambda_2.$$

These cross-subtype terms reflect shared genetic etiology. If $r_{\mathrm{g}} > 0$, subtype 1 cases also tend to have elevated standardized genetic liability for subtype 2, and subtype 2 cases also tend to have elevated standardized genetic liability for subtype 1. This shared component reduces genetic separability between the two subtype groups.

#### 1.6 General Linear Genetic Discriminant and Mean Separation

Consider a general linear genetic discriminant on the standardized genetic scale:

$$D_w = w_1 g_1 - w_2 g_2.$$

Positive values favor subtype 1, whereas negative values favor subtype 2. The minus sign reflects that a higher value of $g_2$ supports subtype 2 rather than subtype 1.

Let

$$w = \binom{w_1}{w_2}.$$

For subtype 1 cases,

$$\mu_1(w) = E(D_w \mid L_1 > t_1).$$

Using

$$D_w = w_1 g_1 - w_2 g_2,$$

we have

$$\mu_1(w) = w_1 E(g_1 \mid L_1 > t_1) - w_2 E(g_2 \mid L_1 > t_1).$$

Substituting the conditional means,

$$E(g_1 \mid L_1 > t_1) = h_1 \lambda_1,$$

and

$$E(g_2 \mid L_1 > t_1) = r_\mathrm{g} h_1 \lambda_1,$$

gives

$$\begin{aligned} \mu_1(w) &= w_1 h_1 \lambda_1 - w_2 r_\mathrm{g} h_1 \lambda_1 \\ &= \lambda_1 h_1 (w_1 - r_\mathrm{g} w_2). \end{aligned}$$

For subtype 2 cases,

$$\mu_2(w) = E(D_w \mid L_2 > t_2).$$

Again,

$$\mu_2(w) = w_1 E(g_1 \mid L_2 > t_2) - w_2 E(g_2 \mid L_2 > t_2).$$

Using

$$E(g_1 \mid L_2 > t_2) = r_\mathrm{g} h_2 \lambda_2,$$

and

$$E(g_2 \mid L_2 > t_2) = h_2\lambda_2,$$

we obtain

$$\begin{aligned}\mu_2(w) &= w_1 r_{\mathrm{g}} h_2 \lambda_2 - w_2 h_2 \lambda_2 \\ &= \lambda_2 h_2 (r_{\mathrm{g}} w_1 - w_2).\end{aligned}$$

The between-subtype mean separation is

$$\Delta(w) = \mu_1(w) - \mu_2(w).$$

Substituting the two expressions above,

$$\begin{aligned}\Delta(w) &= \lambda_1 h_1 (w_1 - r_{\mathrm{g}} w_2) - \lambda_2 h_2 (r_{\mathrm{g}} w_1 - w_2) \\ &= w_1(\lambda_1 h_1 - r_{\mathrm{g}} \lambda_2 h_2) + w_2(\lambda_2 h_2 - r_{\mathrm{g}} \lambda_1 h_1).\end{aligned}$$

Define the signal vector, the difference between the mean of $(g_1, -g_2)^\top$ in subtype-1 cases and in subtype-2 cases,

$$a_{\mathrm{g}} = \begin{pmatrix} \lambda_1 h_1 - r_{\mathrm{g}} \lambda_2 h_2 \\ \lambda_2 h_2 - r_{\mathrm{g}} \lambda_1 h_1 \end{pmatrix}.$$

Then

$$\Delta(w) = w^\top a_{\mathrm{g}}.$$

The mean separation is thus the projection of the signal vector onto the weight vector, and its components show that the case–case genetic signal depends on subtype prevalence through $\lambda_i$, on the genetic standard deviation $h_i$, equivalently the subtype heritability $h_i^2$, and on shared genetic etiology through $r_{\mathrm{g}}$.

#### 1.7 Marginal and Conditional Variance of the General Discriminant

Define the standardized genetic contrast vector

$$X_{\mathrm{g}} = \begin{pmatrix} g_1 \\ -g_2 \end{pmatrix}.$$

Then

$$D_w = w^\top X_{\mathrm{g}}.$$

Because

$$\mathrm{Var}(g_1) = 1, \mathrm{Var}(g_2) = 1, \mathrm{Cov}(g_1, g_2) = r_\mathrm{g},$$

the covariance matrix of $X_\mathrm{g}$ is

$$B_\mathrm{g} = \mathrm{Var}(X_\mathrm{g}) = \begin{pmatrix} 1 & -r_\mathrm{g} \\ -r_\mathrm{g} & 1 \end{pmatrix}.$$

Therefore,

$$\mathrm{Var}(D_w) = w^\top B_\mathrm{g} w.$$

We now derive the within-subtype conditional variances.

For subtype 1 cases,

$$\sigma_1^2(w) = \mathrm{Var}(D_w \mid L_1 > t_1).$$

Using the conditional variance identity under liability-threshold truncation,

$$\mathrm{Var}(D_w \mid L_1 > t_1) = \mathrm{Var}(D_w) - \mathrm{Cov}(D_w, L_1)^2 \delta_1.$$

The covariance term is

$$\mathrm{Cov}(D_w, L_1) = \mathrm{Cov}(w_1 g_1 - w_2 g_2, L_1).$$

Since

$$L_1 = h_1 g_1 + \sqrt{1 - h_1^2} e_1,$$

we have

$$\mathrm{Cov}(g_1, L_1) = h_1,$$

and

$$\mathrm{Cov}(g_2, L_1) = r_\mathrm{g} h_1.$$

Therefore,

$$\begin{aligned} \mathrm{Cov}(D_w, L_1) &= w_1 h_1 - w_2 r_\mathrm{g} h_1 \\ &= h_1 (w_1 - r_\mathrm{g} w_2). \end{aligned}$$

Thus,

$$\sigma_1^2(w) = w^\top B_\mathrm{g} w - h_1^2 (w_1 - r_\mathrm{g} w_2)^2 \delta_1.$$

Similarly, for subtype 2 cases, we have

$$\sigma_2^2(w) = w^\top B_{\mathrm{g}} w - h_2^2 (r_{\mathrm{g}} w_1 - w_2)^2 \delta_2.$$

These conditional variances are exact under the single-threshold liability model.

#### 1.8 AUC-Based Signal-to-Noise Ratio and Optimal Discriminant Weights

This signal-to-noise ratio is motivated by the Gaussian approximation to the case–case AUC. If the discriminant scores in subtype 1 and subtype 2 cases are approximated by normal distributions with means $\mu_1(w)$, $\mu_2(w)$ and variances $\sigma_1^2(w)$, $\sigma_2^2(w)$, then

$$\mathrm{AUC}(w) \approx \Phi\left(\frac{\Delta(w)}{\sqrt{\sigma_1^2(w) + \sigma_2^2(w)}}\right),$$

where $\Delta(w) = \mu_1(w) - \mu_2(w)$. Because $\Phi(\cdot)$is monotone, maximizing the approximate AUC is equivalent to maximizing

$$\mathrm{SNR}(w) = \frac{\Delta(w)^2}{\sigma_1^2(w) + \sigma_2^2(w)}.$$

Using

$$\Delta(w) = w^\top a_{\mathrm{g}},$$

and the conditional variances above,

$$\sigma_1^2(w) + \sigma_2^2(w) = 2w^\top B_{\mathrm{g}} w - h_1^2 (w_1 - r_{\mathrm{g}} w_2)^2 \delta_1 - h_2^2 (r_{\mathrm{g}} w_1 - w_2)^2 \delta_2.$$

Therefore,

$$\mathrm{SNR}(w) = \frac{\left(w^\top a_{\mathrm{g}}\right)^2}{2w^\top B_{\mathrm{g}} w - h_1^2 (w_1 - r_{\mathrm{g}} w_2)^2 \delta_1 - h_2^2 (r_{\mathrm{g}} w_1 - w_2)^2 \delta_2}.$$

This ratio uses the exact conditional means and variances under the single-threshold model.

To obtain an analytic discriminant, we derive the leading-order optimal direction by omitting the truncation-induced variance corrections during the optimization step:

$$\sigma_1^2(w) \approx w^\top B_{\mathrm{g}} w, \sigma_2^2(w) \approx w^\top B_{\mathrm{g}} w.$$

Then

$$\sigma_1^2(w) + \sigma_2^2(w) \approx 2w^\top B_{\mathrm{g}} w.$$

The leading-order SNR becomes

$$\mathrm{SNR}_{\mathrm{LO}}(w) = \frac{\left(w^\top a_{\mathrm{g}}\right)^2}{2w^\top B_{\mathrm{g}} w}.$$

The factor $2$ does not affect the maximizing direction. Therefore, maximizing $\mathrm{SNR}_{\mathrm{LO}}(w)$ is equivalent to maximizing

$$f(w) = \frac{\left(w^\top a_{\mathrm{g}}\right)^2}{w^\top B_{\mathrm{g}} w}.$$

Here,

$$B_{\mathrm{g}} = \begin{pmatrix} 1 & -r_{\mathrm{g}} \\ -r_{\mathrm{g}} & 1 \end{pmatrix}.$$

When $\mid r_{\mathrm{g}} \mid < 1$, $B_{\mathrm{g}}$ is positive definite. Therefore, for any nonzero $w$,

$$w^\top B_{\mathrm{g}} w > 0.$$

Because the numerator is squared,

$$\left(w^\top a_{\mathrm{g}}\right)^2 \geq 0.$$

Thus,

$$f(w) \geq 0.$$

The value $f(w) = 0$ occurs only when

$$w^\top a_{\mathrm{g}} = 0,$$

which corresponds to a direction with no mean separation between the two subtype groups. Such directions are not useful for discrimination unless $a_{\mathrm{g}} = 0$, in which case there is no genetic separation to exploit.

The objective is also scale-invariant. For any nonzero scalar $c$,

$$f(cw) = \frac{\left((cw)^\top a_{\mathrm{g}}\right)^2}{(cw)^\top B_{\mathrm{g}}(cw)} = \frac{c^2(w^\top a_{\mathrm{g}})^2}{c^2 w^\top B_{\mathrm{g}} w} = f(w).$$

Therefore, only the direction of $w$ is identifiable; its overall magnitude is arbitrary. Differentiating $f(w)$ gives

$$\nabla_w f(w) = \frac{2(w^\top a_{\mathrm{g}})a_{\mathrm{g}}(w^\top B_{\mathrm{g}} w) - 2(w^\top a_{\mathrm{g}})^2 B_{\mathrm{g}} w}{\left(w^\top B_{\mathrm{g}} w\right)^2}.$$

At an interior optimum with nonzero mean separation, $w^\top a_{\mathrm{g}} \neq 0$. Setting the numerator to zero gives

$$2(w^\top a_{\mathrm{g}})a_{\mathrm{g}}(w^\top B_{\mathrm{g}} w) - 2(w^\top a_{\mathrm{g}})^2 B_{\mathrm{g}} w = 0.$$

Dividing by $2(w^\top a_{\mathrm{g}})$, we obtain

$$a_{\mathrm{g}}(w^\top B_{\mathrm{g}} w) - (w^\top a_{\mathrm{g}})B_{\mathrm{g}} w = 0.$$

Therefore,

$$B_{\mathrm{g}} w = \frac{w^\top B_{\mathrm{g}} w}{w^\top a_{\mathrm{g}}} a_{\mathrm{g}}.$$

The scalar

$$\frac{w^\top B_{\mathrm{g}} w}{w^\top a_{\mathrm{g}}}$$

only changes the magnitude of $w$, not its direction. Hence,

$$B_{\mathrm{g}} w \propto a_{\mathrm{g}}.$$

Since $B_{\mathrm{g}}$ is invertible when $| r_{\mathrm{g}} | < 1$, the leading-order SNR-maximizing direction is

$$w_{\mathrm{LO}}^* \propto B_{\mathrm{g}}^{-1} a_{\mathrm{g}}.$$

Now recall that

$$a_{\mathrm{g}} = \begin{pmatrix} \lambda_1 h_1 - r_{\mathrm{g}} \lambda_2 h_2 \\ \lambda_2 h_2 - r_{\mathrm{g}} \lambda_1 h_1 \end{pmatrix},$$

and

$$B_{\mathrm{g}} = \begin{pmatrix} 1 & -r_{\mathrm{g}} \\ -r_{\mathrm{g}} & 1 \end{pmatrix}.$$

We can factorize $a_{\mathrm{g}}$ as

$$a_{\mathrm{g}} = B_{\mathrm{g}} \begin{pmatrix} \lambda_1 h_1 \\ \lambda_2 h_2 \end{pmatrix}.$$

Therefore,

$$B_{\mathrm{g}}^{-1} a_{\mathrm{g}} = \begin{pmatrix} \lambda_1 h_1 \\ \lambda_2 h_2 \end{pmatrix}.$$

Thus, the leading-order optimal weights are, for a common constant $c > 0$,

$$w_1^* = c\lambda_1 h_1, \qquad w_2^* = c\lambda_2 h_2.$$

The squared SNR does not determine the sign of $w$, because it depends on

$$\left(w^\top a_{\mathrm{g}}\right)^2.$$

We choose the sign such that the mean separation $\Delta(w) = w^\top a_{\mathrm{g}}$ is positive for subtype 1 relative to subtype 2.

Therefore, taking $c = 1$, the leading-order optimal discriminant is

$$D_\lambda = \lambda_1 h_1 g_1 - \lambda_2 h_2 g_2.$$

Since

$$G_i = h_i g_i,$$

this is equivalent to the unstandardized form

$$D_\lambda = \lambda_1 G_1 - \lambda_2 G_2.$$

The standardized form makes the weight structure explicit:

$$w_i = \lambda_i h_i.$$

Thus, subtype prevalence and subtype heritability together determine the leading-order optimal weights.

When the two subtypes have equal prevalence,

$$K_1 = K_2,$$

we have

$$\lambda_1 = \lambda_2.$$

Then

$$D_\lambda \propto h_1 g_1 - h_2 g_2 = G_1 - G_2.$$

If the two subtypes also have equal heritability, then

$$D_\lambda \propto g_1 - g_2.$$

#### 1.9 Genetic Separation Variance $V_S$

Using the leading-order optimal discriminant,

$$D_\lambda = \lambda_1 h_1 g_1 - \lambda_2 h_2 g_2,$$

the marginal variance is

$$\mathrm{Var}(D_\lambda) = \mathrm{Var}(\lambda_1 h_1 g_1 - \lambda_2 h_2 g_2).$$

Using

$$\mathrm{Var}(g_1) = 1, \mathrm{Var}(g_2) = 1, \mathrm{Cov}(g_1, g_2) = r_g,$$

we obtain

$$\mathrm{Var}(D_\lambda) \quad = \lambda_1^2 h_1^2 + \lambda_2^2 h_2^2 - 2\lambda_1\lambda_2 r_g h_1 h_2.$$

Define

$$V_S = \lambda_1^2 h_1^2 + \lambda_2^2 h_2^2 - 2\lambda_1\lambda_2 r_g h_1 h_2.$$

Thus,

$$\mathrm{Var}(D_\lambda) = V_S.$$

The quantity $V_S$ measures the genetic separation variance between the two subtypes on the standardized genetic-value scale with the leading-order optimal, prevalence- and heritability-weighted coefficients.

$V_S$ is defined for the canonical scaling $D_\lambda = \lambda_1 h_1 g_1 - \lambda_2 h_2 g_2$ and changes if the weights are rescaled. Multiplying the weight vector by a positive constant leaves the ordering of the scores, and therefore the case–case AUC, unchanged, but multiplies the mean separation by that constant and every variance by its square. The compact results of Sections 1.10 and 1.13, $\Delta_\lambda = \mathrm{Var}(D_\lambda) = V_\mathrm{S}$ and $\mathrm{AUC}_{\mathrm{cc}}^{\mathrm{TGV}} \approx \Phi\left(\sqrt{V_\mathrm{S}/2}\right)$, therefore hold only for the canonical scaling; for any other scaling the general expressions in $\Delta(w)$ and $\sigma_i^2(w)$ apply. A negative constant reverses the ordering and gives $1 - \mathrm{AUC}$, so the sign of the weights is fixed by requiring subtype 1 cases to score higher on average.

#### 1.10 Mean Separation Under the Optimal Discriminant

For subtype 1 cases,

$$\mu_1 = E(D_\lambda \mid L_1 > t_1).$$

Substituting the discriminant,

$$\mu_1 = \lambda_1 h_1 E(g_1 \mid L_1 > t_1) - \lambda_2 h_2 E(g_2 \mid L_1 > t_1).$$

Using the conditional means of Section 1.5,

$$\begin{aligned} \mu_1 &= \lambda_1 h_1 (h_1 \lambda_1) - \lambda_2 h_2 (r_\mathrm{g} h_1 \lambda_1) \\ &= \lambda_1^2 h_1^2 - \lambda_1 \lambda_2 r_\mathrm{g} h_1 h_2. \end{aligned}$$

For subtype 2 cases,

$$\mu_2 = E(D_\lambda \mid L_2 > t_2).$$

Thus,

$$\mu_2 = \lambda_1 h_1 E(g_1 \mid L_2 > t_2) - \lambda_2 h_2 E(g_2 \mid L_2 > t_2).$$

Using the conditional means of Section 1.5,

$$\begin{aligned} \mu_2 &= \lambda_1 h_1 (r_\mathrm{g} h_2 \lambda_2) - \lambda_2 h_2 (h_2 \lambda_2) \\ &= \lambda_1 \lambda_2 r_\mathrm{g} h_1 h_2 - \lambda_2^2 h_2^2. \end{aligned}$$

The between-subtype mean separation is

$$\Delta_\lambda = \mu_1 - \mu_2.$$

Substituting the expressions above,

$$\begin{aligned} \Delta_\lambda &= \lambda_1^2 h_1^2 - \lambda_1 \lambda_2 r_\mathrm{g} h_1 h_2 - \lambda_1 \lambda_2 r_\mathrm{g} h_1 h_2 + \lambda_2^2 h_2^2 \\ &= \lambda_1^2 h_1^2 + \lambda_2^2 h_2^2 - 2 \lambda_1 \lambda_2 r_\mathrm{g} h_1 h_2. \end{aligned}$$

Therefore,

$$\Delta_\lambda = V_S.$$

With the canonical scaling of $D_\lambda$ (Section 1.9), the between-subtype mean separation and the marginal variance of the discriminant are both equal to $V_S$.

#### 1.11 Conditional Variance Under the Optimal Discriminant

We now compute the within-subtype conditional variances of the leading-order optimal discriminant. Since

$$\mathrm{Var}(D_\lambda) = V_S,$$

the truncation variance identity gives

$$\mathrm{Var}(D_\lambda \mid L_i > t_i) = V_S - \mathrm{Cov}(D_\lambda, L_i)^2 \delta_i.$$

For subtype 1 cases,

$$\sigma_1^2 = \mathrm{Var}(D_\lambda \mid L_1 > t_1) = V_S - \mathrm{Cov}(D_\lambda, L_1)^2 \delta_1.$$

Now,

$$\mathrm{Cov}(D_\lambda, L_1) \quad = \mathrm{Cov}(\lambda_1 h_1 g_1 - \lambda_2 h_2 g_2, L_1).$$

Using the covariances of Section 1.5,

$$\begin{aligned} \mathrm{Cov}(D_\lambda, L_1) \quad &= \lambda_1 h_1 h_1 - \lambda_2 h_2 r_g h_1 \\ &= \lambda_1 h_1^2 - \lambda_2 r_g h_1 h_2. \end{aligned}$$

Define

$$a_1 = \lambda_1 h_1^2 - \lambda_2 r_g h_1 h_2.$$

Then

$$\sigma_1^2 = V_S - a_1^2 \delta_1.$$

For subtype 2 cases,

$$\sigma_2^2 = \mathrm{Var}(D_\lambda \mid L_2 > t_2) = V_S - \mathrm{Cov}(D_\lambda, L_2)^2 \delta_2.$$

Using the covariances of Section 1.5,

$$\begin{aligned} \mathrm{Cov}(D_\lambda, L_2) \quad &= \lambda_1 h_1 r_g h_2 - \lambda_2 h_2 h_2 \\ &= \lambda_1 r_g h_1 h_2 - \lambda_2 h_2^2. \end{aligned}$$

Since this covariance enters the variance formula through its square, define

$$a_2 = \lambda_2 h_2^2 - \lambda_1 r_g h_1 h_2.$$

Then

$$\text{Cov}(D_\lambda, L_2)^2 = a_2^2.$$

Therefore,

$$\sigma_2^2 = V_S - a_2^2 \delta_2.$$

Adding the two within-subtype variances gives

$$\sigma_1^2 + \sigma_2^2 = 2V_S - a_1^2 \delta_1 - a_2^2 \delta_2.$$

These conditional variances are exact under the single-threshold liability model. The approximation enters later when we approximate the conditional distributions of $D_\lambda$ as Gaussian for AUC calculation.

**1.12 Case–Case AUC Based on True Genetic Values**

The case–case AUC is defined as

$$\text{AUC}_{\text{cc}}^{\text{TGV}} = P(D_{\lambda,1} > D_{\lambda,2}),$$

where $D_{\lambda,1}$ is the discriminant score for a randomly selected subtype 1 case, and $D_{\lambda,2}$ is the score for a randomly selected subtype 2 case.

The exact conditional distributions of $D_{\lambda,1}$ and $D_{\lambda,2}$ after liability-threshold selection are not generally Gaussian. However, their conditional means and variances are given above. Using a moment-matched Gaussian approximation,

$$D_{\lambda,1} \approx N(\mu_1, \sigma_1^2),$$

and

$$D_{\lambda,2} \approx N(\mu_2, \sigma_2^2).$$

Assuming the two sampled individuals are independent,

$$D_{\lambda,1} - D_{\lambda,2} \approx N(\mu_1 - \mu_2, \sigma_1^2 + \sigma_2^2).$$

Therefore,

$$\begin{aligned}\text{AUC}_{\text{cc}}^{\text{TGV}} &= P(D_{\lambda,1} > D_{\lambda,2}) \\ &= P(D_{\lambda,1} - D_{\lambda,2} > 0) \\ &\approx \Phi\left(\frac{\mu_1 - \mu_2}{\sqrt{\sigma_1^2 + \sigma_2^2}}\right).\end{aligned}$$

Because

$$\mu_1 - \mu_2 = V_{\mathrm{S}},$$

and

$$\sigma_1^2 + \sigma_2^2 = 2V_{\mathrm{S}} - a_1^2\delta_1 - a_2^2\delta_2,$$

we obtain

$$\mathrm{AUC}_{\mathrm{cc}}^{\mathrm{TGV}} \approx \Phi\left(\frac{V_{\mathrm{S}}}{\sqrt{2V_{\mathrm{S}} - a_1^2\delta_1 - a_2^2\delta_2}}\right).$$

Here,

$$V_{\mathrm{S}} = \lambda_1^2 h_1^2 + \lambda_2^2 h_2^2 - 2\lambda_1\lambda_2 r_{\mathrm{g}} h_1 h_2,$$

$$a_1 = \lambda_1 h_1^2 - \lambda_2 r_{\mathrm{g}} h_1 h_2,$$

$$a_2 = \lambda_2 h_2^2 - \lambda_1 r_{\mathrm{g}} h_1 h_2,$$

and

$$\delta_i = \lambda_i(\lambda_i - t_i).$$

The truncation term $a_1^2\delta_1 + a_2^2\delta_2$ is denoted $C_{\mathrm{trunc}}$ in main-text equation (2).

This AUC expression uses exact conditional first and second moments under the single-threshold liability model, but the final AUC calculation is approximate because it uses a Gaussian approximation for the within-subtype discriminant distributions.

### 1.13 Leading-Order AUC Approximation

If the truncation correction $C_{\mathrm{trunc}}$ of Section 1.12 is small relative to $2V_{\mathrm{S}}$ and is therefore ignored ($\frac{C_{\mathrm{trunc}}}{2V_{\mathrm{S}}} \ll 1$ with $V_{\mathrm{S}} > 0$; when $V_{\mathrm{S}} = 0$ there is no mean separation and the AUC is 0.5), that is,

$$a_1^2\delta_1 \approx 0, a_2^2\delta_2 \approx 0,$$

then

$$\sigma_1^2 + \sigma_2^2 \approx 2V_{\mathrm{S}}.$$

The AUC simplifies to

$$\begin{aligned}\mathrm{AUC}_{\mathrm{cc}}^{\mathrm{TGV}} &\approx \Phi\left(\frac{V_{\mathrm{S}}}{\sqrt{2V_{\mathrm{S}}}}\right)\\ &= \Phi\left(\sqrt{\frac{V_{\mathrm{S}}}{2}}\right).\end{aligned}$$

This is the leading-order expression, referred to in the main text as the simplified approximation (main-text equation (3)). It shows that the primary determinant of case–case genetic discrimination is the genetic separation variance $V_{\mathrm{S}}$. The subtype prevalences $K_1$ and $K_2$ enter the AUC through the selection intensities $\lambda_1$ and $\lambda_2$, while subtype heritabilities enter through $h_1^2$, $h_2^2$, and $h_1 h_2$.

#### 1.14 Relationship Between $V_{\mathrm{S}}$ and GDIS

GDIS[2] summarizes the genetic distance between two subtypes on the 50/50 observed scale, whereas $V_{\mathrm{S}}$ (Section 1.9) is defined on the liability-threshold discriminant scale and determines the leading-order oracle AUC $\Phi(\sqrt{V_{\mathrm{S}}/2})$ (Section 1.13). We relate the two using the standard observed-to-liability heritability conversion, with the same subtype-specific heritabilities, genetic correlation and prevalences as inputs.

To avoid confusion with liability-scale heritability $h_i^2$, let $H_i^{50}$ denote the subtype $i$-versus-control SNP heritability on the 50/50 observed scale. The squared GDIS distance can be written as

$$H_{12}^{50} = H_1^{50} + H_2^{50} - 2r_{\mathrm{g}}\sqrt{H_1^{50} H_2^{50}}.$$

The GDIS distance is its square root,

$$d_{\mathrm{GDIS}} = \sqrt{H_{12}^{50}}.$$

In this section we write $H_{12}^{50}$ for the squared GDIS distance.

To connect GDIS with $V_{\mathrm{S}}$, we first convert the subtype–control heritabilities from the 50/50 observed scale to the liability scale. The standard observed-to-liability conversion[3] gives

$$h_i^2 = H_i^{50}\frac{K_i^2(1-K_i)^2}{P_i(1-P_i)\phi(t_i)^2}.$$

On the 50/50 observed scale,

$$P_i = \frac{1}{2}, \qquad P_i(1 - P_i) = \frac{1}{4}.$$

Moreover, by the definition of the selection intensity (Section 1.3),

$$\phi(t_i) = K_i \lambda_i.$$

Substituting these into the conversion formula gives

$$h_i^2 = \frac{4(1 - K_i)^2}{\lambda_i^2} H_i^{50}.$$

Substituting these expressions into $V_{\mathrm{S}}$, we obtain

$$\lambda_1^2 h_1^2 = 4(1 - K_1)^2 H_1^{50}, \lambda_2^2 h_2^2 = 4(1 - K_2)^2 H_2^{50},$$

and

$$2\lambda_1 \lambda_2 r_{\mathrm{g}} h_1 h_2 = 8 r_{\mathrm{g}} (1 - K_1)(1 - K_2)\sqrt{H_1^{50} H_2^{50}}.$$

Therefore,

$$V_{\mathrm{S}} = 4 Q_{12},$$

where

$$Q_{12} = (1 - K_1)^2 H_1^{50} + (1 - K_2)^2 H_2^{50} - 2 r_{\mathrm{g}} (1 - K_1)(1 - K_2)\sqrt{H_1^{50} H_2^{50}}.$$

Note that $Q_{12}$ is not a function of $H_{12}^{50}$ alone: it requires the two subtype–control heritabilities $H_1^{50}$ and $H_2^{50}$, the genetic correlation and the two prevalences separately. In general, $V_{\mathrm{S}}$ cannot be recovered from a single GDIS distance; when the prevalences are equal and known, the relationship simplifies as follows.

A useful special case occurs when the two subtype prevalences are equal:

$$K_1 = K_2 = K.$$

Then

$$Q_{12} = (1 - K)^2 \left[ H_1^{50} + H_2^{50} - 2 r_{\mathrm{g}} \sqrt{H_1^{50} H_2^{50}} \right].$$

Since the term inside brackets is the squared GDIS distance $H_{12}^{50}$, we have

$$Q_{12} = (1 - K)^2 H_{12}^{50}.$$

Therefore,

$$V_S = 4(1-K)^2 H_{12}^{50}.$$

For rare subtypes ($1 - K \approx 1$) this is approximately $4H_{12}^{50}$; the factor 4 comes from the observed-to-liability conversion and does not vanish in that limit.

$V_S$ is therefore proportional to the squared GDIS distance when the subtype prevalences are equal, with factor $4(1-K)^2$. When the prevalences differ, converting a GDIS distance to $V_S$ generally requires the two subtype-specific heritabilities, their genetic correlation and both prevalences.

**Supplementary Note 2: PRS-Based Case–Case AUC and Optimal Genetic Discriminant under the Liability-Threshold Model**

### 2.1 Overview

We extend the true-genetic-value oracle of Supplementary Note 1 to subtype-specific polygenic risk score (PRS) estimates with finite prediction accuracy and derive their case–case AUC under the same liability-threshold model.

We work under the same bivariate liability-threshold model as Supplementary Note 1, on the standardized genetic-value scale $G_i = h_i g_i$, and write $\hat{g}_i$ for the PRS estimate of $g_i$, with $R_i = \mathrm{Cor}(\hat{g}_i, g_i)$ and PRS prediction accuracy $R_i^2$.

Our goal is to derive the optimal linear PRS discriminant

$$S_w = w_1 \hat{g}_1 - w_2 \hat{g}_2$$

for separating subtype 1 cases from subtype 2 cases, and to obtain the corresponding case–case AUC.

The key result is that the leading-order optimal PRS weights are

$$w_{\mathrm{LO}}^* \propto \Sigma_{\mathrm{PRS}}^{-1} a_{\mathrm{PRS}},$$

where $a_{\mathrm{PRS}}$ is the difference between the case-group means of the PRS contrast vector $X_{\mathrm{PRS}}$ (Sections 2.4 and 2.5) and $\Sigma_{\mathrm{PRS}}$ is its covariance matrix in the population (Section 2.5).

In closed form, the leading-order optimal weights are, for a common constant $c > 0$,

$$w_1^* = cR_1\left[\lambda_1 h_1 (1 - R_2^2 r_{\mathrm{g}}^2) - \lambda_2 h_2 r_{\mathrm{g}} (1 - R_2^2)\right],$$

and

$$w_2^* = cR_2\left[\lambda_2 h_2 (1 - R_1^2 r_{\mathrm{g}}^2) - \lambda_1 h_1 r_{\mathrm{g}} (1 - R_1^2)\right].$$

The corresponding PRS genetic separation variance is

$$V_{\mathrm{PRS}} = a_{\mathrm{PRS}}^{\top} \Sigma_{\mathrm{PRS}}^{-1} a_{\mathrm{PRS}}.$$

Its closed form is

$$V_{\mathrm{PRS}} = \frac{R_1^2 A^2 + R_2^2 B^2 + 2\rho R_1 R_2 AB}{1 - \rho^2}.$$

Here, $A$ and $B$ are the signal components of Section 2.9, $A = \lambda_1 h_1 - r_{\mathrm{g}} \lambda_2 h_2$ and

$B = \lambda_2 h_2 - r_g \lambda_1 h_1$, and $\rho = R_1 R_2 r_g$ is the correlation between the two PRS estimates (Section 2.5). $V_{\text{PRS}}$ is the PRS counterpart of $V_S$: with the canonical scaling $w^*_{\text{LO}} = \Sigma^{-1}_{\text{PRS}} a_{\text{PRS}}$ (Section 2.10), the between-subtype mean separation and the marginal variance of the discriminant both equal $V_{\text{PRS}}$, so that, omitting the truncation corrections, the leading-order PRS-based case–case AUC is

$$\text{AUC}^{\text{PRS}}_{\text{cc}} \approx \Phi\left(\sqrt{\frac{V_{\text{PRS}}}{2}}\right).$$

This is the counterpart of main-text equation (3). The moment-based AUC for an arbitrary weight vector, including the truncation corrections, is given in Section 2.7.

When

$$R_1^2 = R_2^2 = 1,$$

or equivalently $R_1 = R_2 = 1$, the PRS estimates become the true standardized genetic values and the true-genetic-value oracle discriminant of Supplementary Note 1 is recovered exactly, on that same standardized genetic-value scale.

**2.2 PRS Model**

The liability-threshold model, the thresholds and selection intensities, and the truncation identities are those of Supplementary Note 1.2–1.4 and are used unchanged. We model the PRS estimate of the standardized genetic value $g_i$ as

$$\hat{g}_i = R_i g_i + \sqrt{1 - R_i^2}\eta_i,$$

where

$$\eta_i \sim N(0,1)$$

is PRS noise independent of all liability components.

By construction,

$$R_i = \text{Cor}(\hat{g}_i, g_i),$$

and therefore

$$R_i^2 = \text{Cor}(\hat{g}_i, g_i)^2$$

is the PRS prediction accuracy. Scores are oriented to be positively correlated with their target genetic values, so that $0 \leq R_i \leq 1$; we use $R_i^2$ for prediction accuracy on the genetic scale and $R_i$ for the attenuation of means and covariances.

Under this model, the population variance is

$$\mathrm{Var}(\hat{g}_i) = 1.$$

Also,

$$\mathrm{Cov}(\hat{g}_i, g_i) = R_i.$$

We assume the PRS noise terms are independent of the true genetic values, residual components, and each other:

$$\mathrm{Cov}(\eta_i, g_j) = 0, \mathrm{Cov}(\eta_i, e_j) = 0,$$

and

$$\mathrm{Cov}(\eta_1, \eta_2) = 0.$$

The covariance between the two PRS estimates is

$$\mathrm{Cov}(\hat{g}_1, \hat{g}_2) = \mathrm{Cov}\left(R_1 g_1 + \sqrt{1 - R_1^2}\eta_1, R_2 g_2 + \sqrt{1 - R_2^2}\eta_2\right).$$

Because the PRS noise terms are independent,

$$\mathrm{Cov}(\hat{g}_1, \hat{g}_2) = R_1 R_2 \mathrm{Cov}(g_1, g_2).$$

Since

$$\mathrm{Cov}(g_1, g_2) = r_\mathrm{g},$$

we obtain

$$\mathrm{Cov}(\hat{g}_1, \hat{g}_2) = R_1 R_2 r_\mathrm{g}.$$

Define

$$\rho = R_1 R_2 r_\mathrm{g}.$$

Then

$$\mathrm{Cov}(\hat{g}_1, \hat{g}_2) = \rho.$$

When $R_i^2 = 1$, we have $R_i = 1$ and

$$\hat{g}_i = g_i.$$

When $R_i^2 = 0$, we have $R_i = 0$, and the PRS estimate contains no information about the true standardized genetic value.

#### 2.3 Conditional Means of PRS Estimates Among Subtype Cases

We first compute the conditional means of PRS estimates among subtype cases.

For subtype $i$, cases satisfy

$$L_i > t_i.$$

Using the conditional mean identity,

$$E(\hat{g}_j \mid L_i > t_i) = \mathrm{Cov}(\hat{g}_j, L_i)\lambda_i.$$

Therefore, we need the covariances between PRS estimates and liabilities.

For subtype 1 liability,

$$L_1 = h_1 g_1 + \sqrt{1 - h_1^2}e_1.$$

First,

$$\mathrm{Cov}(\hat{g}_1, L_1) = \mathrm{Cov}\left(R_1 g_1 + \sqrt{1 - R_1^2}\eta_1, h_1 g_1 + \sqrt{1 - h_1^2}e_1\right).$$

Because $\eta_1$ is independent of all liability components and $g_1$ is independent of $e_1$,

$$\mathrm{Cov}(\hat{g}_1, L_1) = R_1 h_1 \mathrm{Var}(g_1).$$

Since

$$\mathrm{Var}(g_1) = 1,$$

we obtain

$$\mathrm{Cov}(\hat{g}_1, L_1) = R_1 h_1.$$

Second,

$$\mathrm{Cov}(\hat{g}_2, L_1) = \mathrm{Cov}\left(R_2 g_2 + \sqrt{1 - R_2^2}\eta_2, h_1 g_1 + \sqrt{1 - h_1^2}e_1\right).$$

Again, the PRS noise is independent, and $g_2$ is independent of $e_1$. Therefore,

$$\mathrm{Cov}(\hat{g}_2, L_1) = R_2 h_1 \mathrm{Cov}(g_2, g_1).$$

Since

$$\mathrm{Cov}(g_2, g_1) = r_\mathrm{g},$$

we obtain

$$\mathrm{Cov}(\hat{g}_2, L_1) = R_2 r_{\mathrm{g}} h_1.$$

Therefore,

$$E(\hat{g}_1 \mid L_1 > t_1) = R_1 h_1 \lambda_1,$$

and

$$E(\hat{g}_2 \mid L_1 > t_1) = R_2 r_{\mathrm{g}} h_1 \lambda_1.$$

Similarly, for subtype 2 liability,

$$L_2 = h_2 g_2 + \sqrt{1 - h_2^2} e_2.$$

We have

$$\mathrm{Cov}(\hat{g}_2, L_2) = R_2 h_2,$$

and

$$\mathrm{Cov}(\hat{g}_1, L_2) = R_1 r_{\mathrm{g}} h_2.$$

Therefore,

$$E(\hat{g}_2 \mid L_2 > t_2) = R_2 h_2 \lambda_2,$$

and

$$E(\hat{g}_1 \mid L_2 > t_2) = R_1 r_{\mathrm{g}} h_2 \lambda_2.$$

These expressions show that finite PRS accuracy attenuates the conditional mean shift by a factor $R_i = \mathrm{Cor}(\hat{g}_i, g_i)$. If $R_i = 1$, the PRS estimate behaves like the true standardized genetic value. If $R_i = 0$, the PRS has no conditional mean shift after liability-threshold selection.

#### 2.4 General Linear PRS Discriminant and Mean Separation

Consider a general linear PRS discriminant

$$S_w = w_1 \hat{g}_1 - w_2 \hat{g}_2.$$

Positive values favor subtype 1, whereas negative values favor subtype 2. The minus sign reflects that a higher PRS for subtype 2 should support subtype 2 rather than subtype 1.

For subtype 1 cases,

$$\mu_1(w) = E(S_w \mid L_1 > t_1).$$

Using the conditional PRS means,

$$\begin{aligned}\mu_1(w) &= w_1 E(\hat{g}_1 \mid L_1 > t_1) - w_2 E(\hat{g}_2 \mid L_1 > t_1) \\ &= w_1 R_1 h_1 \lambda_1 - w_2 R_2 r_{\mathrm{g}} h_1 \lambda_1 \\ &= \lambda_1 h_1 (R_1 w_1 - R_2 r_{\mathrm{g}} w_2).\end{aligned}$$

For subtype 2 cases,

$$\mu_2(w) = E(S_w \mid L_2 > t_2).$$

Using the conditional PRS means,

$$\begin{aligned}\mu_2(w) &= w_1 E(\hat{g}_1 \mid L_2 > t_2) - w_2 E(\hat{g}_2 \mid L_2 > t_2) \\ &= w_1 R_1 r_{\mathrm{g}} h_2 \lambda_2 - w_2 R_2 h_2 \lambda_2 \\ &= \lambda_2 h_2 (R_1 r_{\mathrm{g}} w_1 - R_2 w_2).\end{aligned}$$

The between-subtype mean separation is

$$\Delta(w) = \mu_1(w) - \mu_2(w).$$

Substituting the expressions above,

$$\begin{aligned}\Delta(w) &= \lambda_1 h_1 (R_1 w_1 - R_2 r_{\mathrm{g}} w_2) - \lambda_2 h_2 (R_1 r_{\mathrm{g}} w_1 - R_2 w_2) \\ &= w_1 R_1 (\lambda_1 h_1 - r_{\mathrm{g}} \lambda_2 h_2) + w_2 R_2 (\lambda_2 h_2 - r_{\mathrm{g}} \lambda_1 h_1).\end{aligned}$$

Define the PRS signal vector

$$a_{\mathrm{PRS}} = \begin{pmatrix} R_1(\lambda_1 h_1 - r_{\mathrm{g}} \lambda_2 h_2) \\ R_2(\lambda_2 h_2 - r_{\mathrm{g}} \lambda_1 h_1) \end{pmatrix}.$$

Then

$$\Delta(w) = w^\top a_{\mathrm{PRS}}.$$

This expression shows that the PRS-based mean-separation signal is attenuated component-wise by PRS accuracy.

Compared with the true-genetic-value signal vector $a_{\mathrm{g}}$ of Section 1.6, written here as

$$a_{\mathrm{TGV}} = \begin{pmatrix} \lambda_1 h_1 - r_{\mathrm{g}} \lambda_2 h_2 \\ \lambda_2 h_2 - r_{\mathrm{g}} \lambda_1 h_1 \end{pmatrix},$$

the PRS signal vector is

$$a_{\text{PRS}} = \begin{pmatrix} R_1 a_{TGV,1} \\ R_2 a_{TGV,2} \end{pmatrix}.$$

Thus, each component of the subtype mean-separation signal is attenuated by the corresponding PRS correlation $R_i$, whose square $R_i^2$ is the PRS prediction accuracy.

**2.5 Marginal Covariance of the PRS Discriminant**

Define the PRS contrast vector

$$X_{\text{PRS}} = \begin{pmatrix} \hat{g}_1 \\ -\hat{g}_2 \end{pmatrix}.$$

Then

$$S_w = w^\top X_{\text{PRS}}.$$

Because

$$\text{Var}(\hat{g}_1) = 1, \text{Var}(\hat{g}_2) = 1,$$

and

$$\text{Cov}(\hat{g}_1, \hat{g}_2) = \rho = R_1 R_2 r_{\text{g}},$$

the covariance matrix of $X_{\text{PRS}}$ is

$$\Sigma_{\text{PRS}} = \text{Var}(X_{\text{PRS}}) = \begin{pmatrix} 1 & -\rho \\ -\rho & 1 \end{pmatrix}.$$

Therefore,

$$\text{Var}(S_w) = w^\top \Sigma_{\text{PRS}} w.$$

When $R_1^2 = R_2^2 = 1$, we have $R_1 = R_2 = 1$, $\rho = r_{\text{g}}$, and $\Sigma_{\text{PRS}}$ reduces to the covariance matrix of the true standardized genetic contrast.

When $R_1^2 = R_2^2 = 0$, we have $R_1 = R_2 = 0$, $\rho = 0$, and the PRS estimates are pure noise with no genetic signal.

**2.6 Conditional Variance of the General PRS Discriminant**

We now derive the within-subtype conditional variances of $S_w$.

For subtype 1 cases,

$$\sigma_1^2(w) = \text{Var}(S_w \mid L_1 > t_1).$$

Using the conditional variance identity under liability-threshold truncation,

$$\sigma_1^2(w) = \mathrm{Var}(S_w) - \mathrm{Cov}(S_w, L_1)^2 \delta_1.$$

We already have

$$\mathrm{Var}(S_w) = w^\top \Sigma_{\mathrm{PRS}} w.$$

Now,

$$\mathrm{Cov}(S_w, L_1) = \mathrm{Cov}(w_1 \hat{g}_1 - w_2 \hat{g}_2, L_1).$$

Using

$$\mathrm{Cov}(\hat{g}_1, L_1) = R_1 h_1,$$

and

$$\mathrm{Cov}(\hat{g}_2, L_1) = R_2 r_{\mathrm{g}} h_1,$$

we get

$$\begin{aligned} \mathrm{Cov}(S_w, L_1) &= w_1 R_1 h_1 - w_2 R_2 r_{\mathrm{g}} h_1 \\ &= h_1 (R_1 w_1 - R_2 r_{\mathrm{g}} w_2). \end{aligned}$$

Define

$$d_1 = h_1 \begin{pmatrix} R_1 \\ -R_2 r_{\mathrm{g}} \end{pmatrix}.$$

Then

$$\mathrm{Cov}(S_w, L_1) = w^\top d_1.$$

Therefore,

$$\sigma_1^2(w) = w^\top \Sigma_{\mathrm{PRS}} w - (w^\top d_1)^2 \delta_1.$$

For subtype 2 cases,

$$\sigma_2^2(w) = \mathrm{Var}(S_w \mid L_2 > t_2).$$

Similarly,

$$\sigma_2^2(w) = \mathrm{Var}(S_w) - \mathrm{Cov}(S_w, L_2)^2 \delta_2.$$

Using

$$\mathrm{Cov}(\hat{g}_1, L_2) = R_1 r_{\mathrm{g}} h_2,$$

and

$$\mathrm{Cov}(\hat{g}_2, L_2) = R_2 h_2,$$

we obtain

$$\begin{aligned}\mathrm{Cov}(S_w, L_2) &= w_1 R_1 r_{\mathrm{g}} h_2 - w_2 R_2 h_2 \\ &= h_2 (R_1 r_{\mathrm{g}} w_1 - R_2 w_2).\end{aligned}$$

Define

$$d_2 = h_2 \begin{pmatrix} R_1 r_{\mathrm{g}} \\ -R_2 \end{pmatrix}.$$

Then

$$\mathrm{Cov}(S_w, L_2) = w^\top d_2.$$

Therefore,

$$\sigma_2^2(w) = w^\top \Sigma_{\mathrm{PRS}} w - (w^\top d_2)^2 \delta_2.$$

Adding the two conditional variances gives

$$\sigma_1^2(w) + \sigma_2^2(w) = 2 w^\top \Sigma_{\mathrm{PRS}} w - (w^\top d_1)^2 \delta_1 - (w^\top d_2)^2 \delta_2.$$

These conditional variances are exact under the single-threshold liability model.

**2.7 Case–Case AUC for a General PRS Discriminant**

The case–case AUC for the PRS discriminant is

$$\mathrm{AUC}_{\mathrm{cc}}^{\mathrm{PRS}}(w) = P(S_{w,1} > S_{w,2}),$$

where $S_{w,1}$ is the score for a randomly selected subtype 1 case and $S_{w,2}$ is the score for a randomly selected subtype 2 case.

The exact conditional distributions of $S_w$ after liability-threshold selection are not generally Gaussian. However, their first and second moments are given above. Using a moment-matched Gaussian approximation,

$$S_{w,1} \approx N(\mu_1(w), \sigma_1^2(w)),$$

and

$$S_{w,2} \approx N(\mu_2(w), \sigma_2^2(w)).$$

Assuming the two sampled individuals are independent,

$$S_{w,1} - S_{w,2} \approx N(\Delta(w), \sigma_1^2(w) + \sigma_2^2(w)).$$

Therefore,

$$\mathrm{AUC}_{\mathrm{cc}}^{\mathrm{PRS}}(w) \approx \Phi\left(\frac{\Delta(w)}{\sqrt{\sigma_1^2(w) + \sigma_2^2(w)}}\right).$$

Using

$$\Delta(w) = w^\top a_{\mathrm{PRS}},$$

and

$$\sigma_1^2(w) + \sigma_2^2(w) = 2w^\top \Sigma_{\mathrm{PRS}} w - (w^\top d_1)^2 \delta_1 - (w^\top d_2)^2 \delta_2,$$

we obtain

$$\mathrm{AUC}_{\mathrm{cc}}^{\mathrm{PRS}}(w) \approx \Phi\left(\frac{w^\top a_{\mathrm{PRS}}}{\sqrt{2w^\top \Sigma_{\mathrm{PRS}} w - (w^\top d_1)^2 \delta_1 - (w^\top d_2)^2 \delta_2}}\right).$$

The sign of $w$ is chosen so that

$$\Delta(w) = w^\top a_{\mathrm{PRS}} > 0.$$

This AUC formula applies to any PRS weight vector $w$. In the derivations that follow we use the leading-order optimal weights of Section 2.9; the weights used in the empirical analyses are stated in Supplementary Note 3.5.2.

**2.8 AUC-Based Signal-to-Noise Ratio**

The moment-based AUC approximation motivates the signal-to-noise ratio

$$\mathrm{SNR}(w) = \frac{\Delta(w)^2}{\sigma_1^2(w) + \sigma_2^2(w)}.$$

Substituting the expressions above,

$$\mathrm{SNR}(w) = \frac{(w^\top a_{\mathrm{PRS}})^2}{2w^\top \Sigma_{\mathrm{PRS}} w - (w^\top d_1)^2 \delta_1 - (w^\top d_2)^2 \delta_2}.$$

This ratio uses the exact conditional means and variances under the single-threshold model.

Since the AUC approximation is monotone in

$$\frac{\Delta(w)}{\sqrt{\sigma_1^2(w) + \sigma_2^2(w)}},$$

maximizing the approximate AUC is equivalent to maximizing this SNR, with the sign of $w$ chosen so that

$$\Delta(w) > 0.$$

**2.9 Leading-Order Optimal PRS Weights in Closed Form**

To obtain an analytic discriminant, we derive the leading-order optimal direction by omitting the truncation-induced variance corrections during the optimization step. That is, we drop the truncation corrections from both within-subtype variances, leaving

$$\sigma_1^2(w) + \sigma_2^2(w) \approx 2w^\top \Sigma_{\mathrm{PRS}} w.$$

The leading-order SNR becomes

$$\mathrm{SNR}_{\mathrm{LO}}(w) = \frac{(w^\top a_{\mathrm{PRS}})^2}{2w^\top \Sigma_{\mathrm{PRS}} w}.$$

The factor $2$ does not affect the maximizing direction. Therefore, maximizing $\mathrm{SNR}_{\mathrm{LO}}(w)$ is equivalent to maximizing

$$f(w) = \frac{(w^\top a_{\mathrm{PRS}})^2}{w^\top \Sigma_{\mathrm{PRS}} w}.$$

The denominator is positive for any nonzero weight vector, because $\Sigma_{\mathrm{PRS}}$ is positive definite when $|\rho| < 1$.

The objective is scale-invariant, so only the direction of $w$ is identifiable, and the same differentiation argument as in Supplementary Note 1.8 gives $\Sigma_{\mathrm{PRS}} w \propto a_{\mathrm{PRS}}$.

Thus, the leading-order SNR-maximizing direction is

$$w_{\mathrm{LO}}^* \propto \Sigma_{\mathrm{PRS}}^{-1} a_{\mathrm{PRS}}.$$

This has the form of a Fisher linear discriminant[4], with the population covariance matrix of the PRS contrast in place of the within-group covariance: the leading-order optimal direction is the mean-difference vector premultiplied by the inverse of that covariance matrix.

To obtain an explicit closed form, recall that $\Sigma_{\mathrm{PRS}}$ has unit diagonal and off-diagonal entries $-\rho$ (Section 2.5), where $\rho = R_1 R_2 r_{\mathrm{g}}$. Its inverse is

$$\Sigma_{\mathrm{PRS}}^{-1} = \frac{1}{1-\rho^2}\begin{pmatrix} 1 & \rho \\ \rho & 1 \end{pmatrix}.$$

Because the overall scale of $w$ is arbitrary, the factor

$$\frac{1}{1-\rho^2}$$

can be ignored.

Let

$$A = \lambda_1 h_1 - r_\mathrm{g} \lambda_2 h_2,$$

and

$$B = \lambda_2 h_2 - r_\mathrm{g} \lambda_1 h_1.$$

Then

$$a_\mathrm{PRS} = \begin{pmatrix} R_1 A \\ R_2 B \end{pmatrix}.$$

Therefore,

$$w_\mathrm{LO}^* \propto \begin{pmatrix} 1 & \rho \\ \rho & 1 \end{pmatrix} \begin{pmatrix} R_1 A \\ R_2 B \end{pmatrix}.$$

Multiplying the matrix and vector gives an expression in $\rho = R_1 R_2 r_\mathrm{g}$. Substituting this and the definitions of $A$ and $B$ gives, for a common constant $c > 0$,

$$w_1^* = c R_1 \big[\lambda_1 h_1 (1 - R_2^2 r_\mathrm{g}^2) - \lambda_2 h_2 r_\mathrm{g} (1 - R_2^2)\big],$$

and

$$w_2^* = c R_2 \big[\lambda_2 h_2 (1 - R_1^2 r_\mathrm{g}^2) - \lambda_1 h_1 r_\mathrm{g} (1 - R_1^2)\big].$$

Thus, the leading-order optimal PRS discriminant is

$$S_\mathrm{LO} = w_1^* \hat{g}_1 - w_2^* \hat{g}_2,$$

with $w_1^*, w_2^*$ given above up to an arbitrary common positive scale.

This expression shows that finite PRS accuracy changes both the signal direction and the covariance structure.

### 2.10 PRS Genetic Separation Variance

For the leading-order optimal PRS weight,

$$w_\mathrm{LO}^* \propto \Sigma_\mathrm{PRS}^{-1} a_\mathrm{PRS},$$

define the PRS genetic separation variance as

$$V_{\mathrm{PRS}} = a_{\mathrm{PRS}}^{\top} \Sigma_{\mathrm{PRS}}^{-1} a_{\mathrm{PRS}}.$$

Using the canonical scaling

$$w_{\mathrm{LO}}^{*} = \Sigma_{\mathrm{PRS}}^{-1} a_{\mathrm{PRS}},$$

we have

$$\Delta(w_{\mathrm{LO}}^{*}) = (w_{\mathrm{LO}}^{*})^{\top} a_{\mathrm{PRS}} = a_{\mathrm{PRS}}^{\top} \Sigma_{\mathrm{PRS}}^{-1} a_{\mathrm{PRS}} = V_{\mathrm{PRS}}.$$

The marginal variance of the leading-order PRS discriminant is

$$\mathrm{Var}(S_{w_{\mathrm{LO}}^{*}}) = (w_{\mathrm{LO}}^{*})^{\top} \Sigma_{\mathrm{PRS}} w_{\mathrm{LO}}^{*}.$$

Substituting $w_{\mathrm{LO}}^{*} = \Sigma_{\mathrm{PRS}}^{-1} a_{\mathrm{PRS}}$,

$$\mathrm{Var}(S_{w_{\mathrm{LO}}^{*}}) = a_{\mathrm{PRS}}^{\top} \Sigma_{\mathrm{PRS}}^{-1} \Sigma_{\mathrm{PRS}} \Sigma_{\mathrm{PRS}}^{-1} a_{\mathrm{PRS}}.$$

Therefore,

$$\mathrm{Var}(S_{w_{\mathrm{LO}}^{*}}) = a_{\mathrm{PRS}}^{\top} \Sigma_{\mathrm{PRS}}^{-1} a_{\mathrm{PRS}} = V_{\mathrm{PRS}}.$$

At leading order,

$$\sigma_1^2(w_{\mathrm{LO}}^{*}) \approx V_{\mathrm{PRS}},$$

and

$$\sigma_2^2(w_{\mathrm{LO}}^{*}) \approx V_{\mathrm{PRS}}.$$

Thus,

$$\sigma_1^2(w_{\mathrm{LO}}^{*}) + \sigma_2^2(w_{\mathrm{LO}}^{*}) \approx 2V_{\mathrm{PRS}}.$$

The leading-order AUC becomes

$$\mathrm{AUC}_{\mathrm{cc}}^{\mathrm{PRS}} \approx \Phi\left(\frac{V_{\mathrm{PRS}}}{\sqrt{2V_{\mathrm{PRS}}}}\right) = \Phi\left(\sqrt{\frac{V_{\mathrm{PRS}}}{2}}\right).$$

**Closed form of $V_{\mathrm{PRS}}$**

With $A$, $B$, $\rho = R_1 R_2 r_{\mathrm{g}}$ and $\Sigma_{\mathrm{PRS}}^{-1}$ as in Section 2.9, the definition $V_{\mathrm{PRS}} = a_{\mathrm{PRS}}^{\top} \Sigma_{\mathrm{PRS}}^{-1} a_{\mathrm{PRS}}$ gives

$$V_{\mathrm{PRS}} = \frac{R_1^2 A^2 + R_2^2 B^2 + 2\rho R_1 R_2 AB}{1-\rho^2}.$$

Substituting $\rho = R_1 R_2 r_{\mathrm{g}}$, this can also be written as

$$V_{\mathrm{PRS}} = \frac{R_1^2 A^2 + R_2^2 B^2 + 2R_1^2 R_2^2 r_{\mathrm{g}} AB}{1 - R_1^2 R_2^2 r_{\mathrm{g}}^2}.$$

This is the PRS analogue of the TGV genetic separation variance $V_{\mathrm{S}}$.

In the TGV limit $R_1^2 = R_2^2 = 1$, $\rho = r_{\mathrm{g}}$ and $V_{\mathrm{PRS}} = a_{\mathrm{TGV}}^{\mathrm{T}} B_{\mathrm{g}}^{-1} a_{\mathrm{TGV}} = V_{\mathrm{S}}$, so $\mathrm{AUC}_{\mathrm{cc}}^{\mathrm{PRS}}$ reduces to its true-genetic-value counterpart; in the pure-noise limit $R_1^2 = R_2^2 = 0$, $V_{\mathrm{PRS}} = 0$ and $\mathrm{AUC}_{\mathrm{cc}}^{\mathrm{PRS}} = 0.5$. Therefore, $V_{\mathrm{PRS}}$ and $\mathrm{AUC}_{\mathrm{cc}}^{\mathrm{PRS}}$ provide equivalent leading-order summaries of PRS-based subtype separability. Because each PRS estimate is its scaled target genetic value plus independent noise (Section 2.2), the noise can only reduce the separation, so $V_{\mathrm{PRS}} \le V_{\mathrm{S}}$, with equality only when $R_1^2 = R_2^2 = 1$. Under the model, $V_{\mathrm{S}}$ and the corresponding true-genetic-value AUC are therefore upper bounds for $V_{\mathrm{PRS}}$ and the leading-order PRS-based case–case AUC.

### Supplementary Note 3: Detailed Methods

#### 3.1 Software implementation and standard errors

GenSep (C++17, GPL-3.0) takes the GWAS summary statistics of the two subtypes and an LDAK tagging file, and estimates both heritabilities and the genetic correlation with a port of the LDAK SumHer[5,6] solvers. The heritabilities are estimated from each marginal GWAS with the SumHer intercept fixed at 1, and the genetic correlation from the two GWAS jointly with a free cross-trait intercept, which absorbs the correlation between the two sets of summary statistics induced by shared participants (the shared controls and any individual who is a case of both subtypes). The same settings were used for the point estimates and for every jackknife replicate.

Observed-scale heritabilities were converted to the liability scale[3] as

$$h_{i,L}^2 = h_{i,O}^2 \frac{{K_i}^2(1-K_i)^2}{P_i(1-P_i)\phi(t_i)^2},$$

where $K_i$ is the population prevalence adopted for the analysis, $P_i$ is the case fraction of the GWAS from which $h_{i,O}^2$ was estimated, and $t_i = \Phi^{-1}(1-K_i)$. The genetic correlation is unchanged by this rescaling. The liability-scale heritabilities, the genetic correlation and the prevalences then give the derived quantities of Supplementary Notes 1 and 2 (the separation variance, the oracle AUC and its simplified approximation).

For the block jackknife described in the Methods, the SNPs present in both summary-statistic files and the tagging file were split into 200 contiguous blocks with equal numbers of SNPs. In each replicate the heritabilities and the genetic correlation were re-estimated with the settings above, the genetic correlation was truncated to [−0.999, 0.999], and the derived quantities were recomputed. Liability-scale standard errors were the observed-scale ones multiplied by the liability conversion factor. Point estimates were computed from all SNPs.

All analyses were run with the block-jackknife mode of the program as

gensep --se-method jackknife --tagfile <TAGGING> --summary <SUB1> --
summary2 <SUB2> \
--K1 <K1> --K2 <K2> --P1 <P1> --P2 <P2> --num-blocks 200 --out
<PREFIX>

#### 3.2 Cohorts and genetic data

#### 3.2.1 UK Biobank

UK Biobank was the primary cohort: oracle AUCs, internally trained scores and the sample-size projections were estimated here, and FinnGen-trained and consortium-trained scores were also evaluated here. UK Biobank is a population cohort of approximately 500,000 UK residents aged 40–69 years at recruitment[7]. We used the directly genotyped array data. Genotypes were processed with PLINK[8], and individuals with more than 10% missing genotypes were removed. Genetically inferred ancestry groups were taken from UK Biobank Data-Field 30079 (Genomic ancestry)[9], and the European group was used. Related individuals were removed using the KING kinship coefficients released by UK Biobank, which cover all pairs related at third degree or closer (kinship > 0.0442), keeping one member of each related pair. Variant-level quality control then retained autosomal variants with missingness below 5%, minor allele frequency above 1% and Hardy–Weinberg equilibrium $P > 10^{-8}$. The analysis set comprised 357,373 unrelated individuals and 561,997 variants.

The SumHer tagging file was computed with LDAK (--calc-tagging) on 100,000 individuals randomly selected from the analysis set, with uniform predictor weights and $\alpha = -1$ (the GCTA heritability model). The SNP–SNP correlation matrices used by MegaPRS[10] were computed with LDAK (--calc-cors) on 10,000 individuals randomly selected from the analysis set, using the Berisa LD-block breakpoints[11].

Individuals were randomly partitioned once into a training set (80%, $n$ = 285,898) and a held-out test set (20%, $n$ = 71,475). The same partition was used for every subtype pair, for the simulations described in the Methods, and for every polygenic score evaluated in UK Biobank.

#### 3.2.2 All of Us Research Program

All of Us served as an independent cohort for replication, in which oracle AUCs and internally trained scores were estimated for the 18 pairs and FinnGen-trained and consortium-trained scores were also evaluated; it was processed with the same pipeline except where noted below. We used the All of Us Controlled Tier v9 curated data repository[12] and its genotyped array data. Participants were assigned to the European group when the genetic ancestry prediction released by All of Us gave a European probability above 0.95, giving 229,152 participants, of whom the 224,334 with complete covariate data formed the analysis set. Variant-level quality control retained autosomal variants with missingness below 5%, minor allele frequency above 1% and Hardy–Weinberg equilibrium $P > 10^{-8}$, leaving 737,766 variants. The principal components used as covariates (Methods) were those supplied with the release, and age was computed from the recorded date of birth. The SumHer tagging file and the SNP–SNP correlation matrices used by MegaPRS were both computed with LDAK on the same random subset of 50,000 participants of the analysis set, per chromosome and then combined, with the tagging settings used in UK Biobank ($\alpha = -1$, uniform weights). Participants were randomly partitioned once into a training set (80%, $n$ = 179,467) and a held-out test set (20%, $n$ = 44,867), with the constraint that cases and controls of every phenotype were present in both sets.

#### 3.2.3 FinnGen

We used FinnGen data freeze 13[13] (DF13, released publicly in June 2026), a Finnish founder-population resource with no sample overlap with UK Biobank, All of Us or the Million Veteran Program. For each pair we selected the two FinnGen endpoints whose registry definitions most closely match the ICD-10 definitions used in UK Biobank; endpoint identifiers and case and control counts are given in Supplementary Table 1. Summary statistics were harmonized to a common format with the effect allele set to the reference-genome alternate allele and $Z = \beta/\mathrm{SE}$, retaining the total sample size $N = N_{\text{case}} + N_{\text{control}}$, the effective sample size $N_{\text{eff}} = 4N_{\text{case}}N_{\text{control}}/(N_{\text{case}} + N_{\text{control}})$ and the case fraction (cases divided by the total sample size). Each quantity has one use: SumHer was given the total sample size; the liability conversion of Section 3.1 used the case fraction together with the population prevalence $K$ adopted for the pair, the same value as in UK Biobank, so the case fraction did not replace the prevalence; and MegaPRS was given the effective sample size, the usual input for logistic-regression statistics.

Because SumHer requires a tagging file matched to the discovery population, we built a Finnish tagging file. Under the GCTA heritability model the LDAK tagging of a predictor equals its LD score, so we computed HapMap3-restricted LD scores, $\ell_j = 1 + \sum_k r_{jk}^2$, summing over the other HapMap3[14] variants $k \neq j$ within 3 Mb, from the public FinnGen R12 in-sample LD matrices (520,210 Finns), the most recent release with public LD matrices, and converted them into an LDAK tagging file. The R12 matrices are on GRCh38 and list every variant pair within 3 Mb with $r^2 > 0.01$; the resulting LD scores cover 1,200,426 HapMap3 variants, and the tagging file was written under the GCTA heritability model (LDAK power −1), the model of the UK Biobank and All of Us tagging files. The DF13 summary statistics are on GRCh38 and were matched to the rsID-keyed UK Biobank array variants by rsID, so no coordinate conversion was needed (Section 3.7).

#### 3.2.4 Million Veteran Program

We used the openly released Million Veteran Program summary statistics[15] (dbGaP accession phs002453), published per phecode[16] and per genetically inferred ancestry group. Fifteen of the 18 subtype pairs could be mapped to MVP phecodes. The other three (hip versus knee osteoarthritis, internal derangement of the knee versus knee osteoarthritis, and single-episode versus recurrent depression) could not, because the phecodes do not distinguish the ICD-10 categories that define them. Analyses were run in the European, African and Admixed American groups; the pairs analyzed in each group are listed in main-text Table 1.

Case and total counts per phecode and ancestry were taken from the released data dictionary and used to form the case fraction $P$, whereas the population prevalences $K$ were held at the UK Biobank analysis values. MVP reports odds ratios with confidence intervals and $P$ values but no standard errors, so $Z$ was taken as the signed normal quantile of the $P$ value; the total sample size was used. Ancestry-matched tagging files were built from 1000 Genomes reference panels[17], using the phase 3 European (503 individuals), African (AFR, 661) and Admixed American (AMR, 347) samples and biallelic HapMap3 SNPs with minor allele frequency above 1%. Tagging was computed with LDAK with uniform weights and $\alpha = -1$, as in the other cohorts.

#### 3.3 Subtype pairs and phenotype definitions

##### 3.3.1 Selection of the 18 subtype pairs

The 18 clinically defined disease contrasts, referred to as subtype pairs, were selected by three criteria. (i) The two members were clinically distinct entities: either subtypes of a single disease, distinguished by etiology or pathology, anatomical site, disease course or cell lineage, or related conditions of the same organ system that are conventionally classified as separate entities. (ii) Neither member was restricted to one sex. (iii) In UK Biobank, both liability heritabilities were positive and the genetic correlation was finite and away from the ±0.999 estimator boundary. Criteria (i) and (ii) are clinical; criterion (iii) depends on the UK Biobank genetic-parameter estimates and not on polygenic score performance. The basis on which the two members of each pair are distinguished, with one supporting reference per pair, is given in Supplementary Table 1.

Twelve pairs are forms of one disease, distinguished by etiology or pathology (four pairs), disease course (three), anatomical site (four) or cell lineage (one). The remaining six pairs are related conditions of one organ system that are classified as separate diagnoses, including the two residual ICD-10 categories N39 (other disorders of urinary system) and N32 (other disorders of bladder), which are contrasted by anatomical level.

##### 3.3.2 Phenotype definitions in UK Biobank and All of Us

Phenotypes were defined from ICD-10 diagnoses, taken from hospital inpatient records in UK Biobank and from electronic health records in All of Us, with the same rules in both cohorts. Each three-character ICD-10 category was matched together with all of its subcategories. The two record systems differ in coverage, so the same codes need not identify cases of the same severity or with the same probability of being recorded. For each subtype we built a marginal case–control phenotype. These phenotypes were used for the marginal GWAS and for the case–control evaluation of each score, and their cases form the case–case evaluation groups of Section 3.5.2.

Controls were defined operationally as participants with no record of the relevant codes, not as participants known to be free of disease. For 13 pairs, controls are participants with no record of either subtype. For the other five, controls also exclude the surrounding disease block: E10–E14 for diabetes, F30–F39 for depression, I60–I64 for stroke, M15–M19 for osteoarthritis and E00–E07 for thyroid disease. Participants with none of these codes, including those with no hospital or electronic health record, were controls.

For four pairs, we imposed mutually exclusive case definitions (type 1 versus type 2 diabetes, Crohn disease versus ulcerative colitis, ischemic versus hemorrhagic stroke and hypothyroidism versus hyperthyroidism): individuals with both subtypes were removed from the cases, as were diabetes cases with any other diabetes code (E12–E14) and stroke cases with an unspecified stroke code (I64). In the other 14 pairs, individuals with both subtypes were kept in both marginal phenotypes. Supplementary Table 1 gives the ICD-10 codes, the handling of individuals with both subtypes and the case counts for each pair.

#### 3.3.3 Assumed prevalences and sensitivity to them

We specified a population prevalence $K$ for each subtype based on published estimates. The values used, their sources and any adjustments are listed in Supplementary Table 1.

To assess sensitivity to these assumptions, we halved or doubled the prevalence of either subtype or both subtypes in UK Biobank, giving six perturbations per pair and 108 recomputations across the 18 pairs. We held the observed-scale heritabilities, GWAS case fractions and genetic correlation fixed, recalculated the liability-scale heritabilities using the conversion in Section 3.1, and recomputed the oracle AUC. The AUC changed by a median of 0.002 and at most 0.018, the latter for inguinal versus diaphragmatic hernia when both prevalences were doubled. For 17 of the 18 pairs, the largest change was smaller than 1.96 times the baseline jackknife s.e.; the maximum change for the remaining pair was 2.1 s.e. (Supplementary Fig. 10).

The modest sensitivity arises because the effects of $K$ on selection intensity and liability-scale heritability largely cancel in the prevalence-weighted genetic standard deviation:

$$\lambda_i h_{i,L} = (1 - K_i)\sqrt{\frac{h_{i,O}^2}{P_i(1 - P_i)}}.$$

Thus, with the observed-scale estimates and case fractions fixed, the separation variance $V_S$ depends on the assumed prevalences only through $1 - K_i$, which changes little at low prevalences. The moment-based oracle AUC additionally depends on prevalence through its truncation correction. The estimates were therefore robust to the tested prevalence changes in UK Biobank. We used the same $K$ values across cohorts and ancestry groups to provide a common reference for comparison; the resulting estimates are not calibrated to each target population's own prevalence.

#### 3.4 Association testing and estimation of the genetic ceiling

##### 3.4.1 Genome-wide association analyses

Within UK Biobank and All of Us, association testing used ldak --linear, a linear model on the 0/1 phenotype adjusted for sex, age, age$^2$, age$^3$, the interactions sex × age, sex × age$^2$ and sex × age$^3$, and the first 20 (UK Biobank) or 10 (All of Us) genetic principal components. Two GWAS were run per pair: subtype 1 versus controls and subtype 2 versus controls. Two sets of GWAS served two purposes. We used full-cohort GWAS to estimate heritabilities, genetic correlations and the oracle quantities. For internally trained polygenic scores, GWAS were restricted to the 80% training set, and every score was evaluated in the held-out 20% test set, using the same training–test partition for all subtype pairs.

##### 3.4.2 Reliability rule

Within each cohort, a subtype pair is considered reportable (reliable in Supplementary Table 2) if (i) both liability-scale heritability estimates are positive, (ii) the cross-subtype genetic correlation estimate is finite and does not lie on either estimator boundary (±0.999) and (iii) the derived quantities (the separation variance and the oracle AUC) are defined. A liability-scale heritability above 1 is recorded as a flag but does not make a pair unreliable; because the model of Supplementary Note 1 requires $h_i^2 \leq 1$, such an estimate lies outside the parameter range, and its oracle AUC should be read together with the sensitivity analysis of Section 3.4.3. The reliability rule governs which estimates are reported and compared; it is distinct from the parameter range of the model. All cross-cohort correlations and per-pair difference tests are restricted to pairs meeting these criteria in both analyses being compared.

Supplementary Table 2 reports each pair's reliability status and flags.

#### 3.4.3 Type 1 versus type 2 diabetes across cohorts and definitions

Estimates of genetic separability between type 1 and type 2 diabetes varied across cohorts, suggesting a contribution of phenotype definition. In the primary analyses, the estimated oracle AUC was 1.000 in UK Biobank, 0.999 in All of Us and 0.966 in FinnGen (T1D_EARLY), with corresponding cross-subtype genetic correlations of 0.04, −0.03 and −0.07. In MVP, where the subtypes were defined using phecodes 250.1 and 250.2, the estimated oracle AUCs were lower across all three ancestry groups: 0.605 in European, 0.641 in African and 0.655 in Admixed American participants, with corresponding genetic correlations of 0.95, 0.95 and 0.78 (Supplementary Table 2).

To assess sensitivity to phenotype definition, we compared seven cohort–definition combinations: FinnGen T1D_EARLY (2,960 type 1 diabetes cases) and T1D_WIDE (11,197); MVP phecode 250.1 in the European ancestry analysis (16,971); and strict and broad definitions in UK Biobank and All of Us. The strict type 1 diabetes definition required an E10 code without E11 or E12–E14 codes, yielding 688 cases in UK Biobank and 364 in All of Us. The broad definition included any E10 code, retaining individuals with an E11 code, and yielded 3,502 and 2,883 cases, respectively; the type 2 diabetes definition was also broadened to include any E11 code. Applying these alternative definitions within the same underlying cohort lowered the estimated oracle AUC from 1.000 to 0.871 in UK Biobank and from 0.999 to 0.890 in All of Us, while the genetic correlation increased from 0.04 to 0.56 and from −0.03 to 0.43, respectively. The seven combinations differ in cohort, record system, sample size and definition at the same time, so they are compared as combinations and not as an ordering of diagnostic stringency; the within-cohort comparisons, which change only the definition, support a contribution of phenotype definition to the observed differences in genetic separability (Supplementary Fig. 13b, Supplementary Table 6).

The estimated liability-scale heritability of type 1 diabetes exceeded 1 in UK Biobank and All of Us (1.38 ± 0.42 and 1.15 ± 0.64, respectively; estimate ± s.e.). The observed-scale estimates behind them are unremarkable (0.024 and 0.018); the liability-scale values exceed 1 because the conversion uses the population prevalence $K = 0.005$ against sample case fractions of only 0.21% and 0.19%, respectively, so that a small observed-scale sampling error is multiplied by a large factor, not because the estimation itself failed. These estimates have substantial uncertainty, and the corresponding oracle AUC estimates close to 1 should be interpreted cautiously. To assess sensitivity to the type 1 diabetes heritability estimate, we varied its value from approximately 0.149 (the MVP estimate) to 1.377 (the UK Biobank estimate), holding the type 2 diabetes heritability, genetic correlation and both prevalences at their UK Biobank values. Across this approximately ninefold range, the estimated oracle AUC ranged from 0.88 to 1.00 between the two anchor values; the curve in Supplementary Fig. 13c is drawn over the wider grid 0.05 to 1.45.

#### 3.5 Polygenic scores and observed case–case discrimination

##### 3.5.1 Score accuracy on the genetic scale

Genetic score accuracy is defined as $R_i^2 = \mathrm{Cor}(\hat{g}_i, g_i)^2$, where $\hat{g}_i$ is the polygenic score and $g_i$ is the standardized genetic value (Supplementary Note 2). In real data, we estimated $R_i^2 = r_{iL}^2/h_i^2$, where $r_{iL}^2 = \mathrm{Cor}(\hat{g}_i, L_i)^2$ was inferred from the score's case–control AUC and $h_i^2$ is the liability-scale SNP heritability, assuming that the score is uncorrelated with the non-genetic component of liability.

Following Wray et al.[18], we used a normal approximation to the score distributions in cases ($L > t$) and controls ($L < t$). For a single subtype, omitting the subtype index, the inversion is

$$r_L^2 = \frac{2z^2}{m^2 + z^2 S}, \qquad z = \Phi^{-1}(\mathrm{AUC}),$$

where $\phi$ and $\Phi$ are the standard normal density and distribution function, $t = \Phi^{-1}(1-K)$ is the liability threshold at population prevalence $K$, $m = \phi(t)/[K(1-K)]$, and

$$S = i_{\mathrm{case}}(i_{\mathrm{case}} - t) + i_{\mathrm{ctrl}}(i_{\mathrm{ctrl}} - t), \qquad i_{\mathrm{case}} = \frac{\phi(t)}{K}, \qquad i_{\mathrm{ctrl}} = -\frac{\phi(t)}{1-K}.$$

##### 3.5.2 Observed case–case AUC

We evaluated case–case discrimination using

$$D = w_1^{\mathrm{PRS}} z(\hat{g}_1) - w_2^{\mathrm{PRS}} z(\hat{g}_2),$$

where $z(\cdot)$ standardizes each score over all test individuals. The leading-order optimal weights $w_i^{\mathrm{PRS}}$ were computed from the estimated liability heritabilities, genetic correlation, population prevalences and score accuracies $R_i^2$ (Supplementary Note 2.9; Section 3.5.1). Subtype-1 and subtype-2 cases formed the positive and negative classes, respectively, following the case definitions in Section 3.3.2.

We used two-fold cross-fitting to estimate and evaluate the weights. Test participants were split into two halves using a fixed seed shared across cohorts and simulations.

Scores were oriented, accuracies estimated and weights formed in one half, and $D$ was evaluated in the other. After exchanging the halves, we averaged the two AUCs with weights proportional to the number of case pairs in each evaluation half.

An alternative weighting was evaluated on the same halves: a weight direction selected to maximize case–case AUC in the weight half by searching the weight-vector angle in 1° steps.

Internally trained, FinnGen-trained and consortium-trained scores were compared in the same held-out individuals scored by all available sources for each pair, using identical splits and the same weighting procedure. In All of Us, weights used the cohort's own heritability and genetic correlation estimates. UK Biobank estimates were substituted for ischemic versus hemorrhagic stroke and gastric versus duodenal ulcer because their All of Us genetic correlations could not be estimated; no model-predicted PRS AUC was reported for these pairs. Supplementary Table 3 reports the primary cross-fitted AUCs and the alternative weighting; standard errors refer to the primary cross-fitted estimates.

We compared observed AUCs with the moment-based predictions of Supplementary Note 2.7 using Pearson correlation, the calibration slope from an unweighted regression of observed on predicted AUC with an intercept, and mean absolute error (Supplementary Fig. 11). Predictions used the estimated genetic parameters, prevalences and score accuracies, with standardized-score covariance $\rho = R_1 R_2 r_g$.

For empirical separation recovery, we converted the observed AUC to $V_{PRS} = 2[\Phi^{-1}(\mathrm{AUC_{PRS}})]^2$ and divided by the target cohort's own $V_S$, clipping the ratio to [0,1]. Recovery was left missing when the observed AUC was at or below 0.5. Empirical AUC recovery, the recovery measure reported in the main text and Supplementary Table 3, was computed directly from the observed AUC and the target cohort's own analytical oracle AUC (Section 3.5.3), without clipping.

#### 3.5.3 Separation recovery and AUC recovery

Separation recovery, $f = V_{PRS}/V_S$, is the proportion of the genetic separation variance captured by the score (Section 3.5.2). AUC recovery is the proportion of the above-chance oracle AUC achieved by the score:

$$f_{\mathrm{AUC}} = \frac{\mathrm{AUC_{PRS}} - 0.5}{\mathrm{AUC_{oracle}} - 0.5}.$$

Under the simplified approximation, $\mathrm{AUC_{oracle}} = \Phi\left(\sqrt{\frac{V_S}{2}}\right)$ and $\mathrm{AUC_{PRS}} =$

$\Phi\left(\sqrt{\frac{V_{\mathrm{PRS}}}{2}}\right)$. Substituting these and using $\Phi(x)-\frac{1}{2}=\frac{1}{2}\mathrm{erf}(x/\sqrt{2})$, where $\mathrm{erf}(z)=\frac{2}{\sqrt{\pi}}\int_0^z e^{-t^2}\mathrm{d}t$ is the error function, gives a closed form that involves no further approximation:

$$f_{\mathrm{AUC}}=\frac{\mathrm{erf}\left(\frac{\sqrt{V_{\mathrm{PRS}}}}{2}\right)}{\mathrm{erf}\left(\frac{\sqrt{V_{\mathrm{S}}}}{2}\right)}=\frac{\mathrm{erf}\left(\frac{\sqrt{f}\sqrt{V_{\mathrm{S}}}}{2}\right)}{\mathrm{erf}\left(\frac{\sqrt{V_{\mathrm{S}}}}{2}\right)}.$$

AUC recovery is therefore a function of both the separation recovery and the ceiling $V_{\mathrm{S}}$, not of $f$ alone. Conversely, under the same approximation, separation recovery is the squared ratio of the two AUCs on the probit scale:

$$f=\frac{V_{\mathrm{PRS}}}{V_{\mathrm{S}}}=\left(\frac{\Phi^{-1}(\mathrm{AUC}_{\mathrm{PRS}})}{\Phi^{-1}(\mathrm{AUC}_{\mathrm{oracle}})}\right)^2.$$

Expanding the numerator in a Taylor series about zero, $\mathrm{erf}(z)=\frac{2}{\sqrt{\pi}}\left(z-\frac{z^3}{3}+\cdots\right)$, with $z=\sqrt{f}\sqrt{V_{\mathrm{S}}}/2$, gives for small $f$:

$$f_{\mathrm{AUC}}\approx\sqrt{f}\kappa(V_{\mathrm{S}})\left(1-\frac{fV_{\mathrm{S}}}{12}\right),\kappa(V_{\mathrm{S}})=\frac{\sqrt{\frac{V_{\mathrm{S}}}{\pi}}}{\mathrm{erf}\left(\frac{\sqrt{V_{\mathrm{S}}}}{2}\right)}\approx 1+\frac{V_{\mathrm{S}}}{12}.$$

For small $f$, $f_{\mathrm{AUC}}\approx\sqrt{f}\kappa(V_{\mathrm{S}}),\kappa(V_{\mathrm{S}})\geq 1$.

In UK Biobank, AUC recovery followed $\sqrt{f}$ closely for the 16 pairs with an oracle AUC of 0.822 or below ($f_{\mathrm{AUC}}/\sqrt{f}$ between 0.97 and 1.11) but exceeded it for the two pairs with the highest ceilings, hypothyroidism versus hyperthyroidism and type 1 versus type 2 diabetes (Supplementary Fig. 18a). Multiplying $\sqrt{f}$ by $\kappa(V_{\mathrm{S}})$ brought all 18 pairs onto the identity line, reducing the mean absolute error from 0.023 to 0.009 and the largest deviation from 0.23 to 0.05 (Supplementary Fig. 18b). The remaining differences reflect the higher-order term $(1-fV_{\mathrm{S}}/12)$ and the truncation correction retained in the reported oracle AUC (main-text equation (2)).

#### 3.6 Projection of case–case AUC against training sample size

#### 3.6.1 From training sample size to the recovered fraction

We projected the case–case AUC against the number of training cases per subtype. The projection rests on the standard relationship between polygenic-score accuracy and training sample size[19–21]: under the infinitesimal model, for the squared correlation $R^2$ between a PRS and the true genetic liability, $1/R^2 - 1$ is proportional to $1/N_{\mathrm{eff}}$, where $N_{\mathrm{eff}} = 4N_{\mathrm{case}}N_{\mathrm{ctrl}}/(N_{\mathrm{case}} + N_{\mathrm{ctrl}})$ is the effective discovery sample size. We applied this relationship to each subtype's score separately. Let $R_i^2$ denote the accuracy of the score for subtype $i$ on the genetic scale (Section 3.5.1) and $\kappa_i$ the factor by which the effective sample size of arm $i$ is multiplied. Because multiplying $N_{\mathrm{eff}}$ by $\kappa_i$ divides $1/R_i^2 - 1$ by $\kappa_i$,

$$R_i^2(\kappa_i) = \frac{\kappa_i R_{i,\mathrm{curr}}^2}{1 - R_{i,\mathrm{curr}}^2 + \kappa_i R_{i,\mathrm{curr}}^2},$$

where the current accuracy $R_{i,\mathrm{curr}}^2$ was obtained from the case–control AUC of each subtype's score (Section 3.5.1).

The two extrapolated accuracies were recombined through the simplified-approximation expressions for the observed case–case score. Under the simplified approximation, the leading-order optimal combination of the two scores has separation variance

$$V_{\mathrm{PRS}} = a_{\mathrm{PRS}}^{\top} \Sigma_{\mathrm{PRS}}^{-1} a_{\mathrm{PRS}},$$

which depends on the score accuracies $R_i^2$, the prevalence-weighted genetic standard deviations $\lambda_i h_i$ and the genetic correlation $r_{\mathrm{g}}$ (Supplementary Notes 2.9–2.10). Substituting $R_i^2(\kappa_i)$ gives $V_{\mathrm{PRS}}(\kappa_1, \kappa_2)$ and the recovered fraction

$$f(\kappa_1, \kappa_2) = \frac{V_{\mathrm{PRS}}(\kappa_1, \kappa_2)}{V_{\mathrm{S}}},$$

which rises to 1 as both $\kappa_i$ grow. The fraction is mapped to the AUC scale through the oracle ceiling $V_{\mathrm{ceiling}} = 2[\Phi^{-1}(\mathrm{AUC}_{\mathrm{oracle}})]^2$, where $\mathrm{AUC}_{\mathrm{oracle}}$ is the analytical oracle AUC reported for the pair:

$$\mathrm{AUC}(\kappa_1, \kappa_2) = \Phi\left(\sqrt{\frac{f(\kappa_1, \kappa_2) V_{\mathrm{ceiling}}}{2}}\right).$$

Every projected curve therefore asymptotes at the oracle AUC reported for that pair. Together with the accuracy expression above, this is main-text equation (6).

#### 3.6.2 Calibration, projection scenarios and target sample sizes

Because the current accuracies are phenotype-based proxies, they fix the relative signal-to-noise levels of the two arms, but substituting them directly ($\kappa_1 = \kappa_2 = 1$) need not reproduce the observed case–case AUC. We therefore calibrated a common scaling factor $\kappa_0$ to anchor each curve at its observed AUC. Defining

$$f_{\text{curr}} = \frac{2[\Phi^{-1}(\text{AUC}_{\text{PRS}})]^2}{V_{\text{ceiling}}},$$

we selected $\kappa_0$ such that $f(\kappa_0, \kappa_0) = f_{\text{curr}}$, found by one-dimensional numerical root finding. A sample-size multiple $s_i$ relative to the current training size of arm $i$ then corresponds to $\kappa_i = \kappa_0 s_i$, with projected AUC

$$\text{AUC}(s_1, s_2) = \Phi\left(\sqrt{\frac{f(\kappa_0 s_1, \kappa_0 s_2)\, V_{\text{ceiling}}}{2}}\right).$$

Each curve therefore passes through the observed AUC at $s_1 = s_2 = 1$ and approaches the oracle AUC as both arms grow. The calibration target $f_{\text{curr}}$ differs from the empirical recovery in Section 3.5.2 whenever $V_{\text{ceiling}} \neq V_{\text{S}}$; the theoretical function $f = V_{\text{PRS}}/V_{\text{S}}$ is unchanged.

The main projection scales both arms by the same factor, $s_1 = s_2 = N/N_{\text{curr}}$ (so that $\kappa_1 = \kappa_2$), and assumes an unlimited supply of controls, so that effective sample size grows in proportion to case count. Here $N_{\text{curr}}$ is the total of the two subtypes' case counts in the 80% training split; $N$ on the horizontal axis of Fig. 3b is thus the total training case count of the two arms, and $\text{AUC}(N)$ in main-text equation (6) equals $\text{AUC}(N/N_{\text{curr}}, N/N_{\text{curr}})$.

The number of cases required to reach a target AUC was obtained by solving

$$f(\kappa_1, \kappa_2) = f_{\text{target}}, \qquad f_{\text{target}} = \frac{2\left[\Phi^{-1}(\text{AUC}_{\text{target}})\right]^2}{V_{\text{ceiling}}}$$

numerically by one-dimensional root finding. Targets above the oracle ceiling ($f_{\text{target}} > 1$) were classified as unreachable. We also projected growth in each arm separately, holding the other at its current size ($\kappa_j = \kappa_0$), to identify which arm offered the greater projected gain. In this scenario a target was unreachable if the limiting value of $f$ remained below $f_{\text{target}}$; limits were evaluated up to $\kappa = 10^{12}$.

Supplementary Table 4 reports calibration factors, training case counts, and projected counts for each arm and their total, and classifies each target as already attained, attainable at a finite size, above the oracle ceiling, or unattainable with the other arm fixed. Because the projection holds the genetic architecture fixed and is calibrated to one observed AUC per pair, the required sample sizes should be interpreted as order-of-magnitude estimates rather than as the output of a power calculation.

#### 3.7 External discovery GWAS and transfer analyses

FinnGen summary statistics were aligned to target-cohort variants by rsID in UK Biobank, requiring no genome-build conversion, and by chromosome–position–reference–alternate identifier in All of Us. Alleles were checked, effects were reoriented to the target A1 allele, and unresolved strand-ambiguous variants were excluded. For each jointly analyzed pair, both scores used the intersection of the two subtypes' variant sets. MegaPRS used the target cohort's LD correlation matrices, and scores were evaluated under the protocol in Section 3.5.2. Consortium summary statistics were processed in the same way.

The thyroid comparison additionally involved different phenotype definitions. UK Biobank excluded individuals with both E03 and E05 codes, whereas the consortium meta-analyses defined hypothyroidism and hyperthyroidism independently, allowing individuals to be cases in both discovery GWAS. In UK Biobank, 50.3% of E05 cases also carried E03, potentially reflecting hypothyroidism following definitive treatment of Graves disease. The external genetic correlation was 0.675, compared with 0.217 internally. This definition difference should therefore be considered when interpreting the thyroid transfer results.

#### 3.8 Effect of subtype misclassification on the observed case–case AUC

Let $D$ be a fixed case–case score and let $A = P(D_1 > D_2)$ be its AUC between correctly labeled cases, where $D_1$ and $D_2$ are the scores of independently drawn true subtype-1 and true subtype-2 cases. What is computed in practice is instead the observed AUC, $A_{\text{obs}} = P(\widetilde{D}_1 > \widetilde{D}_2)$, where $\widetilde{D}_1$ and $\widetilde{D}_2$ are the scores of independently drawn cases recorded as subtype 1 and subtype 2. To relate the two, suppose that a fraction $\varepsilon_1$ of the cases recorded as subtype 1 are in truth subtype 2, and a fraction $\varepsilon_2$ of the cases recorded as subtype 2 are in truth subtype 1. Here $\varepsilon_i$ is the fraction of the cases recorded as subtype $i$ that belong to the other true subtype. It is not a false-negative rate or the probability that a true case is mislabeled. We assume that mislabeling is independent of the score within each true subtype, so that a mislabeled case has the score distribution of its true subtype.

Under this assumption, each recorded group is a mixture of the two true subtypes. A recorded subtype-1 case is a true subtype-1 case with probability $1 - \varepsilon_1$ and a true subtype-2 case with probability $\varepsilon_1$. A recorded subtype-2 case is a true subtype-2 case with probability $1 - \varepsilon_2$ and a true subtype-1 case with probability $\varepsilon_2$. Because the two cases in a pair are drawn independently, the pair of true subtypes behind a recorded (subtype-1, subtype-2) pair takes one of four configurations: both labels correct, with probability $(1-\varepsilon_1)(1-\varepsilon_2)$; both labels wrong, with probability $\varepsilon_1\varepsilon_2$; both cases truly subtype 1, with probability $(1-\varepsilon_1)\varepsilon_2$; or both cases truly subtype 2, with probability $\varepsilon_1(1-\varepsilon_2)$.

Within each configuration, the probability that the recorded subtype-1 case scores higher follows from the true subtypes alone. When both labels are correct, the comparison is between a true subtype-1 and a true subtype-2 case, so the probability is $A$. When both labels are wrong, the comparison is reversed and the probability is $1-A$. When both cases belong to the same true subtype, they are exchangeable, so the probability is 1/2 for a continuous score. Ties, if present, count one half under the usual AUC definition.

Averaging these conditional probabilities over the four configurations gives the observed AUC:

$$A_{\text{obs}} = (1-\varepsilon_1)(1-\varepsilon_2)A + \varepsilon_1\varepsilon_2(1-A) + \frac{1}{2}[(1-\varepsilon_1)\varepsilon_2 + \varepsilon_1(1-\varepsilon_2)].$$

Therefore, the observed AUC is

$$A_{\text{obs}} = (1-\varepsilon_1-\varepsilon_2)A + \frac{\varepsilon_1+\varepsilon_2}{2},$$

which reduces to $A_{\text{obs}} = (1-2\varepsilon)A + \varepsilon$ when $\varepsilon_1 = \varepsilon_2 = \varepsilon$.

Subtracting 1/2 from both sides shows what this relation does to the discriminative signal:

$$A_{\text{obs}} - \frac{1}{2} = (1-\varepsilon_1-\varepsilon_2)\left(A - \frac{1}{2}\right).$$

The distance of the AUC from 0.5 is shrunk by the factor $1-\varepsilon_1-\varepsilon_2$. Because mislabeled cases are assumed to be a minority of each recorded group $(\varepsilon_1, \varepsilon_2 < 1/2)$, this factor lies between 0 and 1, so the observed AUC is always pulled towards 0.5. For $\varepsilon = 0.05$, an AUC of 0.80 is observed as 0.77 and an AUC of 0.90 as 0.86. When the error fractions are known, the same relation can be inverted to recover the true AUC:

$$A = \frac{1}{2} + \frac{A_{\text{obs}} - 1/2}{1-\varepsilon_1-\varepsilon_2}.$$

### Supplementary Figures

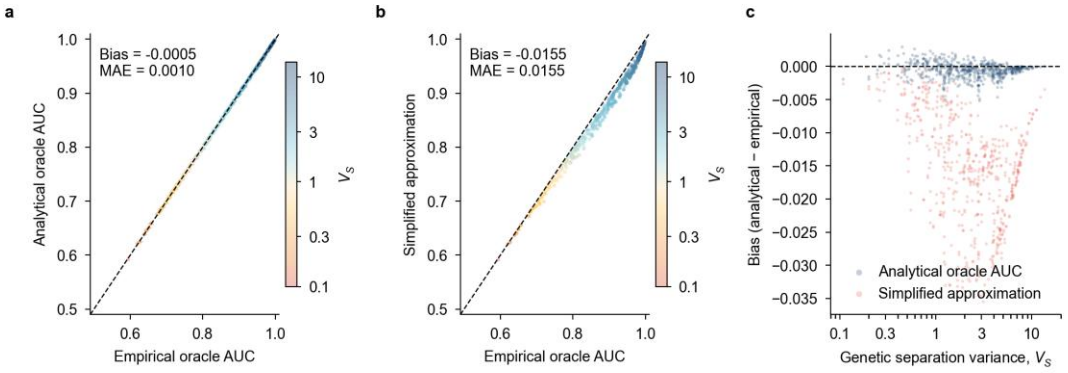


**Supplementary Fig. 1: Validation of the analytical oracle AUC in simulations.**

**a,** Analytical oracle AUC plotted against empirical oracle AUC. **b,** Simplified approximation plotted against empirical oracle AUC. **c,** Bias, defined as the analytical or simplified AUC minus the empirical AUC, as a function of the genetic separation variance, $V_S$ (log scale); blue and pink points denote the analytical oracle AUC and simplified approximation, respectively. Each point represents one of 576 genetic-architecture settings with zero residual correlation. Empirical AUCs are means over 30 replicates of 5,000 cases per subtype; the optimal genetic discriminant weight was estimated by sample-split grid search and evaluated in held-out cases. Main-text Fig. 2a instead evaluates the empirical AUC at the analytical weights, which gives its smaller mean absolute error. Horizontal error bars in **a** denote ±1 s.d. across replicates. Points in **a**,**b** are colored by $V_S$. Dashed lines denote identity (**a,b**) or zero bias (**c**); bias and mean absolute error (MAE) summarize agreement across settings. No statistical tests were performed.

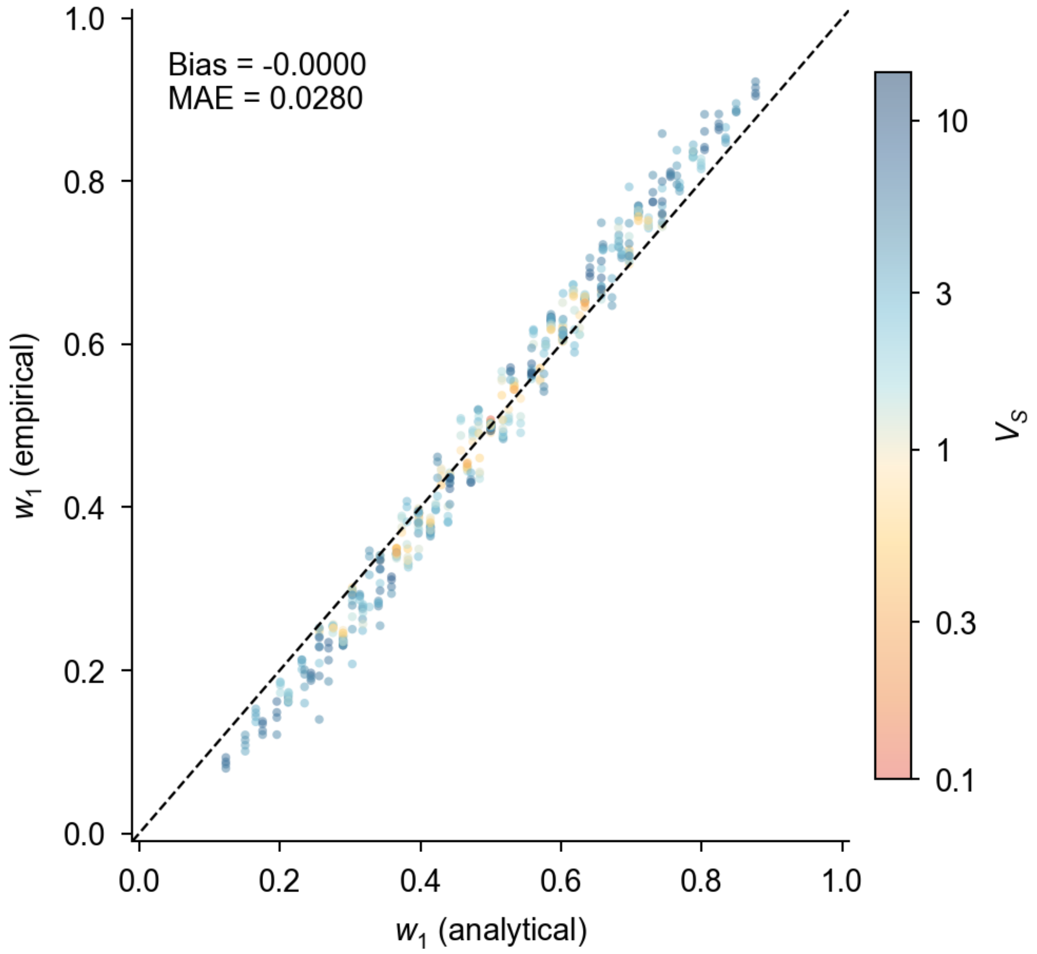


**Supplementary Fig. 2: Validation of the optimal genetic discriminant weight in simulations.**

Empirically estimated weight $w_1$ for the subtype-1 standardized genetic value in the oracle discriminant $w_1 g_1 - (1 - w_1) g_2$, plotted against its analytical value $\frac{\lambda_1 h_1}{\lambda_1 h_1 + \lambda_2 h_2}$; the two weights are normalized to sum to one. Empirical weights were obtained by sample-split grid search in simulated case sets. Each point represents one of 576 genetic-architecture settings, averaged over 30 replicates, and is colored by the genetic separation variance, $V_S$. The dashed line denotes identity. Bias is the mean empirical minus analytical weight, and MAE is the mean absolute error across settings. No statistical tests were performed.

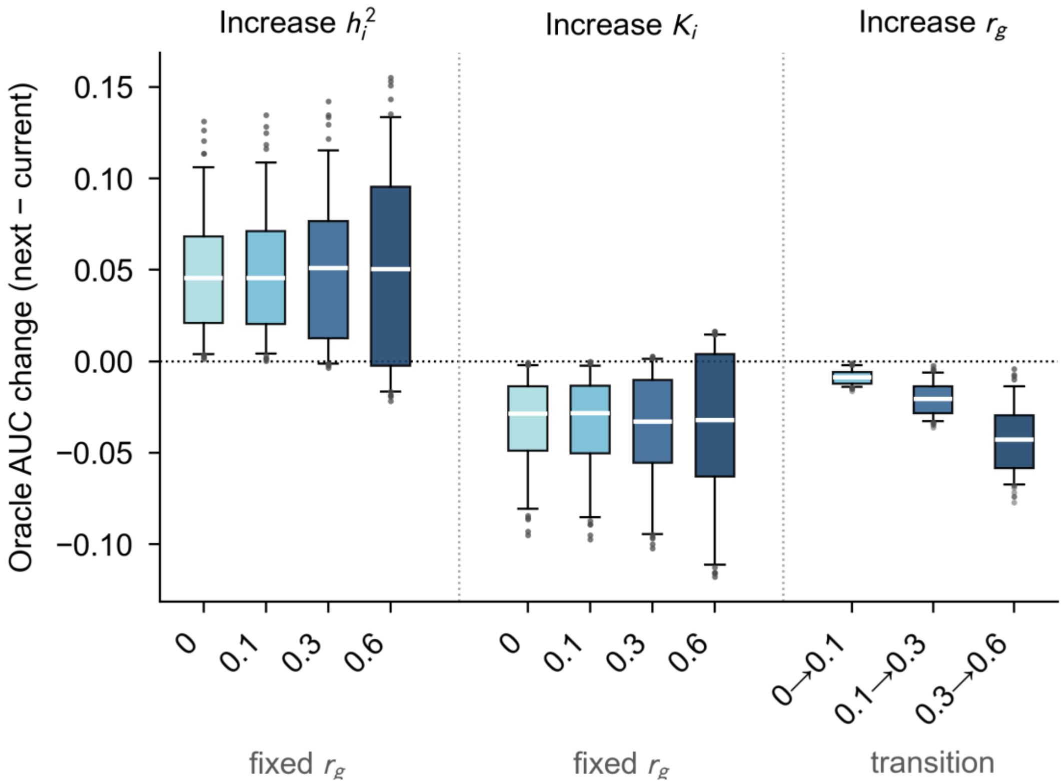


**Supplementary Fig. 3: Changes in oracle AUC across adjacent genetic-architecture levels.**

Change in analytical oracle AUC after increasing one parameter to the next simulation-grid level while holding all other parameters fixed. The plotted quantity is $AUC_{oracle}$(next) − $AUC_{oracle}$(current). The subtype heritability ($h^2$) and prevalence ($K$) groups pool changes to either subtype and all adjacent transitions, stratified by the fixed cross-subtype genetic correlation, $r_g$. The $r_g$ group shows the three adjacent transitions separately. Shading runs from light to dark with the fixed $r_g$ level in the first two groups and the destination $r_g$ level in the third. Boxes show interquartile ranges, white lines mark medians, whiskers extend to the 5th and 95th percentiles, and points denote values outside this range. Each box summarizes 192 ($h^2$), 216 ($K$) or 144 ($r_g$) one-step changes. The dotted horizontal line denotes zero change. Boxes summarize variation across the parameter grid; no statistical tests were performed.

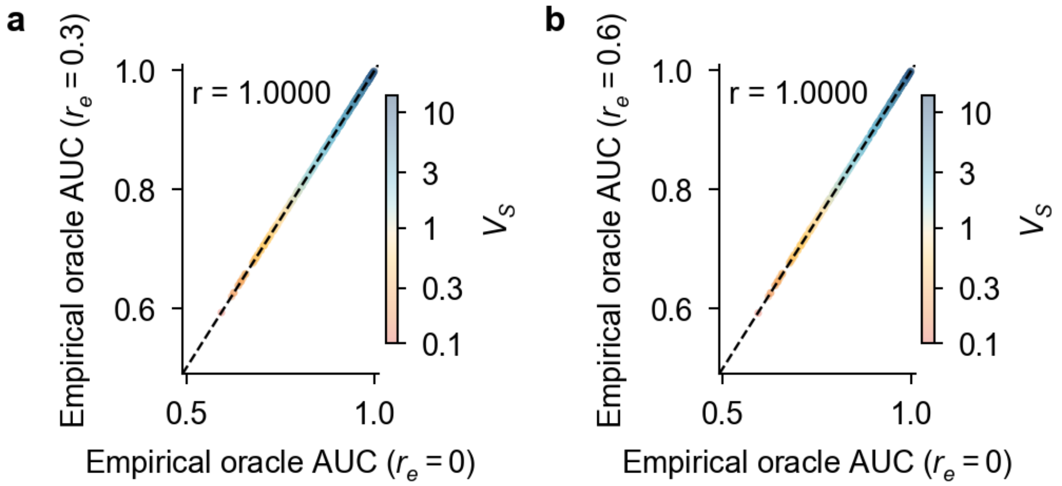


**Supplementary Fig. 4: Sensitivity of oracle AUC to residual correlation between subtype liabilities.**

Empirical oracle AUC at residual correlation **a,** $r_e = 0.3$ or **b,** $r_e = 0.6$ plotted against the corresponding AUC at $r_e = 0$, with subtype heritabilities, prevalences and genetic correlation held fixed. AUCs were evaluated using the analytical optimal genetic discriminant weights. Each panel contains 576 paired genetic-architecture settings, with empirical AUCs averaged over 30 replicates. Settings differing only in $r_e$ used common random numbers. Points are colored by the genetic separation variance, $V_S$, and dashed lines denote identity. The Pearson correlations are descriptive and were not tested. The comparison covers the non-negative residual correlations in the simulation grid.

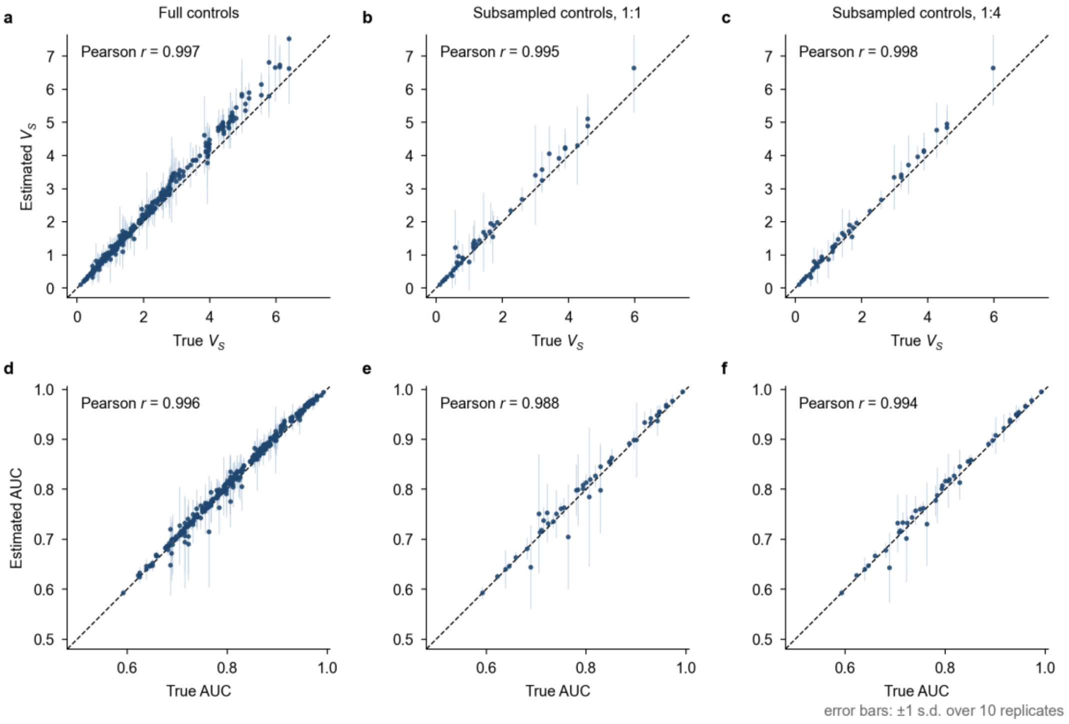


**Supplementary Fig. 5: Estimation of oracle quantities from simulated GWAS summary statistics.**

GenSep estimates plotted against values calculated from the prespecified simulation parameters. Estimates were obtained by substituting SumHer estimates of liability-scale subtype SNP heritabilities and genetic correlation, together with the known subtype prevalences, into the analytical expressions. Top row, genetic separation variance, $V_{\mathrm{S}}$ (**a**–**c**); bottom row, oracle AUC (**d**–**f**). Columns show full controls (all eligible controls; a,d), 1:1 case–control sampling (**b**,**e**) and 1:4 case–control sampling (**c**,**f**). Each point represents one parameter setting averaged over ten replicates: the full simulation grid of 324 settings for full controls (**a**,**d**) and a sensitivity subset of 45 settings (15 heritability–prevalence combinations, nine symmetric and six randomly chosen asymmetric, at three genetic-correlation levels) for the subsampled designs (**b**,**c**,**e**,**f**); vertical error bars denote ±1 s.d. across replicates. Dashed lines denote identity. Each panel reports the Pearson correlation across settings; these correlations are descriptive and were not tested. On the 45 settings shared by the three designs (450 matched replicates), the mean absolute error of the estimated oracle AUC across setting means was 0.010, 0.012 and 0.010 and the Pearson correlation 0.995, 0.988 and 0.994 for full controls, 1:1 and 1:4 sampling, with finite estimates in 447, 442 and 446 of the 450 replicates; the corresponding correlations for $V_{\mathrm{S}}$ were 0.999, 0.995 and 0.998.

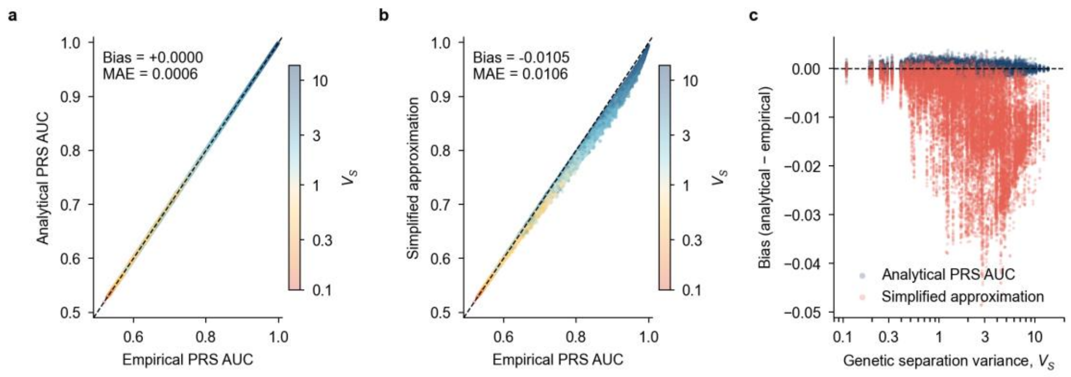


**Supplementary Fig. 6: Validation of the analytical PRS-based case–case AUC in simulations.**

**a,** Analytical PRS case–case AUC plotted against empirical AUC. **b,** Simplified approximation plotted against empirical AUC. **c,** Bias, defined as the analytical or simplified AUC minus the empirical AUC, as a function of the genetic separation variance, $V_S$ (log scale); blue and pink points denote the analytical formula and simplified approximation, respectively. Each point represents one of 20,736 combinations of subtype liability-scale SNP heritabilities, prevalences, genetic correlation and PRS accuracies. Empirical AUCs were evaluated in the full simulated case set at the fixed leading-order optimal angle $\theta_{lo}$ defined in Supplementary Fig. 7, the weights used for every prediction in the manuscript, and averaged over 30 replicates. Panels **a** and **b** therefore compare the moment-based approximation and the simplified approximation with the empirical AUC at one and the same weight rule, so that each panel isolates the error of its formula. Horizontal error bars in **a** denote ±1 s.d. across replicates. Points in **a**,**b** are colored by $V_S$. Dashed lines denote identity (**a,b**) or zero bias (**c**); MAE denotes mean absolute error across settings. No statistical tests were performed.

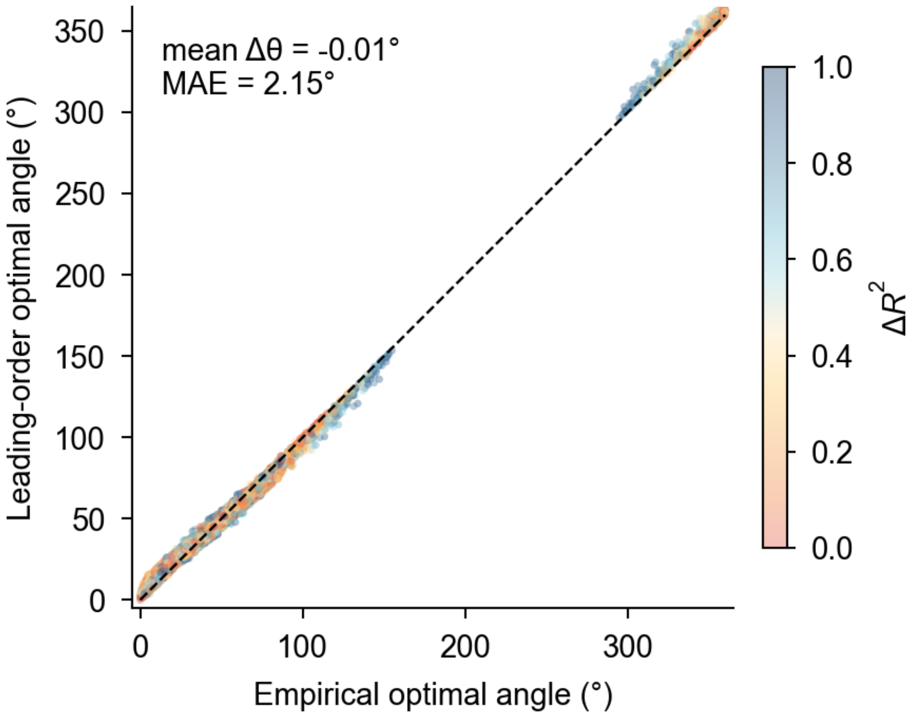


**Supplementary Fig. 7: Validation of the optimal PRS combination angle in simulations.**

Leading-order optimal angle $\theta_{\mathrm{lo}}$ (Supplementary Note 2.9) plotted against the empirical optimal angle. Angles specify the relative weights of the two subtype-specific PRSs in the case–case discriminant. For weights $(w_1, w_2)$ in the contrast of Supplementary Note 2.4, oriented so that subtype-1 cases score higher on average, the angle is $\operatorname{arctan2}(w_2, w_1)$ in [0°, 360°). $\theta_{\mathrm{lo}}$ is the angle of the leading-order weights of Supplementary Note 2.9. Empirical angles were obtained by sample-split grid search and averaged over 30 replicates using the circular mean. Each point represents one of 20,736 parameter settings and is colored by the difference in subtype-specific PRS accuracies, $\Delta R^2 = |R_1{}^2 - R_2{}^2|$. Dashed lines denote identity. Angles and errors are reported in degrees; mean Δθ is the mean circular difference between empirical and predicted angles, and MAE is the mean absolute circular error. No statistical tests were performed.

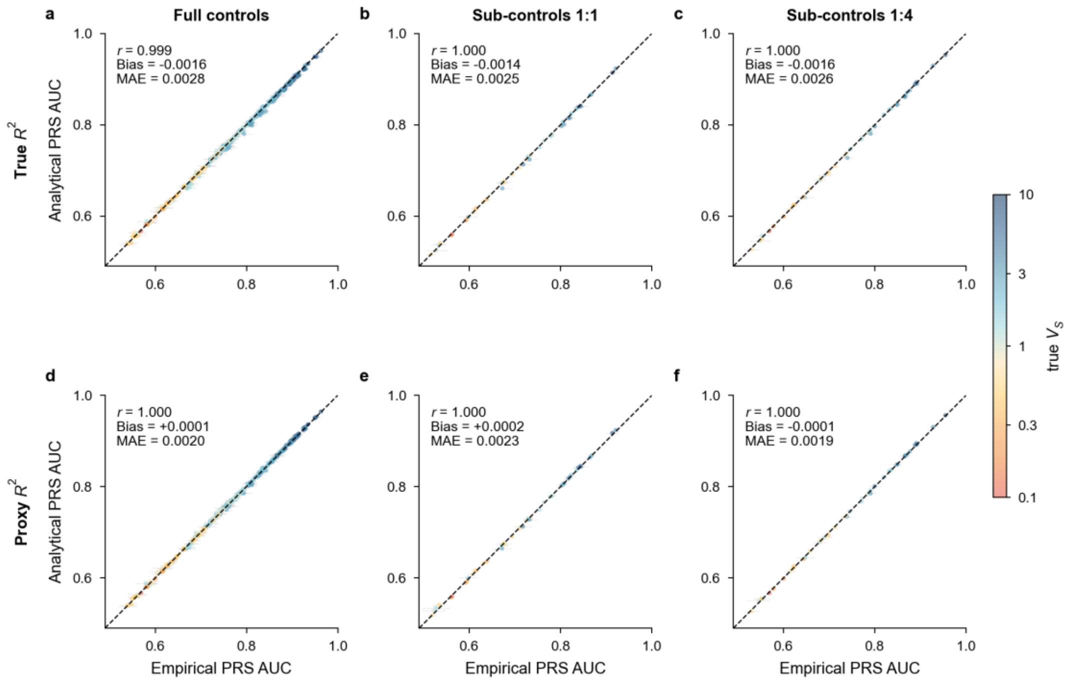


**Supplementary Fig. 8: Prediction of PRS-based case–case AUC across control sampling schemes.**

Validation of the analytical PRS formula in the genotype-based simulations under a cross-fitted evaluation. The held-out test set was split once, by individual, into two halves; in each half the scores were oriented, their accuracies estimated and the leading-order weights of Supplementary Note 2.9 formed, and the resulting discriminant was evaluated in the other half; the two evaluation-half AUCs were averaged with weights proportional to the number of case pairs, so that no weight was chosen on the individuals it was evaluated in. Columns show full controls (**a**,**d**), 1:1 case–control sampling (**b**,**e**) and 1:4 case–control sampling (**c**,**f**). The analytical PRS AUC, from the moment-based approximation of Supplementary Note 2.7 at the leading-order weights with the true simulation parameters, is plotted against the empirical AUC of the discriminant at those weights. In the top row (**a**–**c**) the score accuracies were computed as the squared correlation with the true genetic values; in the bottom row (**d**–**f**) they were inferred from the scores' case–control AUCs as in the real-data analysis (Supplementary Note 3.5.1). Within each row the same accuracy route enters the weights, estimated in the weight half, and the analytical AUC, estimated in the full test set. Each point represents one parameter setting, averaged over ten replicates: 324 settings for full controls (**a**,**d**) and the 45-setting sensitivity subset described in Supplementary Fig. 5 for the subsampled designs (**b**,**c**,**e**,**f**). Horizontal error bars denote ±1 s.d. of the empirical AUC across replicates, and points are colored by true $V_S$. Dashed lines denote identity; $r$ denotes the Pearson correlation, bias the mean of analytical minus empirical AUC and MAE the mean absolute error. No statistical tests were performed.

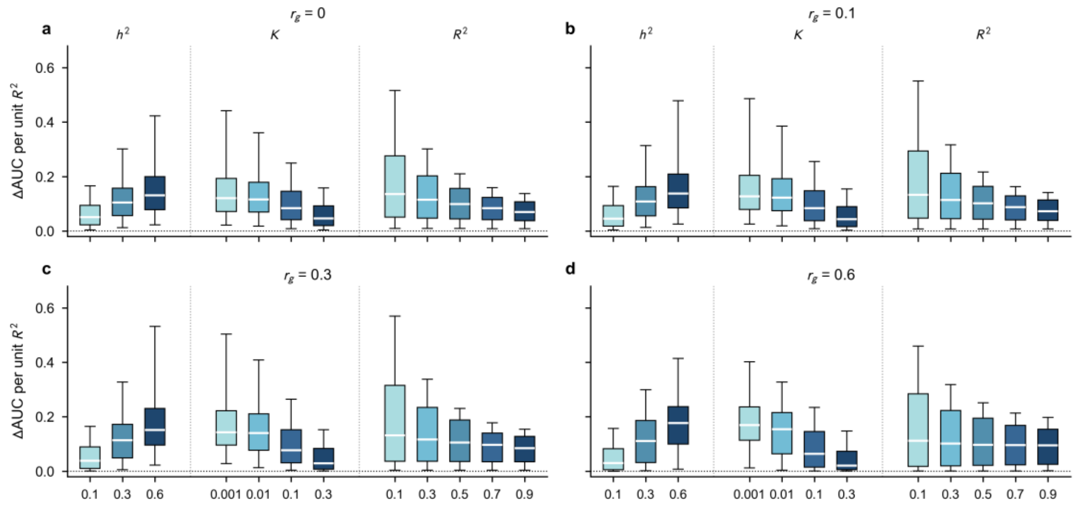


**Supplementary Fig. 9: Marginal gains in PRS-based case–case AUC by genetic correlation.**

Gain in case–case AUC per unit increase in subtype-specific PRS accuracy, $\Delta AUC/\Delta R^2$, at cross-subtype genetic correlation **a,** $r_g = 0$; **b,** 0.1; **c,** 0.3; and **d,** 0.6. As in Fig. 2d, gains are calculated as finite differences between adjacent $R^2$ grid levels while holding all other parameters fixed, and stratified by the improved subtype's heritability ($h^2$), prevalence ($K$) and baseline PRS accuracy ($R^2$). Boxes show interquartile ranges, white lines mark medians, and whiskers extend to the 5th and 95th percentiles; outlying values are omitted. Shading runs from light to dark across the levels of each group. Each panel contains 8,640 one-step increments; individual boxes summarize 2,880 ($h^2$), 2,160 ($K$) or 1,728 ($R^2$) values. Dotted horizontal lines denote zero gain. Boxes summarize variation across the parameter grid; no statistical tests were performed.

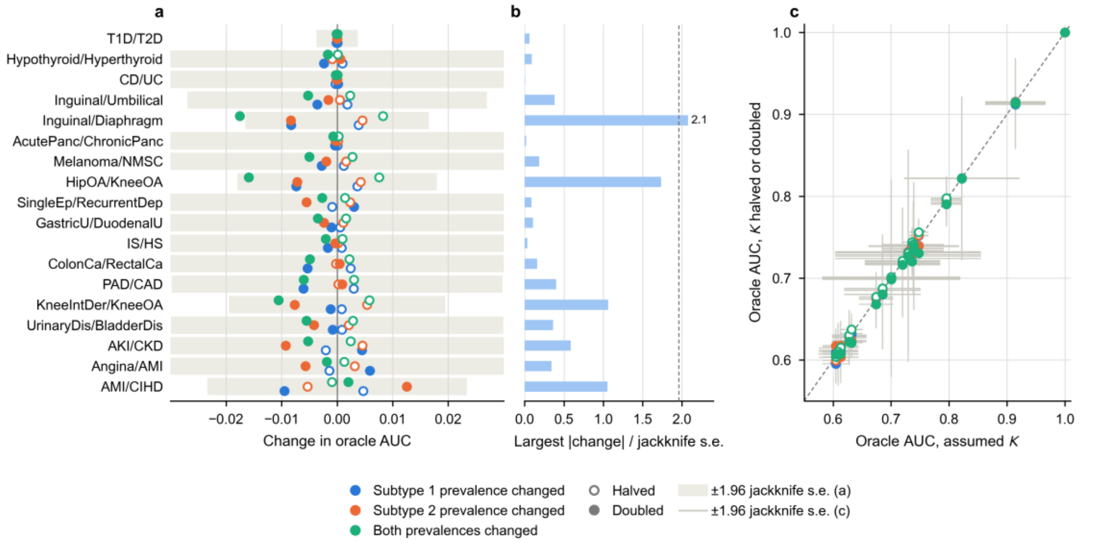


**Supplementary Fig. 10: Sensitivity of the oracle AUC to the assumed population prevalence.**

**a,** Change in the UK Biobank oracle AUC of each subtype pair when the population prevalence $K$ of subtype 1 (blue), subtype 2 (orange) or both subtypes (green) is halved (open circles) or doubled (filled circles), with the observed-scale heritabilities, case fractions and genetic correlation held fixed (Supplementary Note 3.3.3). Grey bands show ±1.96 block-jackknife s.e. of the baseline oracle AUC. Pairs are ordered by oracle AUC. **b,** Largest absolute change for each pair divided by its jackknife s.e.; the dashed line marks 1.96 s.e., a descriptive reference and not a test of the difference. **c,** Oracle AUC under each perturbed prevalence plotted against the value at the assumed prevalence (108 recomputations); error bars are ±1.96 block-jackknife s.e. on both axes, the vertical ones from GenSep re-run under the perturbed prevalence so that the 200-block jackknife is recomputed, and the dashed line is the identity.

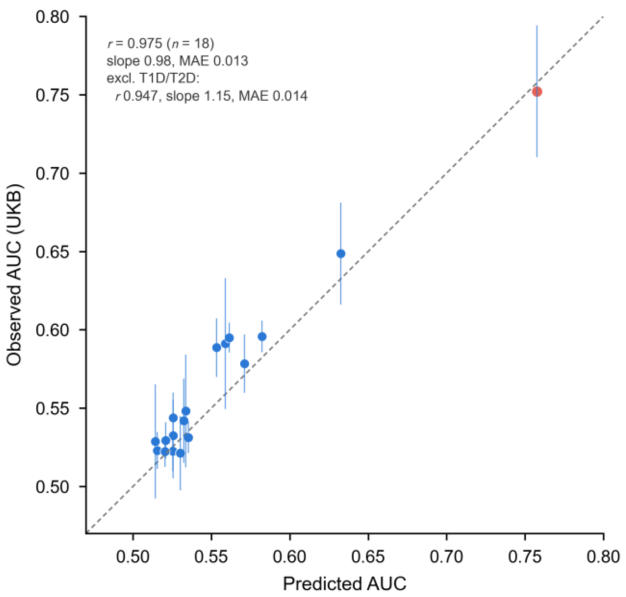


**Supplementary Fig. 11: Predicted and observed PRS-based case–case AUCs in UK Biobank.**

Observed case–case AUC plotted against the analytical prediction for 18 subtype pairs. Predictions use the estimated liability-scale SNP heritabilities, genetic correlation, population prevalences and subtype-specific PRS accuracies inferred from case–control AUCs (Methods). Observed AUCs were evaluated in the 20% held-out UK Biobank test set by two-fold cross-fitting at the leading-order weights, the weights being formed in one half and the AUC read in the other (Methods). Error bars denote ±1.96 s.e. from a delete-a-block jackknife over test participants (Methods). Dashed lines denote identity; type 1 versus type 2 diabetes (T1D/T2D) is highlighted in red. The panel reports the Pearson correlation, calibration slope of observed on predicted AUC and mean absolute error (MAE), with and without T1D/T2D. These summaries are descriptive and were not tested.

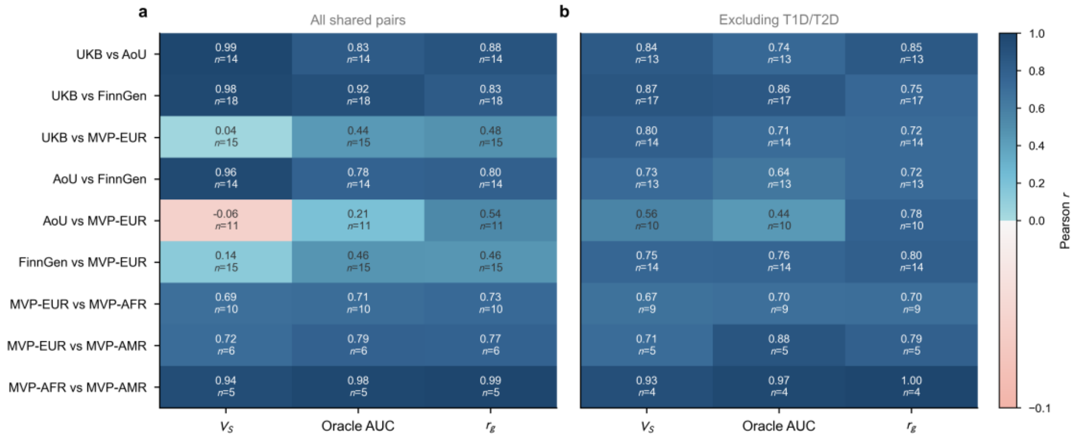


**Supplementary Fig. 12: Agreement in genetic separability across cohorts and ancestries.**

Pearson correlations between analyses for genetic separation variance, $V_S$, oracle AUC and cross-subtype genetic correlation, $r_g$. **a,** All subtype pairs with reliable estimates in both analyses (Methods). **b,** The same comparisons after excluding type 1 versus type 2 diabetes (T1D/T2D). Rows show comparisons among UK Biobank (UKB), All of Us (AoU), FinnGen and the European-ancestry Million Veteran Program analysis (MVP-EUR), followed by cross-ancestry comparisons within MVP involving the African-ancestry (MVP-AFR) and Admixed American (MVP-AMR) analyses. Cell shading indicates the Pearson correlation; each cell reports the correlation and the number of shared subtype pairs. Correlations are descriptive and were not tested. Spearman correlations and numerical results are reported in Supplementary Table 5; sensitivity of T1D/T2D estimates to subtype definition is examined in Supplementary Fig. 13.

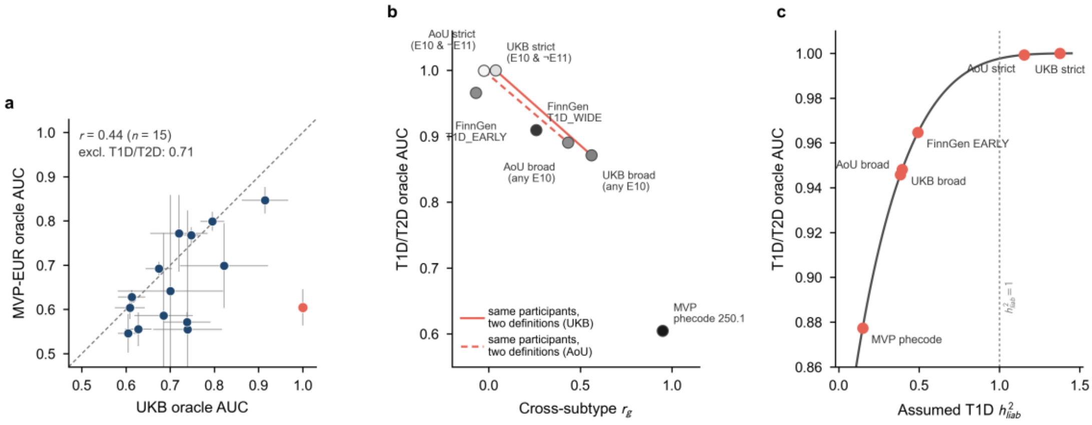


**Supplementary Fig. 13: Agreement between MVP European and UK Biobank, and sensitivity of the T1D/T2D oracle AUC to subtype definition and heritability.**

**a,** Oracle AUC in the European-ancestry MVP analysis (MVP-EUR) versus UK Biobank (UKB) for the 15 subtype pairs with reliable estimates in both analyses, with type 1 versus type 2 diabetes (T1D/T2D) in red; the dashed line denotes identity, error bars denote ±1.96 block-jackknife s.e., and *r* is the Pearson correlation over all pairs and after excluding T1D/T2D. **b,** Oracle AUC for T1D/T2D plotted against cross-subtype genetic correlation, $r_g$, under seven cohort–definition combinations: FinnGen T1D_EARLY (2,960 cases), All of Us strict (364 cases) and UK Biobank strict (688 cases; both E10 without E11 or E12–E14), FinnGen T1D_WIDE (11,197 cases), All of Us broad (2,883 cases) and UK Biobank broad (3,502 cases; both any E10, with the T2D arm likewise any E11) and MVP phecode 250.1 (16,971 cases). Marker shading indicates $\log_{10}$ T1D case count. The red lines connect the two definitions applied within UK Biobank (solid) and within All of Us (dashed); 77% and 86% of cases under the broad definition, respectively, also carry E11. **c,** Oracle AUC as a function of assumed T1D liability-scale SNP heritability, holding T2D heritability, genetic correlation and both prevalences at their UK Biobank values. Red points mark T1D heritability estimates from the labeled sources, used as inputs with the other parameters fixed at their UK Biobank values; they are sensitivity calculations, not the cohort-specific AUC estimates of panel **b**. The dotted vertical line marks $h^2 = 1$. Estimates above 1 and the sensitivity analysis are discussed in Supplementary Note 3.4.3. Definition-specific estimates are reported in Supplementary Table 6. No statistical tests were performed.

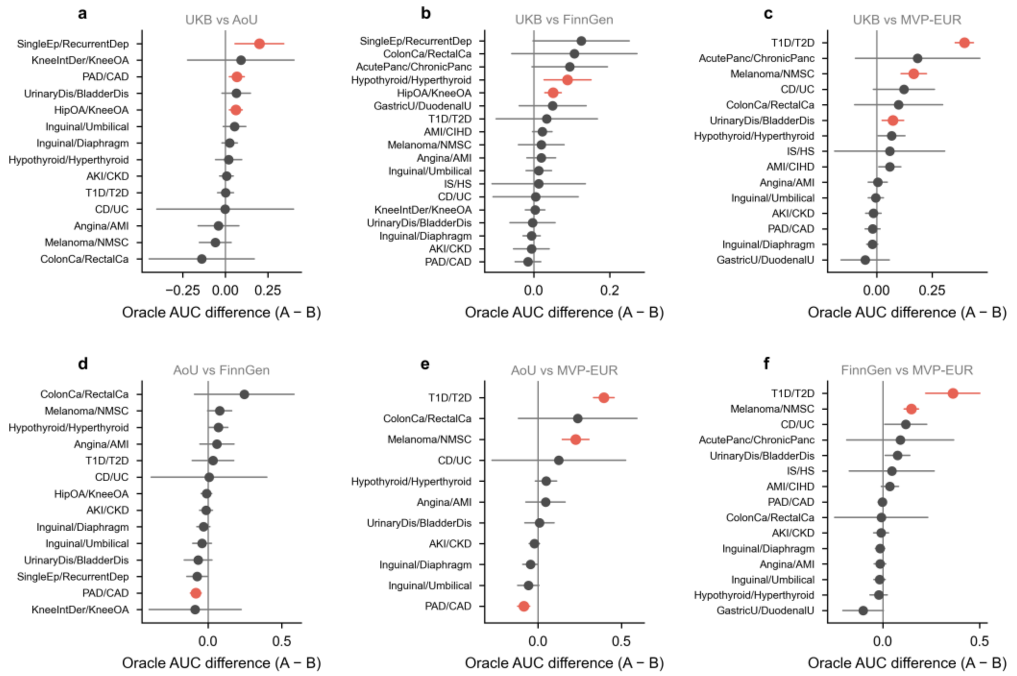


**Supplementary Fig. 14: Differences in oracle AUC across European-ancestry cohorts.**

Per-pair oracle AUC differences between **a,** UK Biobank and All of Us; **b,** UK Biobank and FinnGen; **c,** UK Biobank and MVP European; **d,** All of Us and FinnGen; **e,** All of Us and MVP European; and **f,** FinnGen and MVP European. Differences are calculated as the first analysis minus the second (A − B), restricted to subtype pairs with reliable estimates in both analyses and ordered by the difference within each panel. Error bars denote ±1.96 s.e.; variances are summed across the two independent analyses, using s.e. from 200-block genomic block-jackknife estimates. Vertical lines denote zero difference. Red points and intervals indicate Benjamini–Hochberg-adjusted $q < 0.05$ from two-sided $z$-tests, with correction within each cohort comparison and quantity. Panels contain 11–18 shared pairs, yielding 87 differences, of which 14 pass correction. Apart from type 1 versus type 2 diabetes, three pairs differed in more than one comparison, in each case because one cohort differed from all others in which the pair was analyzed: hip versus knee osteoarthritis (higher in UK Biobank), melanoma versus non-melanoma skin cancer (lower in MVP European) and peripheral artery versus coronary artery disease (lower in All of Us). Exact $P$ and $q$ values, together with results for $V_S$ and $r_g$, are reported in Supplementary Table 7.

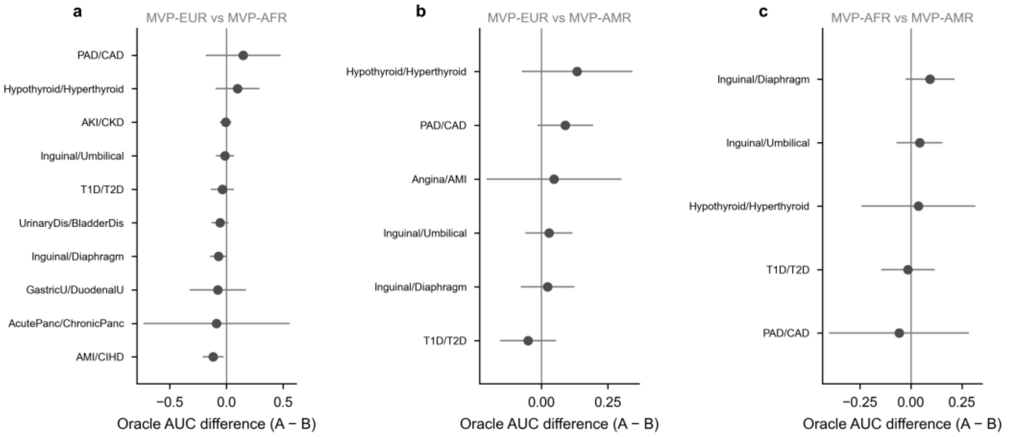


**Supplementary Fig. 15: Differences in oracle AUC across ancestries within MVP.**

Per-pair oracle AUC differences between MVP analyses of **a,** European and African ancestry; **b,** European and Admixed American ancestry; and **c,** African and Admixed American ancestry. Differences are calculated as the first analysis minus the second (A − B), holding the cohort and phecode-based phenotyping system constant. Panels include 10 (**a**), 6 (**b**) and 5 (**c**) subtype pairs with reliable estimates in both analyses. Error bars denote ±1.96 s.e., calculated from the sum of the two 200-block genomic jackknife variances; vertical lines denote zero difference. Two-sided *z*-tests were corrected using the Benjamini–Hochberg procedure within each ancestry comparison and quantity. None of the 21 oracle AUC differences has Benjamini–Hochberg-adjusted $q < 0.05$. Exact *P* and *q* values are reported in Supplementary Table 7. The small numbers of eligible pairs and imprecise estimates limit sensitivity to ancestry differences.

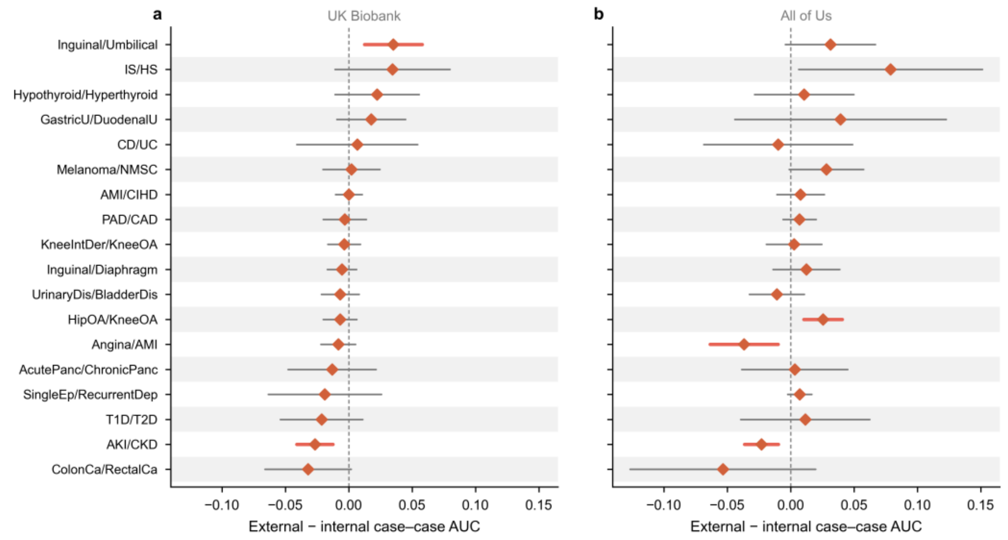


**Supplementary Fig. 16: Differences in case–case AUC between FinnGen-trained and internally trained PRSs.**

FinnGen-trained minus internally trained PRS-based case–case AUC for 18 subtype pairs in **a,** UK Biobank and **b,** All of Us. Error bars denote ±1.96 s.e. from a paired delete-a-block jackknife over the target cohort's held-out test individuals, with both scores re-evaluated within each deletion (Methods). Dashed vertical lines denote zero difference. Red, heavier intervals indicate Benjamini–Hochberg-adjusted $q < 0.05$ from two-sided $z$-tests, with correction across the 18 pairs separately for each target cohort. Both panels use the same horizontal scale and pair order, sorted by the difference in UK Biobank. Five of the 36 differences pass correction. In UK Biobank, inguinal versus umbilical hernia favors the FinnGen-trained score (+0.035) and acute kidney injury versus chronic kidney disease favors the internally trained score (−0.027). In All of Us, hip versus knee osteoarthritis favors the FinnGen-trained score (+0.026), whereas acute kidney injury versus chronic kidney disease (−0.023) and angina versus acute myocardial infarction (−0.037) favor the internally trained score. Exact $P$ and $q$ values are reported in Supplementary Table 8.

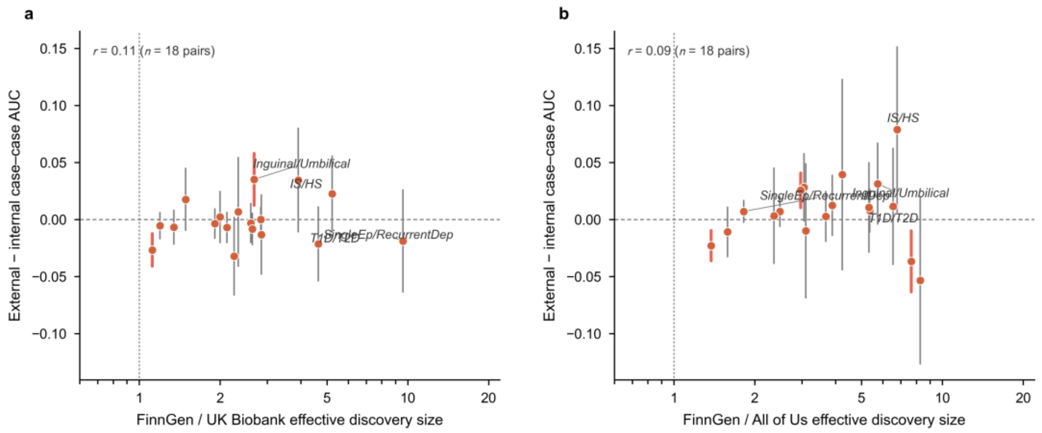


**Supplementary Fig. 17: Discovery sample size and gains from FinnGen-trained PRSs.**

FinnGen-trained minus internally trained PRS-based case–case AUC plotted against the ratio of effective discovery sample sizes in **a,** UK Biobank and **b,** All of Us. Each point represents one of 18 subtype pairs. For each subtype, effective discovery size is $N_{\mathrm{eff}} = 4N_{\mathrm{case}}N_{\mathrm{ctrl}}/(N_{\mathrm{case}} + N_{\mathrm{ctrl}})$; the plotted ratio is the geometric mean of the two subtype-specific FinnGen-to-internal ratios, shown on a logarithmic scale. FinnGen effective sample sizes were computed from the endpoint case and control counts in Supplementary Table 1; internal sizes are those of the training GWAS each score was built from (the 80% training split): the case count of each arm and the number of participants in that GWAS, taken in UK Biobank from the analysis output and the per-variant sample size of the training summary statistics, and in All of Us from the per-arm counts of the training jobs, which equal the per-variant sample size of the training summary statistics. AUC differences and error bars (±1.96 paired-jackknife s.e.) are those in Supplementary Fig. 16. Dotted vertical lines denote equal discovery size, and dashed horizontal lines denote zero AUC difference. Red, heavier intervals indicate $q < 0.05$ using the tests and correction in Supplementary Fig. 16. Each panel reports the Pearson correlation between the AUC difference and log-transformed sample-size ratio; correlations are descriptive and were not tested.

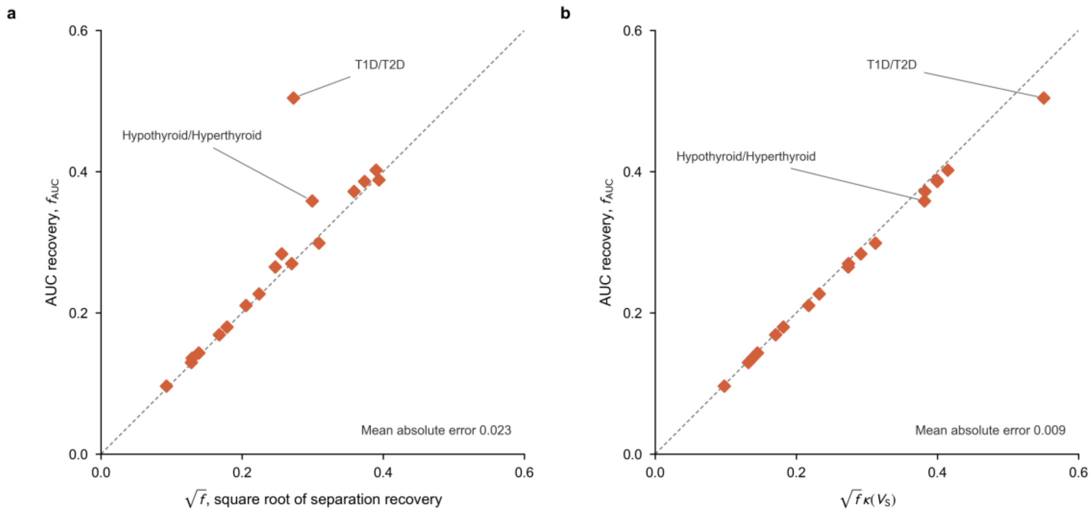


**Supplementary Fig. 18: Relation between AUC recovery and separation recovery in UK Biobank.**

**a,** AUC recovery, $f_{\mathrm{AUC}} = (\mathrm{AUC}_{\mathrm{PRS}} - 0.5)/(\mathrm{AUC}_{\mathrm{oracle}} - 0.5)$, plotted against the square root of separation recovery, $\sqrt{f}$, with $f = V_{\mathrm{PRS}}/V_{\mathrm{S}}$. **b,** AUC recovery plotted against $\sqrt{f}\kappa(V_{\mathrm{S}})$, where $\kappa(V_{\mathrm{S}}) = \sqrt{V_{\mathrm{S}}/\pi}/\mathrm{erf}\left(\sqrt{V_{\mathrm{S}}}/2\right)$ (Supplementary Note 3.5.3). Each point represents one of 18 subtype pairs in UK Biobank. Observed AUCs are the cross-fitted held-out estimates shown in Fig. 3a, and oracle AUCs and $V_{\mathrm{S}}$ are the UK Biobank estimates (Supplementary Tables 2 and 3). Dashed lines denote identity. Each panel reports the mean absolute error from the identity line.

**Supplementary Tables**

Supplementary Tables 1–8 are provided as a separate Excel workbook, with one table per sheet and a Fields sheet describing every column; their titles and legends are given here.

**Supplementary Table 1: Subtype-pair definitions, case counts, prevalences and phenotype mapping.** One row per subtype pair. Columns give the clinical contrast and subtype classification; the criterion that distinguishes the two subtypes, with one supporting reference per pair (Supplementary Note 3.3.1); whether individuals carrying both subtype codes were removed or retained (Supplementary Note 3.3.2); for each subtype the short label, full name, defining ICD-10 codes, the population prevalence $K$ used with its source, basis and DOI or URL (Methods), and the number of cases in the full UK Biobank cohort; the number of UK Biobank controls shared by the two case–control analyses and the codes whose absence defines a control (Supplementary Note 3.3.2); the number of cases of each subtype in All of Us; the matched FinnGen DF13 endpoints with case and control counts; and the matched MVP phecodes with European-, African- and Admixed American-ancestry case counts and mapping notes, "Not available" marking subtypes that no phecode separates (Methods).

**Supplementary Table 2: Genetic separability estimates for all cohorts and ancestries.** One row per subtype pair, cohort and ancestry group. Columns give the liability-scale SNP heritability of each subtype, the cross-subtype genetic correlation, the genetic separation variance $V_{\mathrm{S}}$, the analytical oracle AUC and its simplified approximation, each with its 200-block jackknife standard error, together with the reliability flag and the reason for any failure under the rule of Supplementary Note 3.4.2, or a note on a reportable estimate (a liability heritability above 1 lies outside the $h^2 \leq 1$ domain of Supplementary Note 1). Values for the 18 UK Biobank pairs are shown in Fig. 3a and main-text Table 1.

**Supplementary Table 3: Case–case PRS AUC by discovery cohort and target cohort.** One row per subtype pair, discovery source (internal, FinnGen or published consortium GWAS) and target cohort (UK Biobank or All of Us). Columns give the observed case–case AUC (cross-fitted, leading-order weights; Supplementary Note 3.5.2) with its delete-a-block jackknife standard error, the sensitivity weighting evaluated on the same halves (grid-searched weight direction), the AUC predicted by the moment-based PRS AUC approximation (Supplementary Note 2.7) and the AUC recovery (Supplementary Note 3.5.3) computed from the observed AUC and the target cohort's oracle AUC, left blank where the target cohort has no reliable oracle

AUC (Supplementary Table 2).

**Supplementary Table 4: Training cases required to reach target case–case AUCs.** For each subtype pair: the ceiling AUC, the current number of training cases in each arm (the 80% training split), the current observed AUC and the calibration target (Supplementary Note 3.6.2) $f_{\mathrm{curr}}$, defined as the separation variance implied by the observed AUC divided by that implied by the ceiling AUC (its denominator differs from that of the separation recovery of Supplementary Note 3.5.3, which divides by $V_{\mathrm{S}}$), the accuracy proxies of the two arms, the calibration factor $\kappa_0$ (Supplementary Note 3.6.2), and the number of training cases required to reach case–case AUCs of 0.70 and 0.80 when both arms grow in proportion (total and per subtype) and when only one arm grows with the other held at its current size. "inf" denotes a target that cannot be reached under that scenario, and the status column beside each required number states whether the target is already attained at the current size (the entry then equals the current size), reached at a finite size, or unreachable because the ceiling lies below the target or because the other arm is held fixed (Fig. 3b).

**Supplementary Table 5: Cross-cohort and cross-ancestry correlations of genetic separability.** For each pair of analyses and each quantity ($V_{\mathrm{S}}$, oracle AUC and $r_{\mathrm{g}}$), the number of subtype pairs reliable in both analyses and the Pearson and Spearman correlations across them, computed over all shared pairs and after excluding type 1 versus type 2 diabetes (Supplementary Fig. 12).

**Supplementary Table 6: Sensitivity of the type 1 versus type 2 diabetes oracle AUC to the diabetes phenotype definitions.** For each of the seven cohort–definition combinations described in Supplementary Note 3.4.3: the cohort, the case definition and case count of each arm (the broad UK Biobank and All of Us definitions relax the type 2 diabetes arm together with the type 1 arm; FinnGen T1D_WIDE keeps the same type 2 diabetes endpoint), the cross-subtype genetic correlation between the two arms and the oracle AUC (Supplementary Fig. 13b).

**Supplementary Table 7: Per-pair cross-cohort and cross-ancestry difference tests.** For each comparison, subtype pair and quantity: the estimate and standard error in each analysis, their difference and its standard error, the two-sided $P$ value and the Benjamini–Hochberg $q$ value, corrected within each combination of comparison and quantity (Methods; Supplementary Figs. 14 and 15).

**Supplementary Table 8: External-minus-internal differences in case–case PRS AUC.** One row per subtype pair, external discovery source (FinnGen or published consortium GWAS) and target cohort. Columns give the internally and externally trained AUCs, their difference with its paired delete-a-block jackknife standard error over unique case participants, the weights being refitted in each deletion, the two-sided $P$ value and the Benjamini–Hochberg $q$ value corrected within each combination of discovery source and target cohort (18 pairs for FinnGen, three for the consortium GWAS; the family is stated in each row, and the consortium families include the two UK Biobank comparisons with discovery–target overlap that the main-text summary excludes), and the case counts of the two arms, and, for the FinnGen rows, the genetic correlation of each subtype between the FinnGen endpoint GWAS and the target-cohort GWAS with its standard error (SumHer, sample-overlap term estimated) (Methods; Supplementary Figs. 16 and 17). The one-sided test of each of these genetic correlations against 1 reported in the main text (24 of 36 UK Biobank and 22 of 34 All of Us subtype comparisons with $q < 0.05$) uses these estimates and standard errors, $z = (1 - \text{estimate})/\text{s.e.}$, with Benjamini–Hochberg correction within each target cohort.
